# Best Practice Methods for Living Evidence Synthesis in Health Care: An International Modified Delphi Survey

**DOI:** 10.1101/2025.11.07.25339719

**Authors:** Melanie M. Golob, Jonathan Livingstone-Banks, Per Olav Vandvik, Maria Michaels, Rebecca Hodder, Zachary Munn, James Thomas, Gabriel Rada, Gordon Guyatt, David Nunan

## Abstract

**Background:** Living evidence syntheses (LES) represent continual updates for an important topic for decision-making where there is uncertainty in the evidence. Whereas best practices for traditional evidence syntheses in health care are well established, they are not for LES. This study aimed to establish globally relevant, agreed-upon standards for considering, conducting, publishing, and implementing LES in health care.

**Methods:** A modified Delphi consensus process was conducted. Potential participants were identified through a prior survey, workshop, and targeted outreach based on expertise and representation across global organizations producing or supporting LES. Three modified Delphi rounds were administered using JISC Online Surveys between January and April 2025. Participants rated 23 statements on a five-point Likert scale. Consensus was defined as ≥80% agreement (‘agree’ or ‘strongly agree’) with ≥85% panel response rate required per round. Qualitative feedback guided iterative statement revision.

Draft statements were informed by an overview of living systematic reviews, a living evidence survey plus workshop activity, and an ongoing living critical interpretive synthesis of LES. Statements expanded upon the living methodology reporting guidance published by the PRISMA-LSR group to include other considerations for LES.

**Results:** The Delphi panel comprised 29 experts from around the world, with 27 (93%) completing round 1, 26 (90%) round 2, and 27 (93%) completing round 3; 19 of 23 statements achieved consensus. Statements described conduct (n=12), including set up and maintenance of living mode as well as funding and resources; reporting (n=2); publishing (n=4); and implementation/appraisal (n=1).

Final statements included ways to enable the living mode, such as version history; authoring tools; collaboration; unit of update; transparency; communication; publication considerations; and digital and technological considerations.

**Conclusions:** This study presents the first established consensus for best practices in considering, conducting, publishing, and implementing LES in health care. These LES standards can help align global processes, improve transparency, and promote sustainability. But there is still more to be done to accomplish these objectives, requiring that groups collaborate to embrace digital tools, adopt interoperability standards, and create effective appraisal tools.

## Introduction

No one wants clinical policy or practice informed by out-of-date evidence. This is the premise for why living evidence synthesis (LES) is becoming the norm in health research. “Living” refers to continual updates with new evidence for high priority topics where there is uncertainty. “Evidence synthesis” here means a process of systematically aggregating the health literature (e.g., systematic review, meta-analysis, scoping review, evidence map – see Appendix 1 for other definitions for this context). Whereas LES are still an emerging concept, best practices in traditional evidence syntheses in health care are well established.(1-5) These include a focused research question, systematic search and screen of the literature, data analysis, risk of bias (as applicable), grading of the evidence (as applicable), and conclusions, which may lead to recommendations or policy decisions. Traditional evidence syntheses are typically static, created without a plan for regular updates. LES, on the other hand, are meant to dynamically respond to changes in the evidence. This became particularly critical during the COVID-19 pandemic, which exemplified the need for a living approach: high priority for decision-making, uncertainty in the evidence, and new evidence likely to rapidly emerge. However, consensus on best practices specific to employing the living mode has yet to be established. This is important to establish, as a previous overview of living systematic reviews (LSRs) showed there is a wide variation in why and how LSRs are performed.(6) If there is not a common definition and threshold for what it means to be living, evidence consumers cannot be confident in the currency of the evidence. Achieving consensus on LES should help to increase trust in this type of evidence synthesis and align standards.

Progress has been made to develop LES standards. Though they were not the first to use the term “living” in reference to evidence synthesis, Cochrane was the first to publish standards and launched a pilot program in 2019.(7-11) Other research on best-practice standards for LES included considerations for living guidelines and a checklist for conducting and reporting LES.(12, 13) During the time this Delphi was conducted, a PRISMA extension was published specific to LSRs that described specific items to be included when reporting an LSR.(14) However, these other research studies did not include important aspects of LES, including looking toward the future of long-term sustainability, or taking into account such limiting factors as current peer review and publication models. The Evidence Synthesis Infrastructure Collaborative (ESIC) has formed an LES workgroup to inform living methods, and have found there are opportunities for process improvements.(15) Since their inception, groups have sought to make LES and their products – both processes and outputs (see Appendix 1 for definitions) – computer-interpretable for easy re-use and integration.(16) This research aims to bridge best practices across those processes and outputs, recognizing they are interconnected rather than isolated, creating an evidence ecosystem.(17, 18) In the absence of any consensus-based recommendations for best practices in LES, the goal of this study was to develop statements for considering, conducting, publishing, and implementing LES in global health care contexts.

A consensus method was chosen to foster a sense of ownership in the resulting best-practice statements, increasing the likelihood of implementation. The Delphi technique is an appropriate choice in health research given that LES best practices are subjective and context dependent.(19) The Delphi process is characterized by participant pseudonymity, an iterative structured questionnaire, and pre-defined thresholds for response rates and agreement.(20)

## Methods

This research was conducted and reported in concordance with ACcurate COnsensus Reporting Document (ACCORD) and Conducting and REporting DElphi Studies (CREDES) guidance.(21, 22) The protocol was posted on Open Science Framework on 19 October 2024. MMG directed the Delphi consensus process. Her experience includes facilitating 15 Delphi consensus processes for four organizations, one that was an update conducted as part of a living process. An overall flow chart summary of the Delphi process can be seen in Figure 1.

**Figure 1.**
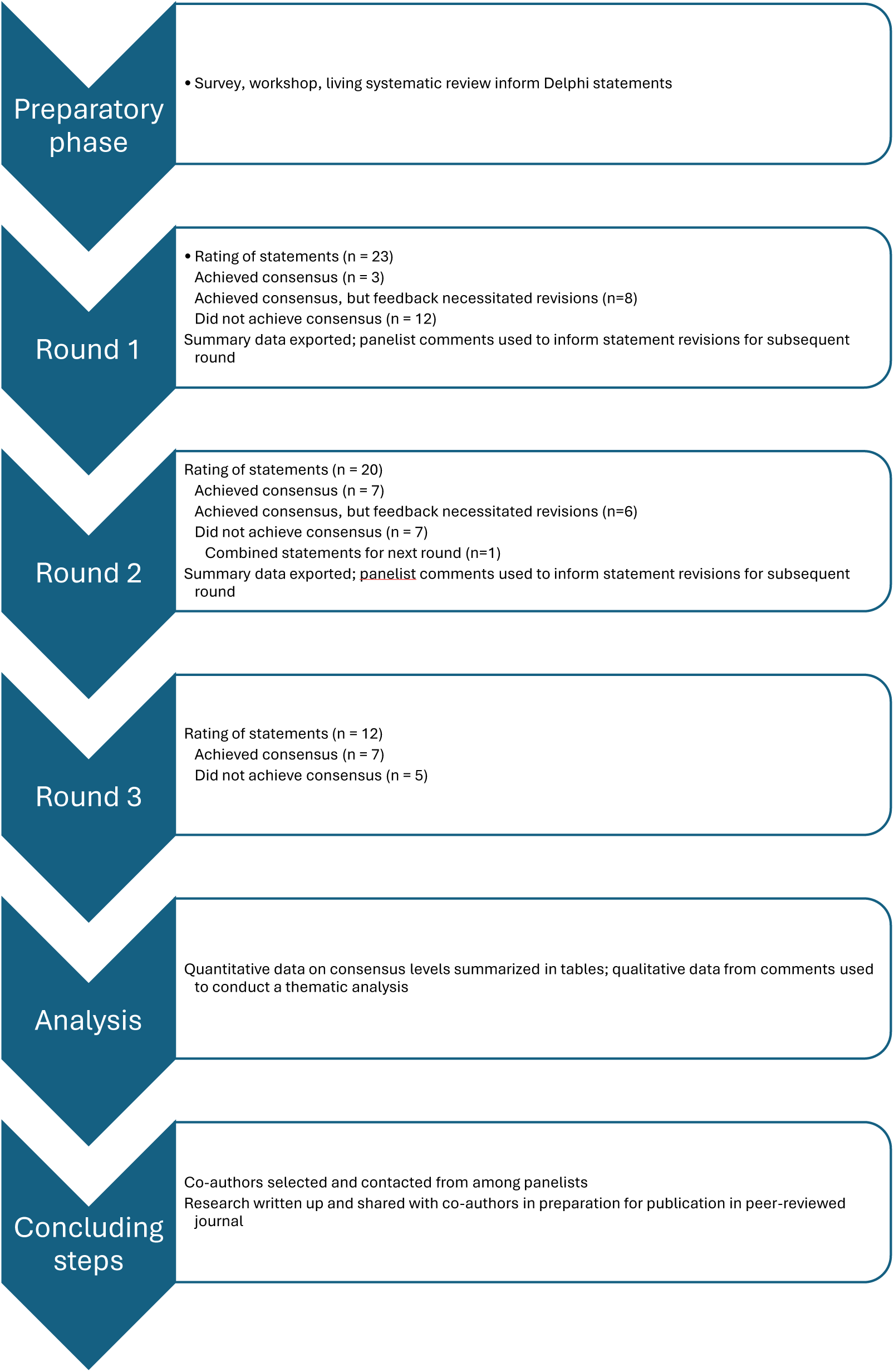
Flow chart illustrating the process of developing the set of statements for best practice methods in LES.

### Statement Development

We drafted initial LES consensus statements informed by a previous overview of LSRs, survey and workshop activity among those publishing LES or LES methods, as well as an ongoing living critical interpretive synthesis of LES.(6, 23, 24) The survey was an online questionnaire from August 2024 that sought to determine facilitators and barriers to conducting and reporting LES; the workshop was held at the 2024 Global Evidence Summit in Prague, presenting survey results and gathering participant input on which aspects of LES require consensus. When drafting statements, existing guidance on employing the living mode was considered to avoid duplication.(7, 12, 13, 25-27) Statements were designed to be able to stand alone for potential re-use. Participants received and agreed to informed consent (see Appendix 2). This information was summarized in the initial email to participants.

Within LES, the reporting of systematic reviews that adopt a living methodology has been addressed by the PRISMA-LSR group(14); however, elements relevant to reporting of other LES types were not included in the reporting guidance, so were thus included in this Delphi process. The questionnaire was piloted after development by testing with JLB.

### Delphi Survey

We conducted a modified, three-round Delphi survey in the form of a questionnaire to obtain expert and stakeholder input on best-practice methods for health-care LES. In December 2024, the Delphi director (MMG) invited participants based on LES experience; organizational role (e.g., government, academia, companies developing LES software); involvement in digital evidence solutions; and availability, while aiming for geographic and national socioeconomic diversity and limiting representation to up to three individuals per organization. Candidates came from three sources: prior LES survey respondents who opted in (n=9, of 29 respondents); LES workshop attendees (n=7, of 9 participants); and authors identified from published LES studies (n=13, of 25 authors emailed). Members of the public, patients, and carers were excluded unless actively involved in LES. The target panel size was 30–40. Appendix 3 lists organizations considered, their representation on the panel, and associated LES software platforms. There is no standard level of agreement for consensus in health research.(20) Evidence-based clinical practice guidelines typically involve up to three rounds, with a minimum response rate of 80-85% and agreement of 80% or higher to reach consensus.(28-30)

Rounds of the Delphi survey lasted approximately two weeks each, allowing time to complete the questionnaire and revise statements between rounds. A response rate of ≥85% was required, though participants could request a time extension as needed. The survey was administered via JISC Online Surveys, a web-based survey management tool; all materials were in English. Participants rated 23 statements on a 5-point Likert scale (options were ‘Strongly Agree’, ‘Agree’, ‘Neutral’, ‘Disagree’, and ‘Strongly Disagree’; see Appendix 4). The Neutral option was provided for those who did not feel comfortable providing a response for a particular statement; it was noted that it should rarely be used. A comment box was provided following each question for suggested additions, removals, or revisions for subsequent Delphi rounds as needed, or if there were topic areas or statements potentially omitted. Though identities of participants were known, survey responses and comments were anonymous and deidentified, making overall responses pseudonymized.

Statements achieving 80% or higher agreement were considered as having achieved consensus; agreement was defined as a vote of ‘Strongly Agree’ or ‘Agree’. Statements achieving below 80% agreement were modified based on comments and re-submitted in subsequent rounds of Delphi voting. Statements that achieved consensus could be minimally edited for clarity without passing through another round of the Delphi, provided the edits did not substantially change the statement that achieved consensus; if more substantial changes were warranted based on participant comments, statements could be edited and voted upon in another Delphi round. If they did not achieve consensus in the subsequent round, statements were permitted to revert to the previous iteration that did achieve consensus. If all statements received greater than 80% agreement at any point before the end of the 3rd round, subsequent rounds were unnecessary. Statements not achieving consensus after three rounds were marked that consensus was not reached.

### Informed consent and ethical approval

Informed consent was obtained via a checkbox in the questionnaire (see Appendix 2 for Delphi informed consent); participants could withdraw at any point in the Delphi prior to publication of the research for any reason. Participants were not excluded due to conflicts of interest (COI), as a variety of different perspectives and interests were sought for this research; however, participants were asked to disclose any COI for transparency (see Appendix 5 for COI form completed by participants, and Appendix 6 for completed COI). Participants were not provided incentives for participating, but were sent reminder emails if they did not participate in a Delphi round.

This Delphi process has received research ethics approval from the University of Oxford (reference: OUDCE C1A 24 36).

### Data analysis

Summary data were exported from JISC Online Surveys into an Excel file after each round for analysis. Comments for those statements that did not reach consensus were used to revise statements for the subsequent round. There were also statements that achieved consensus where comments indicated changes were warranted. These were also edited and resubmitted in the next Delphi round. Participants were shown the revised statement as well as the previous statement to see what edits had been made when voting.

Data were analyzed both quantitatively and qualitatively. Quantitative data on statement consensus levels were organized into tables and summarized.

## Results

Twenty-nine participants with LES experience were included in the Delphi consensus process from January through April 2025: Participants represented a variety of organizations (see Appendix 3) and geographic locations (see Figure 2), though most were from high-income countries.

**Figure 2.**
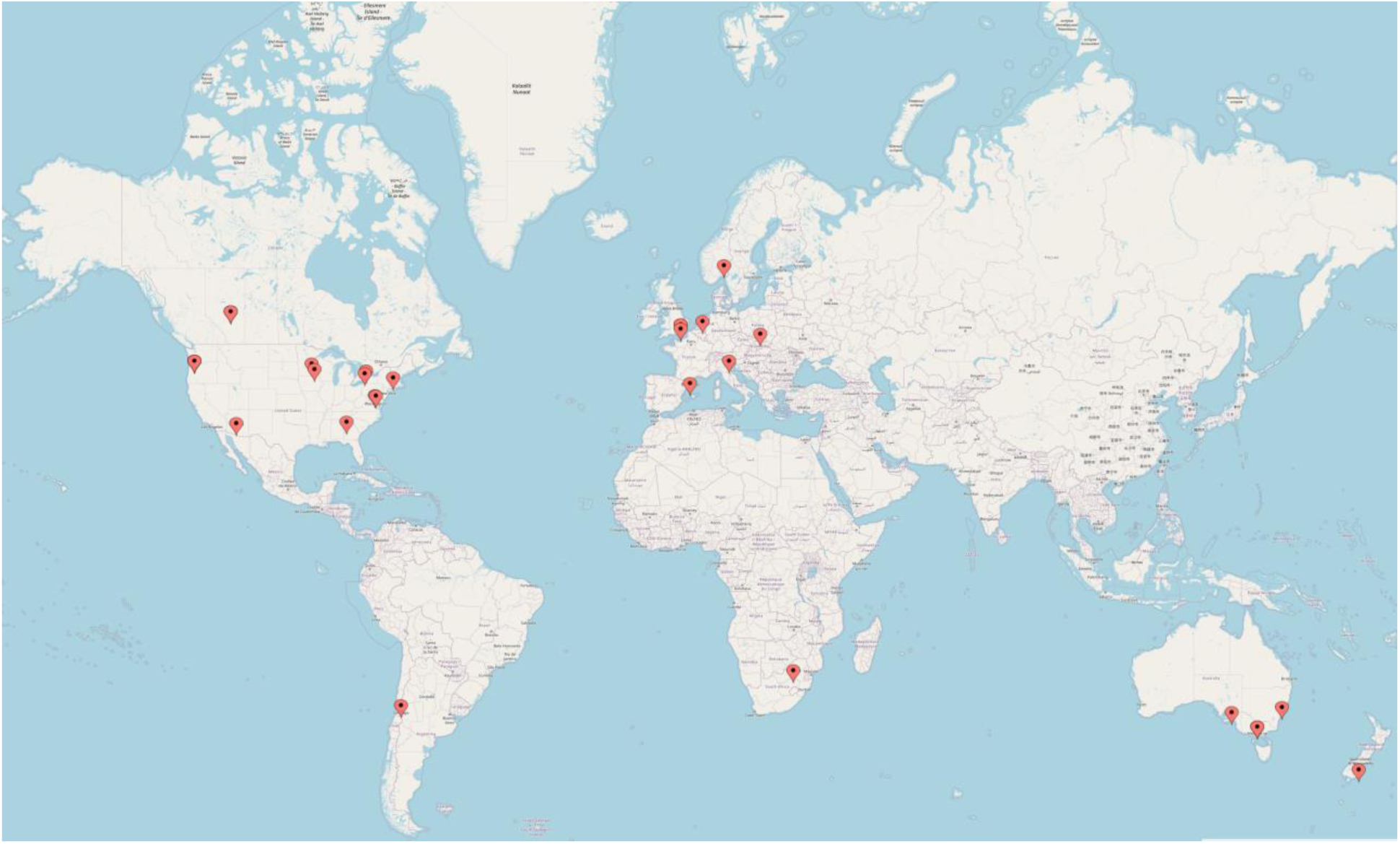
Geographic representation of participants in the Delphi survey.

The duration of each round was longer than the projected two weeks to meet the minimum response threshold of 85%. More time was also required between rounds (up to 16 days rather than the expected seven), as the volume of comments and edits was greater than anticipated. Round 1 lasted 20 days (extended from 17 days to meet minimum response rate), with eight days for editing statements based on comments; Round 2 lasted 21 days (extended from 11 days to meet minimum response rate), with 16 days for revisions; Round 3 was 20 days (extended from 13 days to meet minimum response rate).

Round 1 yielded a 93% response rate (27 of 29 participants voted). Among 23 statements, 11 reached consensus and 12 did not. No additional statements were suggested; however, based on feedback, 20 statements were revised. Round 2 had a 90% response rate (26/29). Thirteen of 20 statements reached consensus; seven did not. Due to comments, six of the 13 were revised and two statements were merged, resulting in 12 statements for voting. Round 3 had a 93% response rate (27/29). Seven of 12 statements reached consensus; five did not. Two of the five non-consensus statements had achieved consensus in round 2, and thus reverted back to the previous versions.

### Consensus statements for best practice LES

Nineteen total statements reached consensus following three rounds of the Delphi process, out of an initial 23 statements. Table 1 presents the final set of LES best practice statements. Final statements concerned ways to enable the living mode, such as version history; authoring tools; collaboration; unit of update; transparency; communication; publication considerations; and digital and technological considerations. See Appendix 1 for definitions.

Most statements (n=12) concerned LES conduct, including how to set up LES (n=8). Seven of the 19 statements were on publishing, reporting, and conduct of LES (see Table 2).

**Table 1.**
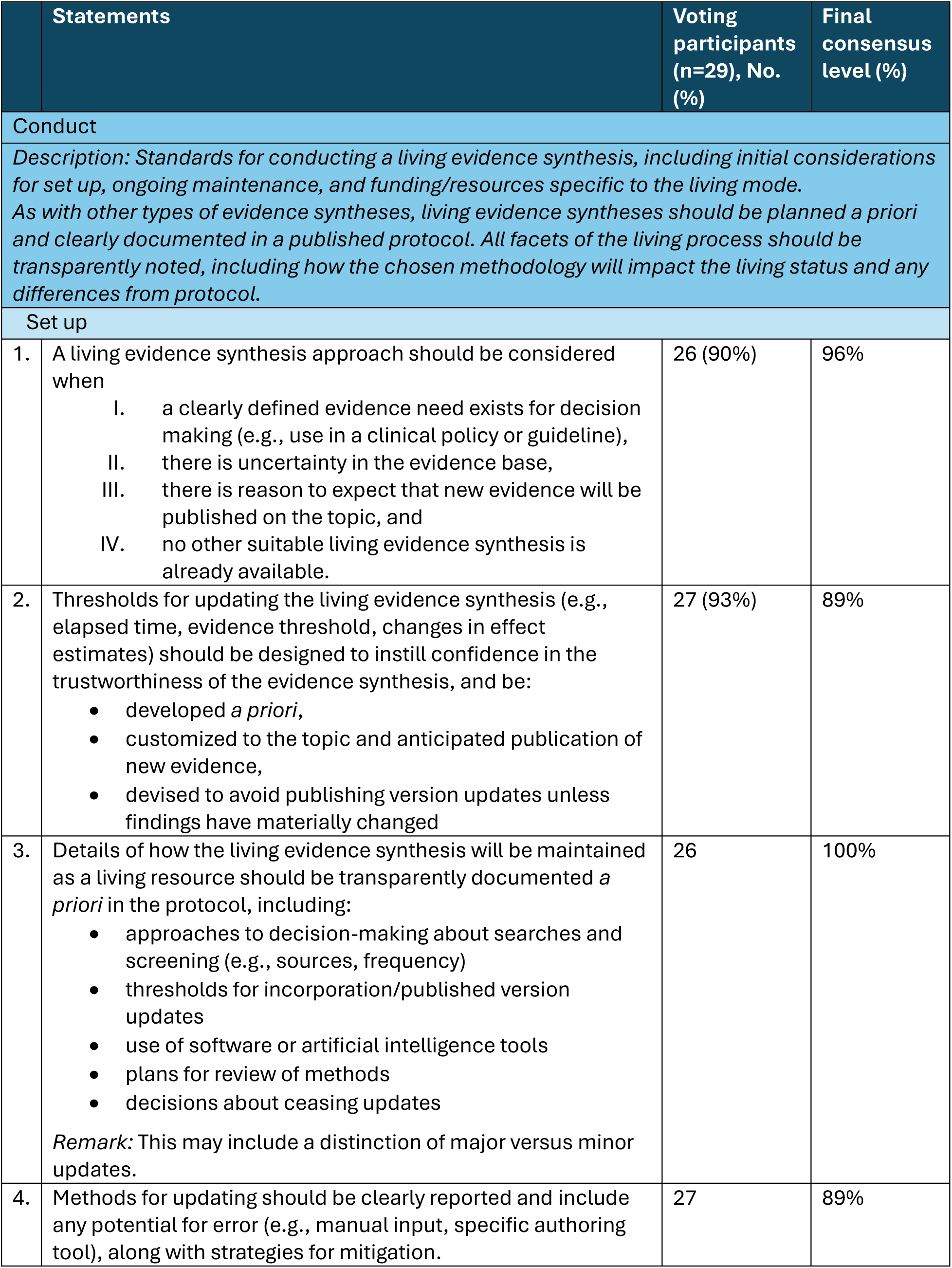
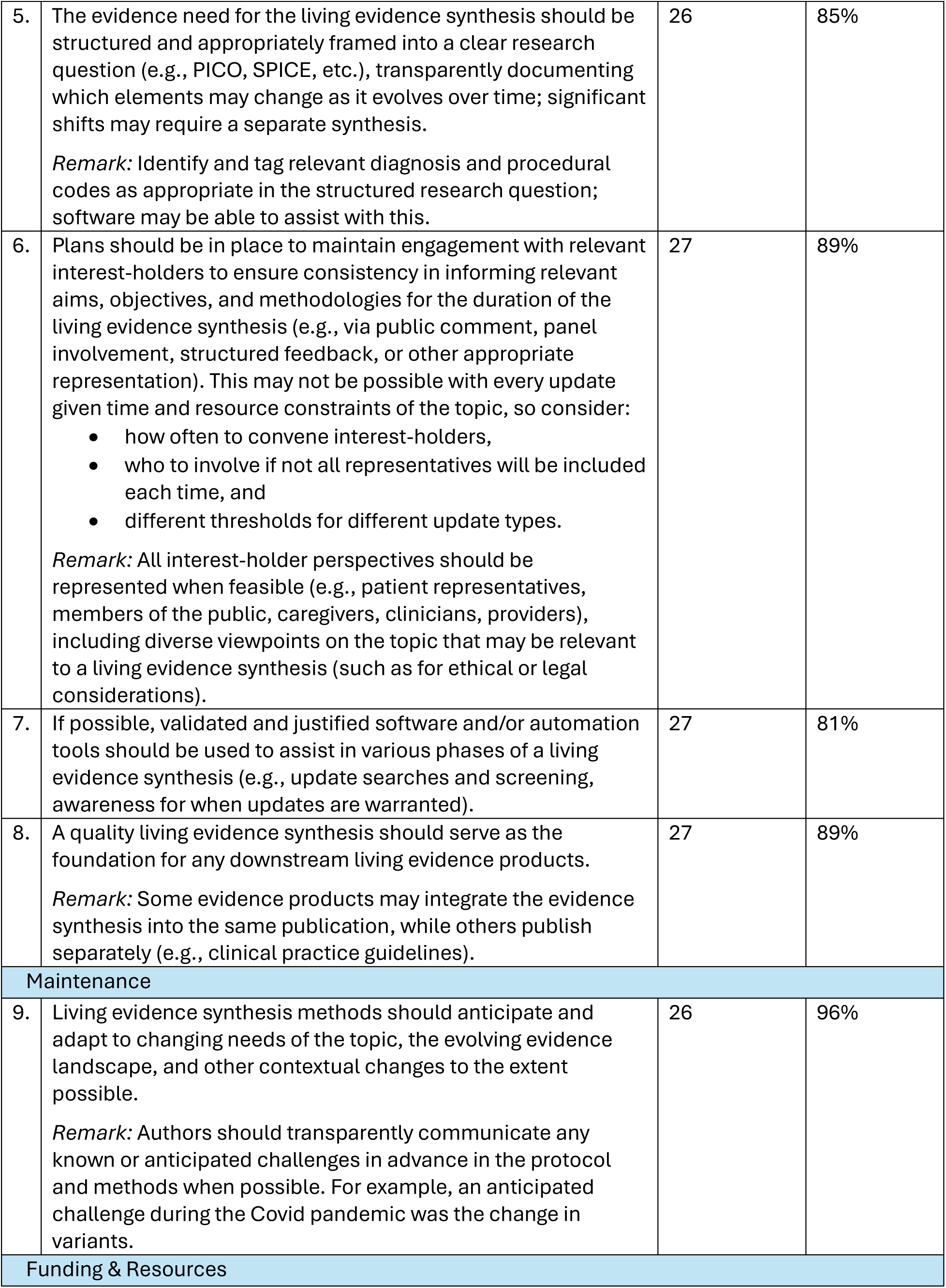
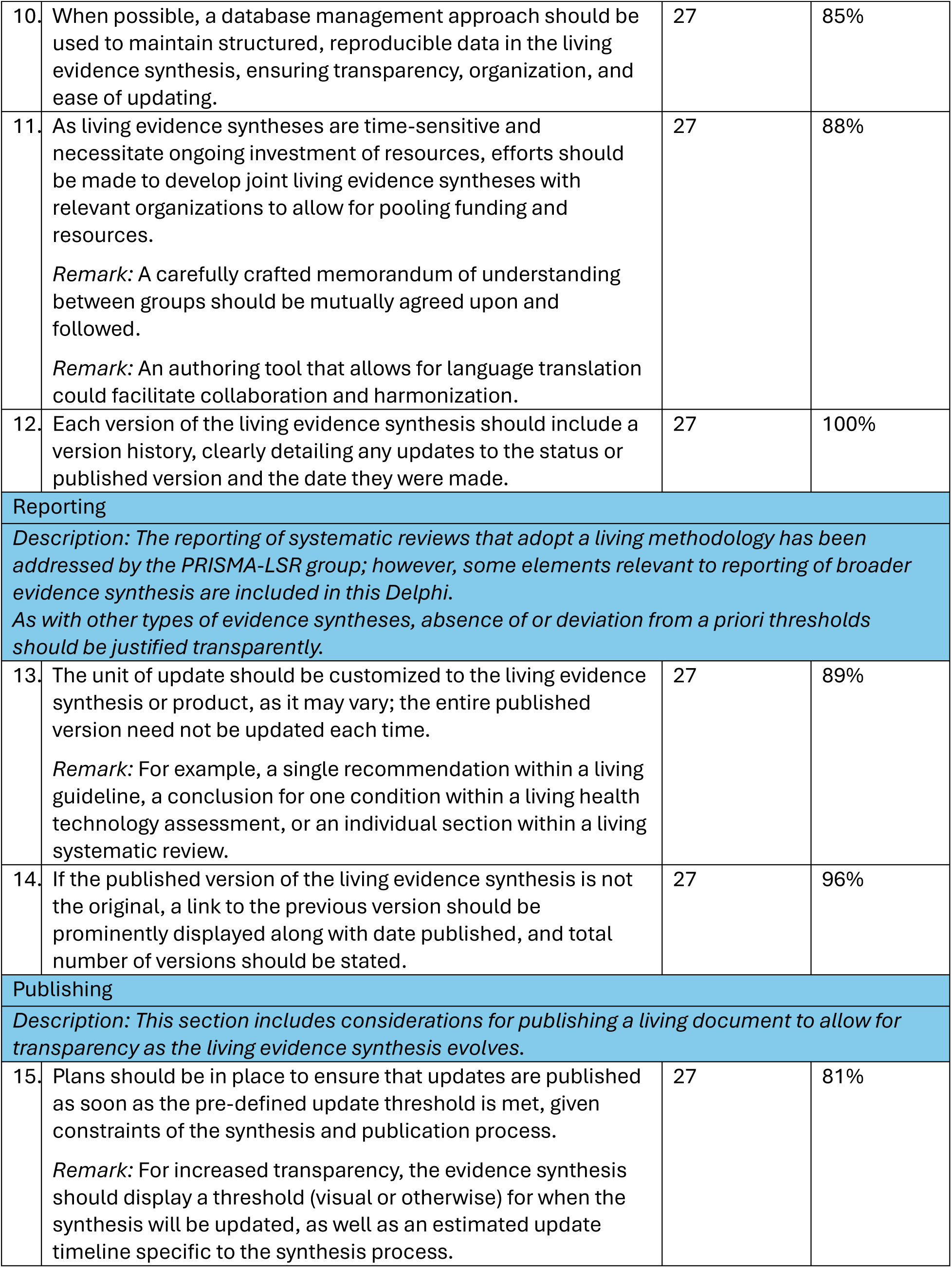
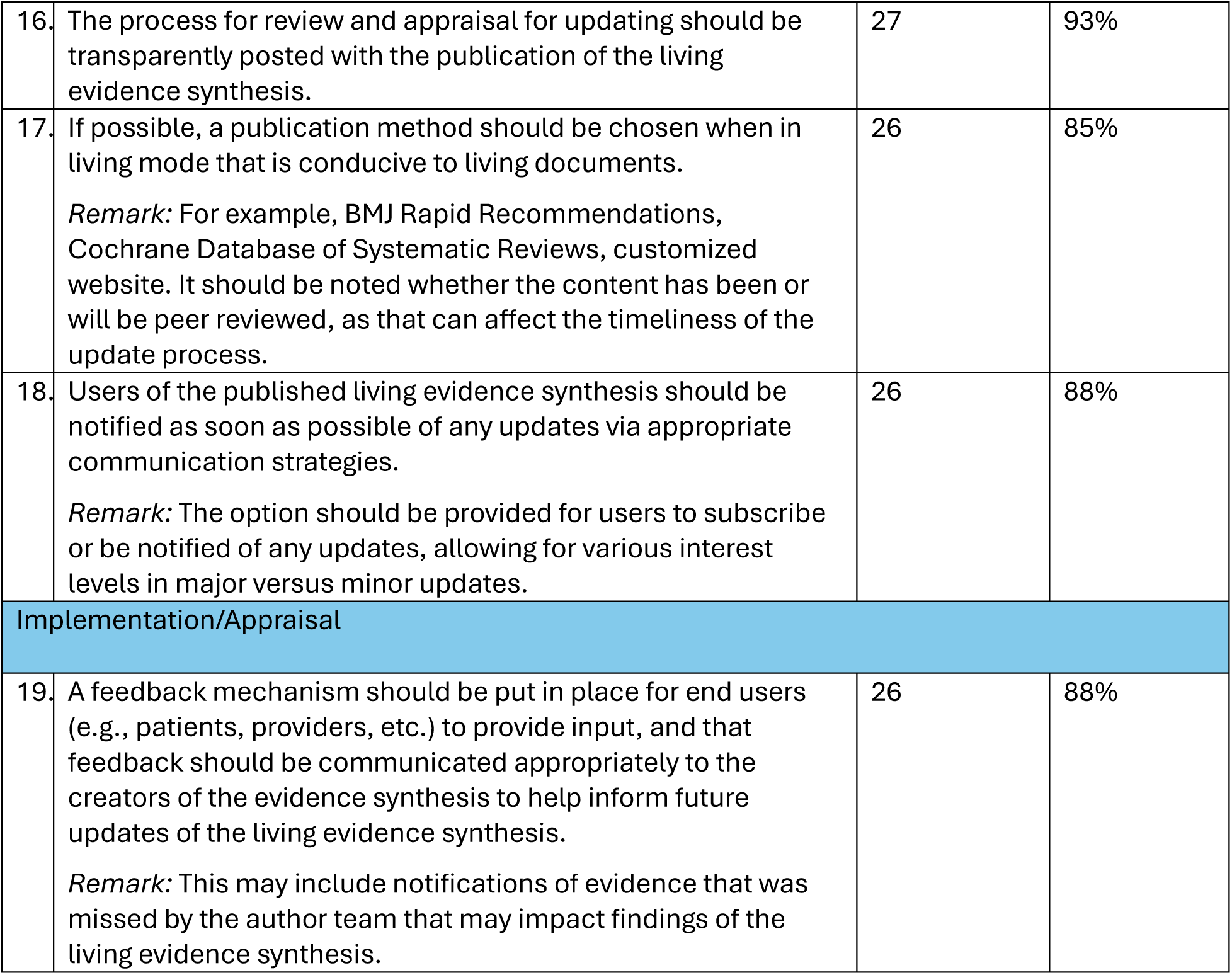
Final set of consensus-based LES best practice statements.

**Table 2.**
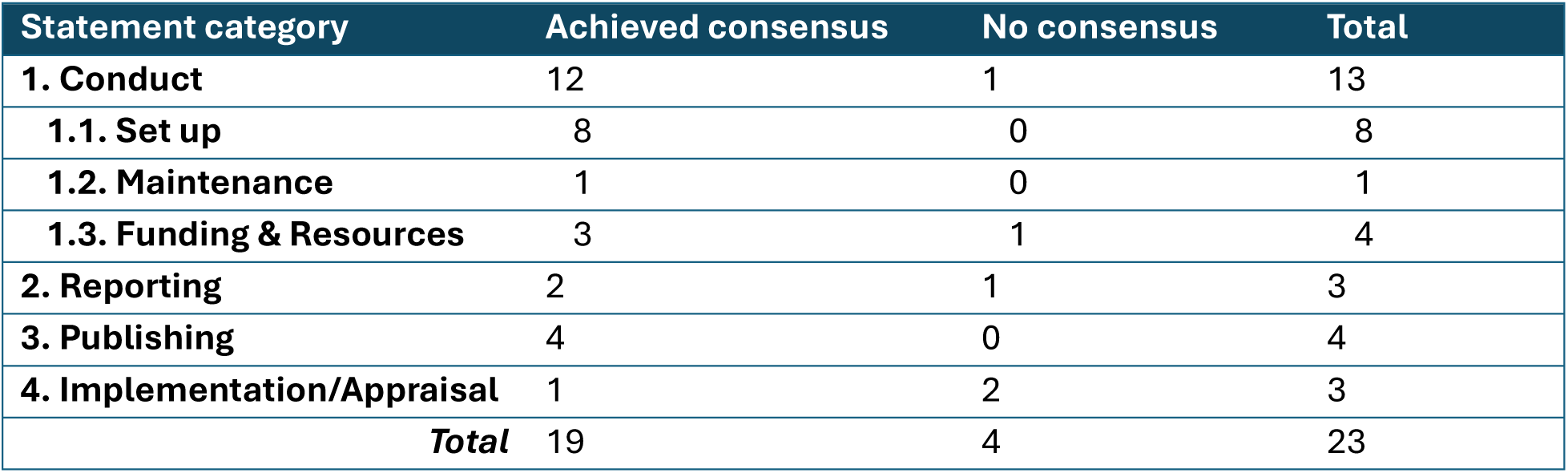
Categories of statements achieving and not achieving consensus.

| Statement category | Achieved consensus | No consensus | Total |
| --- | --- | --- | --- |
| **1. Conduct** | 12 | 1 | 13 |
| **1.1. Set up** | 8 | 0 | 8 |
| **1.2. Maintenance** | 1 | 0 | 1 |
| **1.3. Funding & Resources** | 3 | 1 | 4 |
| **2. Reporting** | 2 | 1 | 3 |
| **3. Publishing** | 4 | 0 | 4 |
| **4. Implementation/Appraisal** | 1 | 2 | 3 |
| **Total** | 19 | 4 | 23 |

### Statements that did not achieve consensus

The below statements did not reach consensus:

### Reporting

- The living evidence synthesis and downstream living evidence products should be structured in such a way as to maximize interoperability.

*Remark:* This may include adherence to Fast Healthcare Interoperability Resources (FHIR) standard, proper tagging and coding of relevant information, or other considerations as applicable. Depending on expertise of the team as well as functionality of available software and tools, this may be achieved by involving a developer or informatics specialist in the process.

(Note: this statement was originally two separate statements in rounds 1 and 2, but combined into one statement for round 3.)

### Implementation/Appraisal

- Sufficient detail and transparency should be provided in the living evidence synthesis to ensure accurate and appropriate evidence translation into evidence products used at the point of care (e.g., insurance policy, electronic health record, decision aid).

*Remark:* Though this is important in traditional evidence synthesis, it is of particular importance for the living process due to potential evolution of the evidence synthesis.

- Where available, evidence synthesis appraisal tools specific to the living mode should be used to assess the quality of living evidence syntheses.

*Remark:* Where unavailable, appraisal tools for traditional evidence syntheses may be modified as appropriate and applied to living evidence syntheses (e.g., adapting AMSTAR 2 to assess methodological quality or AGREE-II for reporting quality).

See Appendix 6 for anonymized participant feedback and Delphi summary results for each round.

## Discussion

This Delphi process among 29 global stakeholders with LES experience resulted in 19 statements for when to consider and how to conduct, publish, and implement LES. These statements are directed at those creating health-related living evidence and guidance, but are relevant to other fields as well. Consensus was lowest on objective, actionable use of automation and digital tools, and highest on subjective, general statements on maintaining LES and employing the living mode. Our statements align with and build on pre-existing literature for the production and publication of living systematic reviews, clinical practice guidelines, and health technology assessments.(7, 12, 13, 25-27) Comments provided in the Delphi surveys provided thoughtful considerations for how to plan and maintain LES, as well as future directions.

### Planning an LES

A quality LES is methodologically sound and well-conducted, following established standards (e.g., Cochrane’s LSR guidance, ALEC’s guidance for living guidelines).(7, 13) If not designed to be living *a priori*, transitioning an LES into living mode should follow all other applicable guidance; if *post facto*, a protocol should be drafted for such updates. Rapid development in the field may not be a factor for certain topics or evidence needs, in which case a responsive approach to monitoring evidence may be best, such as automating literature searches and updating topics as new evidence emerges (even if published infrequently).

Resourcing and commitment to keep LES updated should be weighed before beginning an LES, as funding may not be renewed. Collaboration with others on LES can distribute workload and mitigate risk; data-sharing agreements and memoranda of understanding may also be needed. An evidence synthesis authoring tool with language translation not only enables collaboration where otherwise language would be a barrier, but also allows for wider use in downstream implementation.

Appropriate voices should be represented, including not only the direct consumers of the LES, but also indirect consumers, such as those from low- and middle-income countries (LMIC), where LES are not produced as frequently; for example, ESIC has made equity a priority to ensure global representation. If some interest-holders (e.g., writers, participants, public commenters) will not be involved in certain updates due to the nature of the living process, this should be transparently stated, along with reasons why (e.g., time or resource constraints).

Perhaps the most important concern for LES is how to publish and maintain a living document in scientific journals; as of the writing of this research, there was no commonly accepted dissemination mechanism. Many journals that publish LES do not properly address updates, whether it be replacing the previous version or linking to the updated one. Journal publications can also be time-consuming and costly, preventing groups from publishing LES frequently (e.g., LMIC, small professional societies). It can be helpful to partner with a journal that agrees to publish updates. The ALEC COVID-19 portal was mentioned in comments as a good example for publishing LES. Alternatives could be a customized website, Open Science Framework, F1000 Research, or other post-publication peer review option where feedback and successive iterations occur simultaneously (this was also suggested at a Guidelines International Network North America webinar on guideline challenges).(31) In the interest of disseminating updates as soon as possible, it should be decided *a priori* if authors will choose to publish a preprint prior to peer review or forego peer-reviewed publications altogether. Updates should be balanced with the possibility of confusing evidence consumers with frequently changing decisions or recommendations.

### Maintaining an LES

It may be difficult to anticipate changing needs of the topic when maintaining an LES.

Unanticipated changes should be transparently reflected as soon as possible in future updates; method updates should be avoided. LES currency should be transparent: for example, if searches are run manually on a monthly basis, it should be acknowledged that studies may be published in the intervening time. This could be mitigated by displaying the results of an automated search of certain databases, alongside the date and results of the last full manual search. Version history may apply to the entire synthesis or the individual portions that were updated. For transparency (similar to traditional evidence syntheses), all versions and data should be made publicly available.

The update threshold should differentiate between a major and minor update. For example, a major update may include a material change in findings, such as a change in recommendation, baseline risk estimate, or risk stratification of patients. A minor update may include a new study that does not materially change the findings, which may be incorporated with the next major update. As such, elapsed time alone is not a useful update threshold.

As LES are designed to be updated throughout their lifecycle, a mechanism should be put in place for end users to provide feedback to LES creators. This may be as simple as an email address or contact information for where to send feedback on the publication, or a more structured comment form, depending on who is providing the feedback and what it is regarding; missed or new evidence would be prioritized over a non-actionable comment.

### Future of LES

The future of LES will likely necessitate the use of technology. Any software or automation should be validated and justified, having undergone thorough verification that it meets requirements and fulfils its intended purpose. These tools can be used for versioning, and to maintain tagging of relevant diagnosis or procedural codes as the LES evolves over time. Software should utilize health informatics standards (e.g., HL7 FHIR), which have been developed to support easier flow of information throughout a “learning health system.” For example, encoding LES in FHIR (i.e., computable evidence) could enable updates to living computable guidelines, facilitating incorporation into clinical decision support systems at the point of care and thereby enabling and refining the use of artificial intelligence (AI). ESIC has formed a working group dedicated to planning infrastructure on safe and responsible use of AI in evidence synthesis, with a focus on LES.(32)

A database management approach may involve software or automation tools, but should create consistency with a systematic way of storing, relating, and using data. This may also allow for easier incorporation of data that represents a wider range of patient populations where peer-reviewed evidence is lacking (such as those with multiple comorbidities). Data should be tagged as unpublished and not peer reviewed.

Use of an authoring tool (e.g., MAGICapp, EPPI-Reviewer, DistillerSR) can allow for the dynamic updating required for LES and may include built-in options to notify subscribers of updates. For example, the Cochrane Library allows readers to “follow” a publication to be notified of publication updates. If LES creators are not using an authoring tool or custom process where this step is automated, readers could provide an email address or other method to collect contact information to be notified of LES updates.

Another future consideration is the use of AI in LES (AI-LES). Though machine learning shows promise toward automation of certain tasks, without objective measures to test the reliability of the output of that automation, it has been recommended that human-in-the-loop systems are a better choice for the current state of large language models.(33-35) LES are a particularly promising use case for AI/automation since the baseline review can be a valuable source of training/validation data. Automation pipelines can be tested and validated against the same context (i.e., the same review) where they will then be employed.(36) The move toward AI-LES may make LES more accessible to a variety of authors and organizations, but will likely add unforeseen complications.

### Strengths & Limitations

Though statements reflect best practices, accommodations for time, resources, and staffing will likely need to be made and should be transparently reflected in LES. Broad stakeholder representation in health-related LES – academics, government, software developers – brought diverse perspectives to this research. However, this same diversity caused competing priorities and motivations that influenced responses. Likewise, fitting statements to the broader category of LES (rather than only looking at systematic reviews or guidelines) improved generalizability but complicated consensus. LES do not stand in isolation, and their value increases when considered as part of a broader and enhanced evidence ecosystem, including evaluation of impact on care as the end goal.(17, 18) This was also a limitation, increasing the challenge in identifying specific best practices, as processes vary for the different LES types. For example, the necessity of convening a guideline panel for a living guideline limits the expediency with which an update can be published, but this is not an issue for a living systematic review, which does not typically require a decision-making body to consider the evidence.

Another limitation was that feedback was not summarized and provided to the panel after each round as to why statements changed; however, this was also necessary to avoid biasing participants. The Delphi process may not have been fully understood by all participants. There were instances where participants agreed with a statement but then left comments on how it could be revised. This left it open to interpretation on what edits were considered important enough to revise that statement, if participants thought changes were needed but voted to agree it may have inflated consensus levels. Participants were from a variety of backgrounds and geographic locations, but few were from LMIC. Though this is a limitation for applicability of the consensus statements in those areas, limited staffing and resources is a consistent concern for those developing LES.

## Conclusions

This study presents the first established consensus for best practices in considering, conducting, publishing, and implementing LES in health care. These LES standards can help align global processes, improve transparency, and promote sustainability. But there is still more to be done to accomplish these objectives, requiring that groups collaborate to embrace digital tools, adopt interoperability standards, and create effective appraisal tools.

## Disclosures

MMG has performed paid consultancy work on living evidence syntheses outside of the submitted work for EBQ Consulting, LLC, and owns stock options in Dr.Evidence, an evidence-based insights software platform. DN was a member of the Royal College of General Practitioners (RCGP) steering committee to support the new Physical Activity and Lifestyle clinical priority. He has received funding for research from the NHS National Institute for Health Research School for Primary Care Research (NIHR SPCR) and the RCGP for independent research projects related to physical activity and dietary interventions. ZM is funded by an NHMRC investigator grant 1195676. GG, RH, MM, GR, JT, POV, and JLB have no conflicts or funding to disclose. The views expressed are those of the authors and do not necessarily represent those of the NHS, the NIHR, or the RCGP.

## Data Availability

All data produced in the present work are contained in the manuscript.

## Funding

This research received no specific grant from any funding agency in the public, commercial, or not-for-profit sectors.

## Acknowledgments

The authors would like to thank Delphi participants (to be named with their consent in the final version) for their contribution to these consensus statements.

## Notes

### Clinical Protocols

https://osf.io/2mt35/overview

### Author Declarations

Ethics committee of the University of Oxford gave ethical approval for this work (reference: OUDCE C1A 24 36).

### Summary of Updates

Removal of references to specific individuals in acknowledgments section until consent is obtained.

