## Appendices for "Best Practice Methods for Living Evidence Synthesis in Health Care: An International Modified Delphi Survey"

### Appendix 1. Definitions for this context.

For purposes of these consensus statements, the following definitions apply:

- **Authoring tool** – software or a platform for creating digital content.
- **Evidence synthesis** – process of systematically aggregating the health literature, e.g., systematic review, meta-analysis, scoping review, evidence map.
- **Evidence product** – tangible output of the evidence synthesis process, e.g., published systematic review, clinical practice guideline, health technology assessment.
- **Published** – any accessible version of the LES, e.g., dedicated webpage, journal PDF, or other iteration meant for public consumption of the content.
- **Readers** – any consumer of the LES content.
- **Remarks** – used to clarify or add context to the relevant statements.
- **Screen** – evaluation of studies found in the search to ascertain relevance to pre-defined inclusion criteria.
- **Search** – systematic search of available health literature.
- **Threshold** – the point at which an update is triggered.
- **Unit of update** – the smallest element that can be revised or updated independently (e.g., recommendation in a clinical practice guideline).
- **Update** – change from a previous version. This could be a status update (see Appendix 7) communicated to readers (e.g., new studies found but not incorporated into the previous version) or a published version update (see Appendix 8) that includes new information (e.g., new studies included in the analysis, new outcomes added, methods changed). Published version updates may be major when conclusions/recommendations change, or minor when new studies are added but they do not result in changes to the conclusions/recommendations.

### Appendix 2. Delphi informed consent

#### General Information

The aim of this research is to seek consensus on best-practice guidance for living evidence synthesis.

You may ask any questions before deciding to take part by contacting the researcher (details below).

The Principal Researcher is [redacted], who is attached to the Department of Continuing Education at the University of Oxford. This research is being completed under the supervision of [redacted] and [redacted].

You will be asked to complete a Delphi survey to reach consensus on living evidence synthesis. It will be up to three rounds over the course of ~6 weeks; each round will last 1-2 weeks, with the next round being sent out within 1 week after completion of the previous round. A response rate of 85% will be required. Statements achieving 80% or higher agreement will be considered as having achieved consensus. The data will form a written consensus document submitted for publication to a peer-reviewed journal. It will only be used by the researcher named above for the purposes of this study.

##### Do I have to take part?

No. Please note that participation is voluntary. If you do decide to take part, you may withdraw at any point prior to publication of the research for any reason by emailing [redacted].

We have included a 'Neutral' option if you feel you do not have enough knowledge to adequately provide input for any given statement.

##### How will my data be used?

Your responses to the Delphi survey will be anonymous.

Your IP address will not be stored. We will take all reasonable measures to ensure that data remain confidential.

The responses you provide will be stored in a password-protected electronic file on University of Oxford secure servers and may be used in academic publications and conference presentations. Research data will be stored for 5 years after publication or public release of the work of the research.

##### Who will have access to my data?

The results will be written up for a DPhil degree and published in a peer-reviewed journal.

##### Who has reviewed this research?

This research has been reviewed by, and received ethics clearance through, a subcommittee of the University of Oxford Central University Research Ethics Committee [reference OUDCE C1A 24 36].

#### Who do I contact if I have a concern or I wish to complain?

If you have a concern about any aspect of this research, please contact [redacted] or her supervisors [redacted] or [redacted] and we will do our best to answer your query. We will acknowledge your concern within 10 working days and give you an indication of how it will be dealt with.

If you remain unhappy or wish to make a formal complaint, please contact the University of Oxford Research Governance, Ethics & Assurance (RGEA) team at [redacted] or on [redacted].

Please read the statements below. If you are happy with all of the statements, please agree to continue. This will be considered to constitute giving your consent to participate in the study.

If you have any questions about the research or the statements below, please do not hesitate to reach out.

**I confirm that I have read and understand the information provided to me. I have had the opportunity to consider the information, ask questions and have had these answered satisfactorily.**

**I confirm that I am 18 years of age or over.**

**I understand that my participation is voluntary and that I am free to withdraw until 31 March 2025, without giving any reason, and without any adverse consequences or penalty.**

**I understand what will happen to my data.**

**I agree for anonymised research data collected in this study to be used in other research studies.**

**I am happy to take part in the research.**

☐ Agree

☐ Disagree

### Appendix 3. List of desired LES organizations for Delphi representation.

| Organization | Software platform(s) | Delphi representation |
|---|---|---|
| National Institute for Health and Care Excellence **(NICE)** | n/a | No |
| World Health Organization **(WHO)** | n/a | Yes |
| Cochrane | Covidence | Yes |
| McMaster University | n/a | Yes |
| Joanna Briggs Institute **(JBI)** | SUMARI (also supports Covidence) | No |
| Campbell Collaboration | (also supports Covidence) | Yes |
| Guidelines International Network **(GIN)** | n/a | Yes |
| Australian Living Evidence Collaboration **(ALEC)** | n/a | Yes |
| Agency for Healthcare Research and Quality **(AHRQ)** | n/a | Yes |
| Making GRADE the Irresistible Choice **(MAGIC)** | MAGICapp | Yes |
| US Grade Network | n/a | No |
| Epistemonikos | • Living OVerview of Evidence (L.OVE)<br>• Living Evidence to Inform Health Decisions (LE-IHD) | Yes |
| Evidence Prime | • Laser AI<br>• GradePro Guideline Development Tool (GDT) | Yes |
| Nested Knowledge | Nested Knowledge | Yes |
| Computable Publishing | Fast Evidence Interoperability Resources (FEvIR®) Platform | No |
| RobotReviewer | RobotReviewer | No |
| Alliance for Living Evidence **(Alive)** | n/a | No |
| Future Evidence Foundation | Covidence | No |
| Pitts | Pitts | Yes |
| Mayo Clinic | Living interactive evidence synthesis (LIvE) platform | Yes |

### Appendix 4. Instructions

#### How to use this Delphi

Thank you for your participation in the Delphi consensus survey to develop objective statements on living evidence synthesis. The purpose of this survey is to solicit your thoughts on the individual statements as they pertain to living evidence synthesis, as best practices for traditional evidence synthesis have been previously established. Your responses should be based on both existing evidence in the literature as well as your experience in considering, conducting, publishing, and implementing living evidence.

Please select your level of agreement or disagreement for each statement. The statements should be able to stand alone for potential re-use. Please note: there is also a ‘neutral’ option if you do not feel comfortable providing a response for a particular statement. This option should rarely be used if at all.

Whether you agree or disagree, please add comments on what you would like to see added, removed, or revised. Please provide alternate wording if applicable. This is especially important when you disagree and will help in revising the statements accordingly for the next survey round, if necessary. Please also note if there are topic areas or statements that you feel have been omitted.

***Achieving Consensus***

• A response rate of 85% is required for voting members surveyed. Panelists not responding promptly should expect to receive email reminders. NOTE: Voting will remain anonymous.

• Statements achieving 80% or higher voting 'Strongly Agree' or 'Agree' will be considered as having achieved consensus.

• Statements achieving below 80% agreement will be modified based on the comments, and re-submitted in the 2nd round of Delphi voting.

• There will be a maximum of three rounds of voting; statements not achieving consensus after three rounds will be marked as not achieving consensus.

*Contact [redacted] ([redacted]) with any questions you may have.*

**You will be asked to provide informed consent and complete a conflict of interest form (this should take about 20 minutes) before completing the Delphi survey (this should take less than 45 minutes). You are welcome to save the survey and return to it at any point.**

### Appendix 5. Conflict of interest disclosure

A conflict of interest disclosure form must be completed by all contributors to the Living Evidence Delphi Process. Please complete the form below and return to [redacted].

A conflict of interest is defined as a conflict between a person's duties and responsibilities with regard to the development of consensus statements, and that person's private, professional, business or public interests. A conflict of interest may be real, perceived, or potential.

Disclosures made through this form apply to both material and immaterial interests: financial and commercial, as well as psychological, social, and intellectual. This applies to members of the consensus panel themselves and/or their personal partners within the last 3 years.

The aim of the statement is not to eradicate competing interests, as they are almost inevitable. Participants will not be excluded from participating in the consensus statement due to competing interests.

#### Declaration of Conflict of Interest

Please complete each of the following paragraphs. In each case, if that paragraph does not apply to you please state *"not applicable"*. If the space given is insufficient, please feel free to attach a brief letter.

I. BUSINESS AFFILIATIONS

Please list below any affiliation you, any member of your immediate family (spouse, parent, or child), or anyone else residing in your household has as a director, officer, partner, shareholder, employee, consultant, agent or advisor to any person, firm or organization that might compete with or be in conflict with the interests of a consensus statement on living evidence. If none, please state.

______________________________________________________________

______________________________________________________________

______________________________________________________________

______________________________________________________________

II. GOVERNMENTAL AFFILIATIONS

Please list below any elected or appointed office or position you, any member of your immediate family (spouse, parent or child), or anyone else residing in your household hold in any branch of government or any regulatory agency having authority or jurisdiction over providers of health care, evidence synthesis, digital health, or evidence synthesis software or programs. If none, please state.

______________________________________________________________

______________________________________________________________

______________________________________________________________

______________________________________________________________

#### III. INTELLECTUAL CONFLICTS

Please list below any potential intellectual conflicts which you, any member of your immediate family (spouse, parent or child), or anyone else residing in your household may have, including: political, academic (e.g. you subscribe to a certain "school of thought"), scientific or personal interests that might lead to potential conflicts with the development of a living evidence consensus statement; membership in a medical society; membership of a professional association connected with evidence synthesis; or having a personal relationship to a representative of a company in the health care industry.

____________________________________________________________________________

____________________________________________________________________________

____________________________________________________________________________

____________________________________________________________________________

#### IV. OTHER POSSIBLE CONFLICTS

Please list below material holdings (more than 10% stock, bond or other equitable ownership and/or 5% of the net worth of an individual) which you, any member of your immediate family (spouse, parent or child), or anyone else residing in your household have in any firm or organization which might have any effect upon your independence or judgment with respect to your duties for the living evidence consensus statement.

With respect to living evidence synthesis, please briefly describe any material support (more than 5% of a budget and/or $5,000 USD) which your institution or organization receives, such as grants, fellowships, support for personnel etc. It would be helpful if you could provide an order of magnitude indication of the level of this support. For example: *“this support constitutes approximately 5 percent of the budget of the department in which I work”*, or *“this support amounts to $20,000 per annum”*.

Please list below also any affiliation that you, any member of your immediate family (spouse, parent or child) or anyone else residing in your household might have as a director, officer, partner, shareholder, employee, consultant, agent or advisor to evidence synthesis statements or guidelines, including without limitation, organizations related to health, health care data collection, scientific and research programs and clinical trials.

____________________________________________________________________________

____________________________________________________________________________

____________________________________________________________________________

____________________________________________________________________________

I hereby certify that the foregoing answers which I have furnished are true and accurate to the best of my knowledge and belief. I understand that it is my additional obligation to disclose any new actual or potential conflict of interests.

Date: ________________________________________________

Signature: ________________________________________________

Print name: ________________________________________________

### Appendix 6. Delphi summary results.

### Living Evidence Delphi - Round 1

#### How to use this Delphi

#### Informed Consent

1. I confirm that I have read and understand the information provided to me. I have had the opportunity to consider the information, ask questions and have had these answered satisfactorily.I confirm that I am 18 years of age or over.I understand that my participation is voluntary and that I am free to withdraw until 31 March 2025, without giving any reason, and without any adverse consequences or penalty.I understand what will happen to my data.I agree for anonymised research data collected in this study to be used in other research studies.I am happy to take part in the research.

Responses: 27

Agree **100%** (27)

Disagree **0%** (0)

#### Declaration of Conflict of Interest

2. BUSINESS AFFILIATIONSPlease list below any affiliation you, any member of your immediate family (spouse, parent, or child), or anyone else residing in your household has as a director, officer, partner, shareholder, employee, consultant, agent or advisor to any person, firm or organization that might compete with or be in conflict with the interests of a consensus statement on living evidence. If none, please state.

Responses: 21

none

I am employed by Cochrane, whose income is based on royalties generated by sales of the Cochrane Library.

Founder and CEO of Pitts B.V.

N/A

None.

None

HESRI, University of Adelaide GRADE Working Group

Not applicable

None

I am sole proprietor of a LLC (Limited Liability Company, State of Oregon in the USA) which consults on issues related to guidelineins in public health, including on the use of living reviews and guidelines.

none

None

None

I am an executive, shareholder, and board member in Nested Knowledge, Inc., a company that has commercialized an AI-assisted living systematic review/evidence synthesis software tool. I am a shareholder and director in Superior Medical Experts, Inc., a medical research company that (among other services) performs systematic review services, including living systematic reviews.

None

none

None

Founder of www.magicevidence.org

I'm an employee, and I'm a shareholder of, Evidence Prime, a software company that develops tools for evidence synthesis and guideline development.

I am the lead of the Living Evidence to Inform Health Decisions (LE-IHD) Program, which, among its initiatives, promotes various services to support health sector organizations and groups in incorporating LE synthesis into the development of knowledge transfer products (e.g., Living HTA, Living Guidelines, evidence summaries for policy). https://livingevidenceihd.com/community/

None.

3. GOVERNMENTAL AFFILIATIONSPlease list below any elected or appointed office or position you, any member of your immediate family (spouse, parent or child), or anyone else residing in your household hold in any branch of government or any regulatory agency having authority or jurisdiction over providers of health care, evidence synthesis, digital health, or evidence synthesis software or programs. If none, please state.

Responses: 21

I am not an elected or appointed official. I am a U.S. government employee.

None

N/A

None.

I am a member of GIN

Singapore Ministry of Health, Agency for Care Effectiveness

Not applicable

None

Centers for Disease Control and Prevention, USA: contractor 2022-2023 World Health Organization, Genève, Switzerland: Contractor 2020-present

none

None

None

None.

Agency for Healthcare Research and Quality, Evidence-based Practice Center Program. Director. The program commissions evidence synthesis.

none

> I am a staff at the US Agency for Healthcare Research and Quality, and work in the Evidence-based Practice Center Program, overseeing the production of systematic reviews.

> None

> None

> N/A

> I have no government affiliations.

> Funded by US Agency for Healthcare Research and Quality Evidence-based Practice Center Program and we develop living evidence products.

---

4. INTELLECTUAL CONFLICTSPlease list below any potential intellectual conflicts which you, any member of your immediate family (spouse, parent or child), or anyone else residing in your household may have, including: political, academic (e.g. you subscribe to a certain "school of thought"), scientific or personal interests that might lead to potential conflicts with the development of a living evidence consensus statement; membership in a medical society; membership of a professional association connected with evidence synthesis; or having a personal relationship to a representative of a company in the health care industry.

Responses: 21

> none

> I have contributed to the PRISMA extension for living evidence.

> None

> N/A

> None.

> None

> Evidence synthesis taxonomy initiative, GRADE project groups, Cochrane Evidence Synthesis Unit, Member of Society of Research Synthesis

Not applicable

None

I have authored a paper defining "living systematic review" and "living guidelines". Citation: Norris SL. Current definitions of living systematic reviews and living guidelines need to change. J Evid Based Med. 2022;15:75-76. https://doi.org/10.1111/jebm.12478 I advise WHO on the use of evidence in guidelines, including in the use of "living evidence" and "living guidelines".

none

None

None

I am a proponent of living systematic review and support the use of technology to support such practices.

Cochrane Collaboration member ALIVE (Alliance for Living Evidence), Council member

I am a senior Cochrane editor and senior author of two living systematic reviews.

None

As founder of MAGIC and Chief Scientist in my new role I am working on several large projects on living guidelines, that include living evidence synthesis

I'm listed as an inventor on two patent applications regarding use of AI in evidence synthesis

As the PI and leader of the LE-IHD Program, I have been involved in the development of a series of tools to support authors in the development of evidence synthesis, including manuals and online resources such as a framework-based tool, living evidence synthesis templates for protocols, baseline reports, and updates. I am also authoring papers to present our research in this regard.

None.

---

#### 5. OTHER POSSIBLE CONFLICTS

Responses: 20

none

> None other than Pitts B.V. customers

> N/A

> None.

> None

> Not applicable

> My role is funded by the Australian Government through Cochrane Australia to investigate the potential for living evidence to strengthen health policy. I am affiliated with the Australian Living Evidence Collaboration, whose remit is to develop living clinical guidelines, and with the WHO, who have a focus on implementing living evidence syntheses into health policy and practice.

> None

> none

> None

> I have received a CIHR grant on conducting an LSR with NMA on falls prevention interventions and to conduct a SWAR on meta-analytical LSR methods. The SWAR is currently led by my PhD student.

> I am a shareholder and director in Piraeus Medical, Inc., a medical device company.

> None

> none

> Part of my work involves guideline development, but not for evidence synthesis. Part of my work involves conducting evidence synthesis, in the process of which I assess reporting (quality) of the included reports.

> None

> N/A

> I'm about to start work on a new NIHR 'Living Evidence Synthesis Group' - being co-led by Julian Higgins from University of Bristol

> I am a methodological advisor for the WHO and HTA agencies in Spain, including the Evaluation and Planning

> Service of the Canary Islands Health Service (SESCS) and the Unidad de Evaluación de Tecnologías Sanitarias de Madrid (UETS-Madrid). Currently, the LE-IHD Program receives various forms of support, the majority of which come from research grants: - I am the PI of a project aimed at developing a capacity-building strategy for implementing the LES model among organizations and groups responsible for informing health decisions (e.g., HTA agencies, guideline development groups). The three-year grant for this project is approximately €210,000. - We were awarded an EC Innovation Grant to support the development of the LE-IHD Program, which provides €80,000 per annum for two years (2024-2025). - We were awarded an internal grant from our host research institute to support the hiring of young research staff (personnel support). This grant amounts to €36,000 per annum.

> None.

---

6. I hereby certify that the foregoing answers which I have furnished are true and accurate to the best of my knowledge and belief. I understand that it is my additional obligation to disclose any new actual or potential conflict of interests.

Responses: 27

| Response | Result |
| --- | --- |
| Agree | **100%** (27) |
| Disagree | **0%** (0) |

#### Living evidence synthesis Delphi

Definitions for this context

#### Draft consensus statements for living evidence syntheses

Conduct Standards for conducting a living evidence synthesis, including initial considerations for set up and ongoing maintenance specific to living mode.

7. A clear evidence need or question for decision making (e.g., use in a clinical policy or guideline) should be identified that necessitates employing a living mode, where no other living evidence synthesis is already available.Remark: There should be uncertainty in the evidence base, and a high likelihood that new evidence will be published on the topic.

Responses: 27

| Response | Result |
| --- | --- |
| Strongly Agree | **48%** (13) |

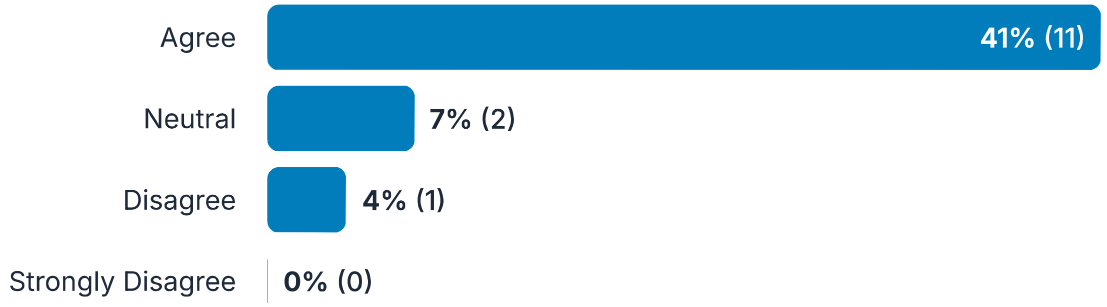

#### 8. Comments:

Responses: 7

> The primary rationale for a living evidence synthesis is the anticipated production of relevant, new evidence in the context of uncertainty with respect to the decision. The "evidence need" is important also, but you have in the "remarks" the primary reason for making a review "living". Suggest rewording for accuracy and clarity.

> Living review can be used to keep up with any research of interest; an update may provide incremental evidence or even evidence of existing and ongoing gaps. A pressing (new) need is not required for living evidence synthesis to be employed.

> While I agree that there should be uncertainty, it may be difficult to define "high likelihood" and to predict the future.

> I strongly agree with the clear need clause; less so re: no other living evidence synthesis is already available (not all evidence syntheses are equal and it can be useful to see if results are consistent across reviews)

> It is important that the topic be of importance such that a large qantity of evidence is being generated in a short timeline, necessitating the need for constant appraisal

> With 12 years of experience with living guidelines, multiple factors may necessitate living evidence and I am not sure this covers it all. This is one of the problems with defining living evidence so broadly: to include also guidelines and HTA etc.

> I would amend the 'no other living evidence synthesis is already available', as this assumes that it's available AND suitable, fit-for-purpose and relevant. If there's a hopeless LSR 'available' that shouldn't preclude others starting

#### 9. The evidence need or question should be translated into a focused PICO question, recognizing and transparently documenting the possibility that elements of the PICO may change as the living evidence synthesis evolves

Responses: 27

over time.Remark: Identify and tag relevant diagnosis and procedural codes in PICO (software may be able to assist with this)

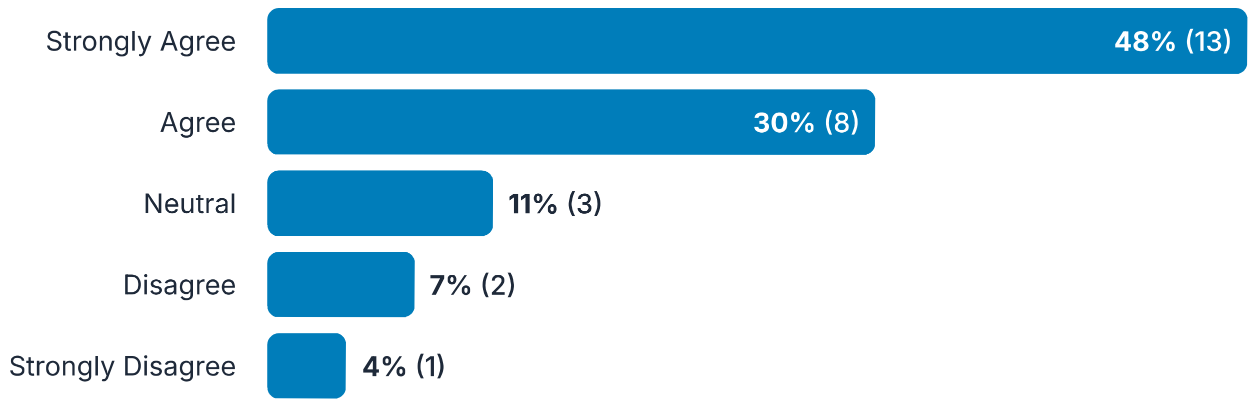

---

#### 10. Comments:

Responses: 10

> If all elements of the PICO (Population, Intervention, Comparator, and Outcome) are subject to change, it essentially creates a different review rather than a living evidence synthesis. Here's a revised perspective: Focus on a Stable PICO Framework: A living evidence synthesis should ideally focus on a review where the core PICO remains largely stable. For example, allowing for changes in the Intervention (e.g., new therapies) or the Comparator makes sense, as these areas often evolve. However, significant shifts in Population or Outcome can redefine the scope of the review entirely, potentially necessitating a new review. Practical Boundaries: Allowing only certain parts of the PICO to change ensures that the review retains coherence and relevance. For example, keeping the Population constant ensures consistency in applicability—starting with adults and later including children would introduce vastly different clinical contexts and needs. Magnitude of Change: If changes occur, their magnitude and implications should be carefully assessed. A small adjustment in intervention scope may be manageable, but broad changes (e.g., entirely new treatment paradigms or different patient groups) could compromise the integrity of the living review. Strategic Planning: Clear criteria should be defined at the outset to determine which aspects of PICO can evolve and to what extent. This ensures that the living evidence synthesis remains feasible and maintains its focus on the decision-making context.

> Appropriate of reviews on effects of interventions, but not all reviews

> The phrase "PICO question" is jargon (I think you mean a key or research question that happens to be in "PICO" format. "PICO" format only applies to quesitons about interventions - there are many other types of questions, and there are other formats (EG SPICE).. Suggest revising this statement.

> Strongly agree on all reviews for which PICOs are relevant. Note that some reviews do not have applicable PICOs (particularly preclinical reviews), but in the context of clinical reviews, this is correct.

> Would recommend flexibility such as "PICO or other question formulation framework""

> I think this is only the case for systematic reviews of interventions - is that all you are focussed on here (if so sorry for missing!). If your focus is broader (which I think it probably should be) then it seems reasonable to

> require a focussed question and inclusion criteria that stem from that, but different frameworks may be more appropriate depending on the topic - for example, PECO for reviews of exposures

> We don't have expereince with identifying and tagging diagnostic and procedural codes in PICO

> Bit limited again, for sure applies to questions of diagnosis, treatment etc but questions could relate to other EtD factors, as long as definition of living evidence is so broad

> Without a focused PICO or PICOS question, it is impossible to implement scalable rules to decide whether new evidence is relevant

> The scope of this is 'living evidence', so PICO may not be the right formulation?

---

#### 11. A threshold for updating the living evidence synthesis (e.g., elapsed time, evidence threshold, etc.) should be developed a priori and customized to the topic, as well as available and anticipated evidence.Remark: Other than a regular search for new studies, red flag warnings, FDA alerts, black box warnings, and other items may prompt the need to review and potentially update.

Responses: 27

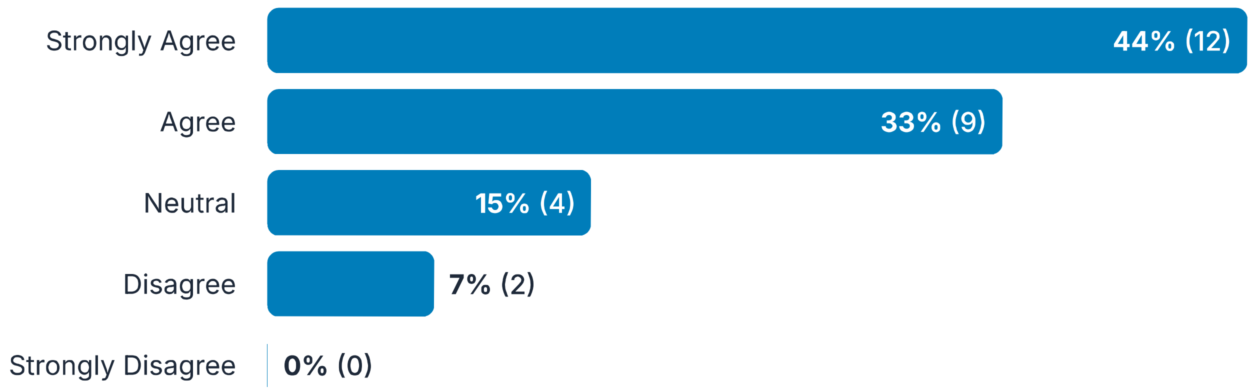

---

#### 12. Comments:

Responses: 7

> living evidence synthesis should continuously monitor for new evidence. As soon as there in enough new evidence what would lead to a change in guideline recommendations, a new living evidence synthesis should be published

> In the case of thresholds like FDA alerts or black box warnings, rapid reviews might be more appropriate unless there is a strong justification for continuous surveillance.

> Don't you usually update at a regular time interval (eg one week, 3 months), then make a decision as to whether you need to take the updated synthesis to the decision body for consideration of an updated recommendation? For the decision to take to the expert panel, you need clear, a priori decision thesholds.

> Agreed in general, best practice is to establish updating schedules or triggers in advance. However, an update on a previous review may be undertaken post facto. This is not optimal, but especially if the initial review was not proposed as 'living', this may be necessary.

> Unclear what is meant by "as well as available and anticipated evidence"

> If changes e.g. in effect estimates are negligible (below decision threshold), the updates carry the risk of changing guidance due to randomness and human factors (e.g. different composition of the panel). This can lead to erosion of trust in the guidance

> I am not certain that thresholds should be developed a priori for all situations, or that anticipated evidence should be stated a priori. In the setting of an emerging pandemic, it seems that stating these a priori may be premature.

---

##### 13. Details of how the evidence synthesis will be maintained as a living document should be transparently documented a priori in the protocol.Remark: This includes approaches to decision-making about searches and screening (sources, frequency, etc.), thresholds for incorporation/published version updates, use of software or artificial intelligence tools, plans for review of methods, and decisions about ceasing updates.

Responses: 27

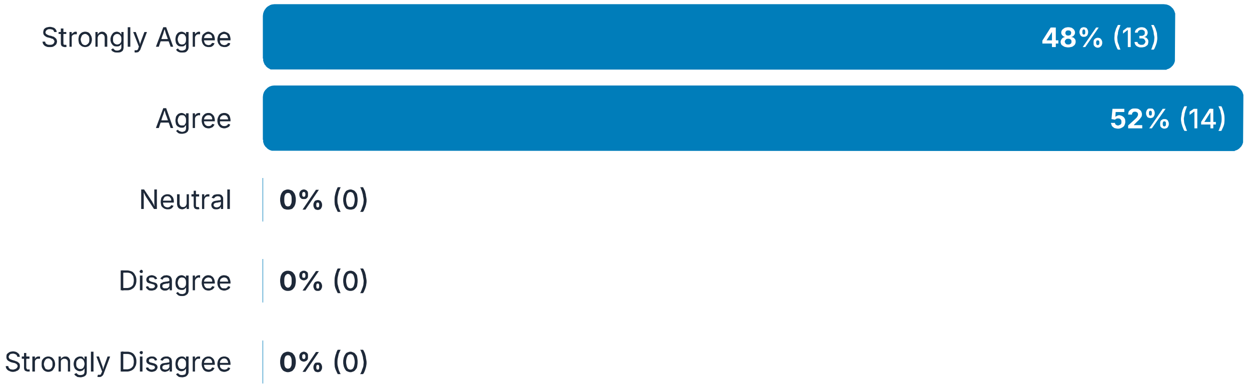

---

##### 14. Comments:

Responses: 4

> This is a very large item. Should it be split into multiple statements?

As above-- it is optimal that 'living' practices are documented a priori, but if a review is updated post facto, that optimal practice is not possible.

Should also consider thresholds that could classify an update as being 'major' or 'minor' (for example if conslusions don't change despite incorporation of a pivotal trial).

In general, it is good practice to document everything a priori. However, it should not prevent us from making the changes if necessary, or if new options (e.g. new tools) become available

---

#### 15. If resources are available, an authoring tool should be chosen that will enable updates to specific sections of the living evidence synthesis to reduce manual input.Remark: For example, if the diagnosis code changes for the relevant patient population, an appropriate databasing tool will allow changes to the code that will impact all downstream associations with that code.

Responses: 27

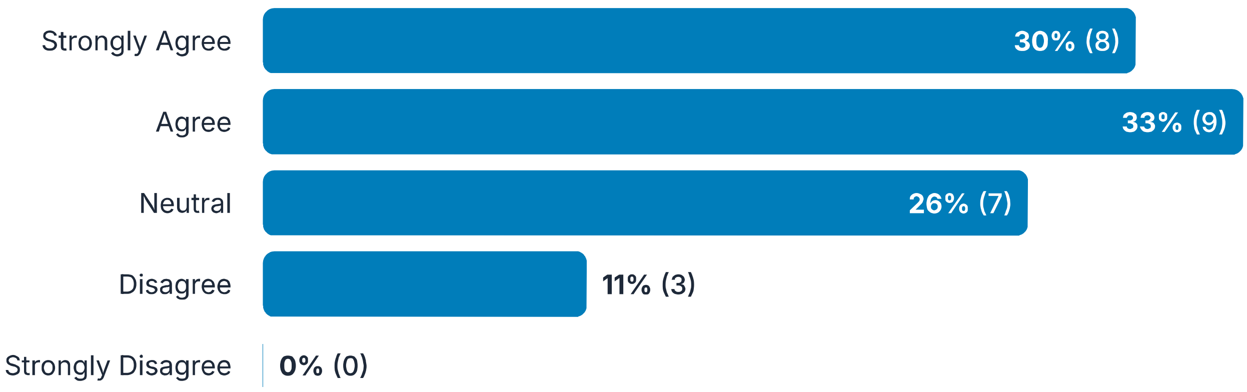

---

#### 16. Comments:

Responses: 10

I respectfully disagree and strongly suggest that an authoring tool should be used for a living review. Conducting updates manually is too risky and increases the likelihood of errors and inconsistencies.

Authoring tools that charge a fee may not be do-able for some author groups. I don't think you can mandate this.

I agree that AI will be helpful, but I would not consider this, reduce manual input, should be a requirement.

This is not in any way required, but would save work and likely promote consistency.

Should these focus on what should be done, rather than how?

> I would be worried about recommending specific software to anyone

> i theory, this sounds very appropriate, but i don't have expereince with a tool that can take clinical context into considertion.

> As responsible for MAGICapp; the only platform that is fully cabable of doing living guidelines authoring, publication and dynamic updating I believe such functionality obviously is critical. But is not only about manual input, it is about versioning, user-friendliness etc and relates to the need for digitally structured and computable data and the publication/ dissemination functionalities

> Tracking updates on the section level is a necessity. It is very difficult to achieve without tool support

> Is this referring to literature databases? This remark is unclear to me.

---

17. A plan should be developed to maintain engagement and consistency with relevant stakeholders (e.g., patient representatives, members of the public, caregivers, clinicians, providers) to inform relevant aims, objectives, and methodologies for the duration of the living evidence synthesis via public comment, panel involvement, structured feedback, or other appropriate representation.Remark: All stakeholder perspectives should be represented when feasible, including diverse viewpoints on the topic that may be relevant to a living evidence synthesis (such as for ethical or legal considerations).Remark: Consideration should also be given to how often to convene stakeholders, who to involve if not all representatives will be included each time, and when an update is needed if there are different thresholds for different update types.

Responses: 27

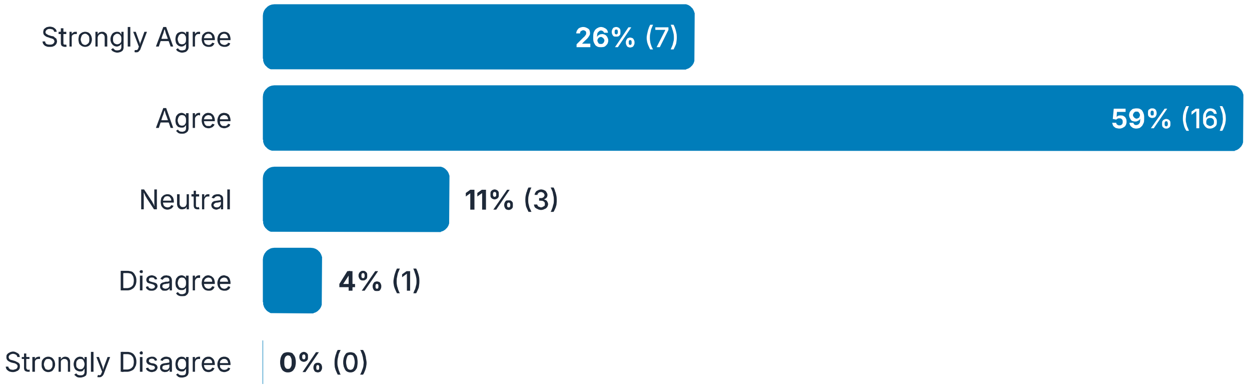

---

18. Comments:

Responses: 6

> This would be nice but it feels a bit like over-reach to mandate this.

> What is "consistency with... stakeholders" mean? I understand "engagement. To me, one of the most important inptus from stakeholders is the feasiblity of making changes to programmes when a new/revised recommendation is issued.

> Note that in many cases, published systematic reviews do not engage with relevant stakeholders. So, this should be considered an optimal practice, but with the recognition that engagement--and particularly ongoing engagement--may not be practical. Then, if the updates themselves do not have any change in aim or message, ongoing engagement would again be optimal but not absolutely necessary.

> Consider using "interest holders" rather than "stakeholders"

> Important to consider the level of resources (time and funds) available while deciding which aspects of the stakeholer engagement will be prioritized.

> Note that "interest-holders" is a proposed term to replace stakeholders. See: https://onlinelibrary.wiley.com/doi/10.1002/cesm.70007

---

19. If possible, properly vetted software and/or automation tools should be used to assist in various phases of a living evidence synthesis to facilitate ease of updating, as well as awareness for when updates are warranted.Remark: Automation and software assistance are particularly helpful for the search and screen phases.

Responses: 27

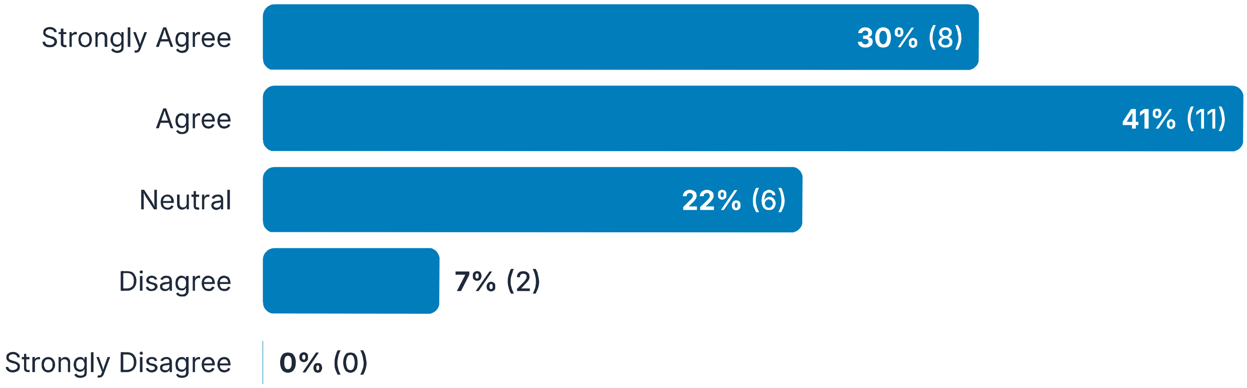

---

20. Comments:

Responses: 12

> AI means a lot of different things to different people. For this reason we need clear standards for adopting and

> using different forms of automation.

> It is helpful, but a purely manual process is not impossible. I don't believe we should exclude those that manually conduct these reviews without access to software

> This is better worded than the first mention of authoring tools above.

> What is the basis for the "remark"? need citation. in this age of rapid changes in large language models, other assistance from AI is anticipated in the near future.

> need to elaborate on what is considered "properly vetted"

> Again, I agree this would be helpful, but maybe not a requirement. An experienced librarian would be helpful to see whether the search requires any improvements at every update.

> Based on the difficulty of manual reviews and the profusion of living reviews that were begun but not carried out, easing the burden of review updates is likely necessary in almost any living review.

> "Properly vetted" could be subject to interpretation

> Again, I'm not sure automation or software tools are things that seem necessary to mandate or recommend

> There are shortcomings of current tools which hopefully the newer ones will be able to solve. For example Distiller AI needs 2000+ citations for training the AI model, and some topics may not yield such a large result from the search.

> Software equally critical for other tasks than search and screen, although I agree automation applies here. This needs to include publication as mentioned above

> Data extraction may be possible before too long, so I wouldn't put artificial limitations on the text. I would say 'validated and justified' rather than 'vetted' - which has security overtones?

#### Draft consensus statements for living evidence syntheses - Duplicate

Conduct Standards for conducting a living evidence synthesis, including initial considerations for set up and ongoing maintenance specific to living mode.

##### 21. When possible, a database management system should be used to maintain data in the living evidence synthesis to allow for ease of updating.Remark: This allows for potential incorporation of real-world data to supplement

Responses: 27

available data until peer-reviewed evidence is published, thereby increasing equity for specific patient populations where evidence is lacking. Data could be tagged as not peer reviewed.

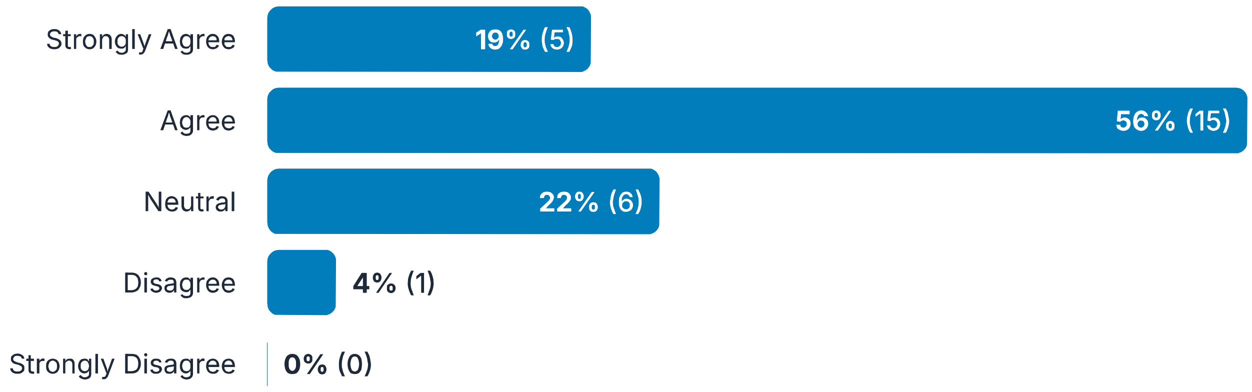

#### 22. Comments:

Responses: 7

> Unclear on this

> I guess so, but still feels a bit prescriptive

> Equity is a whole other/separate concept. Why wrap it into the database mgt system? And What is a "DB mgt system"? An effective, tailored relational database? Please define.

> While not required, one of the largest problems with living reviews is maintaining fidelity to initial data and ensuring that the updates solely add incremental studies.

> I don't quite understand how this facilitates incorporation of "real-world data" - I'd argue available but unpublished literature should be included regardless of whether a database management service is used, and there are many ways you could tag that it was not peer reviewed without using a 'database management system.' I also think 'database management system' is quite vague - would a google spreadsheet count as this?

> incorporation of RWD into a living evidence synthesis may be tricky since there are several methodologic issues that need to be considered when using RWD. Unless there is a compelling reason, it may be helpful to wait for the RWD study to be published

> Disagree with remark, does not make sense to me to include real world data and link to equity here. I would keep broader and label as evidence not yet undergone peer-review and publication and rephrase equity part

#### 23. Living evidence synthesis methods should adapt to and anticipate changing needs of the topic, the evolving evidence landscape, and other contextual changes.

Responses: 27

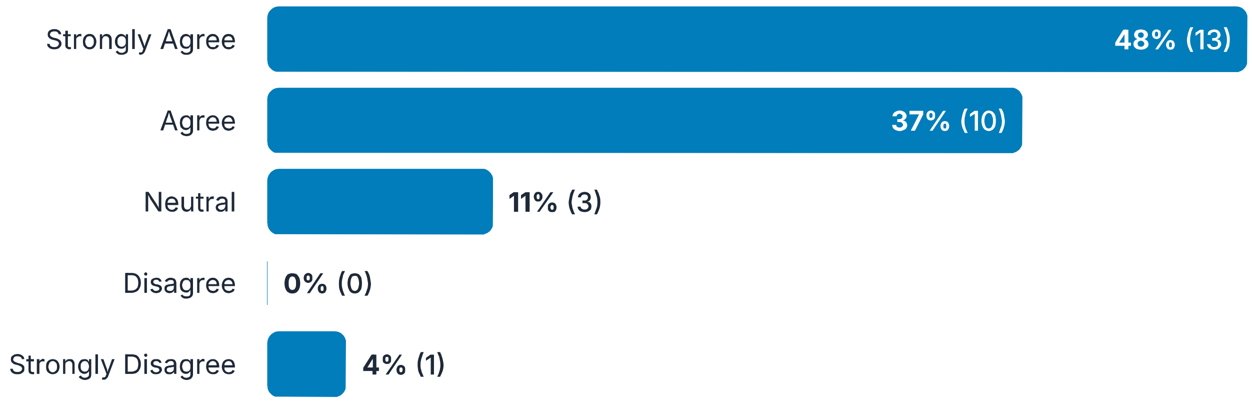

#### 24. Comments:

Responses: 7

> The methods generally should not change based on topic. However, methods and technologies do evolve, and adapting consistently across topics would be acceptable.

> Maybe you need a remark about how these changes need to be communicated to the reader

> phrases "changing needs of a topic" is vague. Please clarify.

> Such adaptations must be reported very transparently and should be due to an exogenous change, not simply due to author preference.

> It might be ideal but difficult, especially for beginner teams, to fully anticipate the changing needs, evolving landscape, and other contextual changes. Additionally, I am unsure how much teams can adapt based on this anticipation. Will the adaption be based on anticipation? If yes, then the clause could be reworded to "anticipate and adapt", and "to the extent possible" can be added to acknowledge the practical challenges. Having said that, perhaps it would be good to prompt authors to identify and state the challenges to anticipation and adaptation.

> What does the asterisk mean here?

> Yes, though then the protocol itself might need updating / rewriting, so it becomes a different project?

#### 25. As living evidence syntheses involve ongoing investments of time and resources compared to traditional evidence syntheses, efforts should be

Responses: 27

made to develop joint living evidence syntheses with relevant organizations to allow for pooling funding and resources.Remark: A carefully crafted memorandum of understanding should be mutually agreed upon and followed.Remark: A software authoring tool that allows for language translation could facilitate collaboration and harmonization.

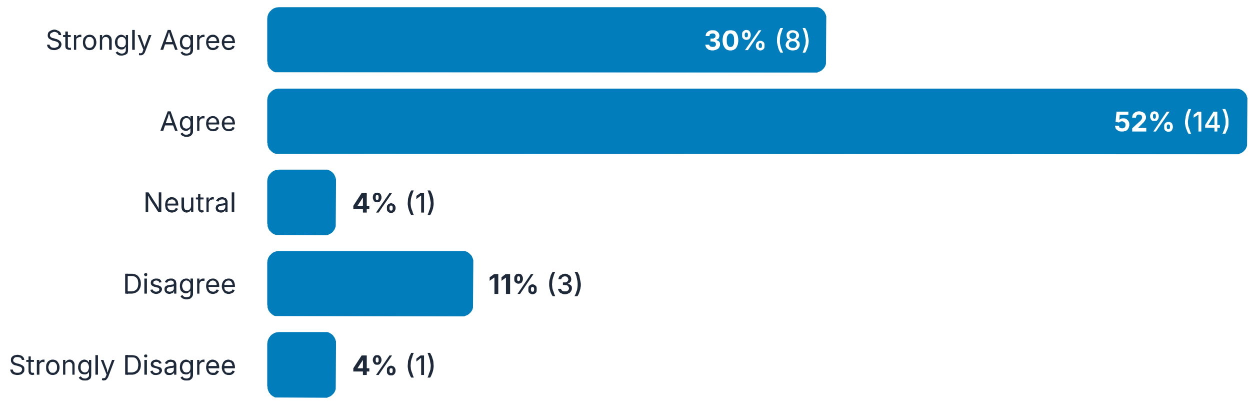

#### 26. Comments:

Responses: 8

> This approach could potentially help decrease unnecessary redundancy and conflicts in similar evidence syntheses that lead to confusing recommendations.

> I am not convinced that joint efforts with organizations will necessarily expedite the process of living evidence syntheses, as these require timeliness and speed. Collaboration may introduce additional layers of coordination and complexity, potentially slowing down the process instead of streamlining it.

> Again, this is a worthy idea but it feels that its straying too far out of methodological conduct. There is a LOT to consider when jointly developing an LES and the remarks feel rather random in terms of what they highlight.

> Would add "traditional evidence syntheses with fixed term updates. You never just do one evidence synthesis - you always plan for updates, whether fixed term (eg 3 years), or with more frequent, regular updates (ie living)

> My experience is that the more organizations involved, the longer things take, and given that the emphasis in living should include quick updating, this feels somewhat problematic. I just wouldn't say anything on this, but of course it's important the team be adequately resourced - maybe using that language would be better.

> Perhaps the wording "should be" might come across as too strong. Would "recommended" instead of "should" be appropriate? I currently get the idea that if a MoU is not followed, then I cannot proceed with living evidence syntheses.

> We will have data to support this once more living evidence synthesis are created (and the process becomes more standard), but it is likely that properly implemented living process is more cost efficient than periodic

updates.

Yes. In particular, see the ALIVE consortium for a model for this

27. A developer or code informatician should be involved or consulted throughout the living evidence synthesis process to ensure consistency and that relevant information is tagged and coded appropriately.

Responses: 27

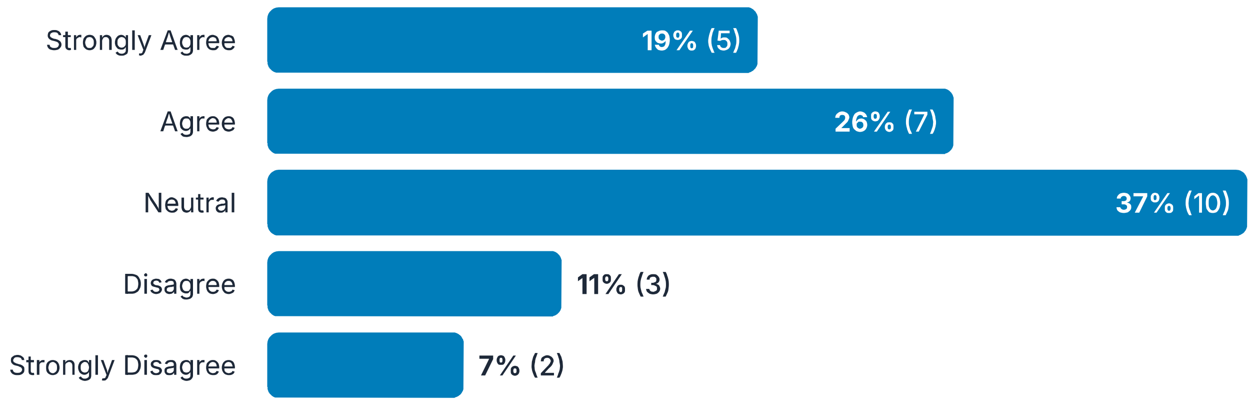

28. Comments:

Responses: 11

Depends on software used etc

Is this just as relevant for any ES? How is this specific to an LES?

I agree if these are "SMART guidelines", but your statement as written is very general and unclear.

This depends on the tools being used and the skill or knowledge of the systematic reviewers. If the tools are sufficiently transparent and usable, and the researchers sufficiently knowledgeable about them, a developer should not be required. This is a very good idea in any other case, however! s

Our LSRs do not involve this and I can't see that we'd need it, which might be me being naive, but I think more likely is because this will depend substantially on the review type and on the volume of evidence

This would be 'good to have' although not 'obligatory'.

The involvement of developer or code informatician should be planned a priori

> I think the solution is software and tools to assist in doing this. Once we put in requirement for having coders involved that further raises the bar for doing living evidence synthesis. Although MAGICapp has that coding and tagging functionality in place hardly anyone uses, even for living guidelines. Lots of collaborative work with Cochrane over 10 years has not resulted in solutions that work well. So I need to understand the rationale for this requirement

> Ideally, the authoring tools should provide enough support in this area, e.g. by merging top-down and bottom-up approaches for coding and ontology management. However, involvement of code informatician is necessary if integrating real-world data.

> Can you phrase this with less jargon? I'm not sure I know what a 'code informatician is'!

> I am unclear on what is being coded exactly?

---

29. Each version of the living evidence synthesis should include a version history, clearly detailing any updates to the status or published version and the date they were made.

Responses: 27

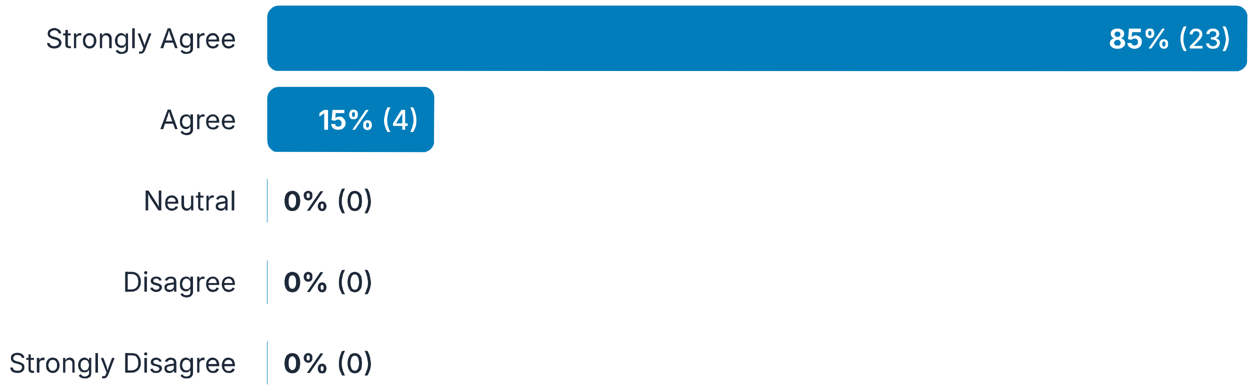

---

#### 30. Comments:

Responses: 4

> The entire living evidence synthesis could have a version, but most important is for the individual portions of the evidence synthesis that were updated to include a version number and date with details of the updates made.

> Fully agree with this. All versions should be publicly available for transparency

> Optimally, older versions of both the living review and the underlying data / database should also be available.

> This plan should be stated apriori

#### Draft consensus statements for living evidence syntheses

Reporting The reporting of systematic reviews that adopt a living methodology has been addressed by the PRISMA-LSR group; however, some elements relevant to reporting of broader evidence synthesis are included in this Delphi.

##### 31. A living systematic review should inform any downstream evidence synthesis, such as a clinical practice guideline or health technology assessment (see PRISMA-LSR for reporting standards).

Responses: 27

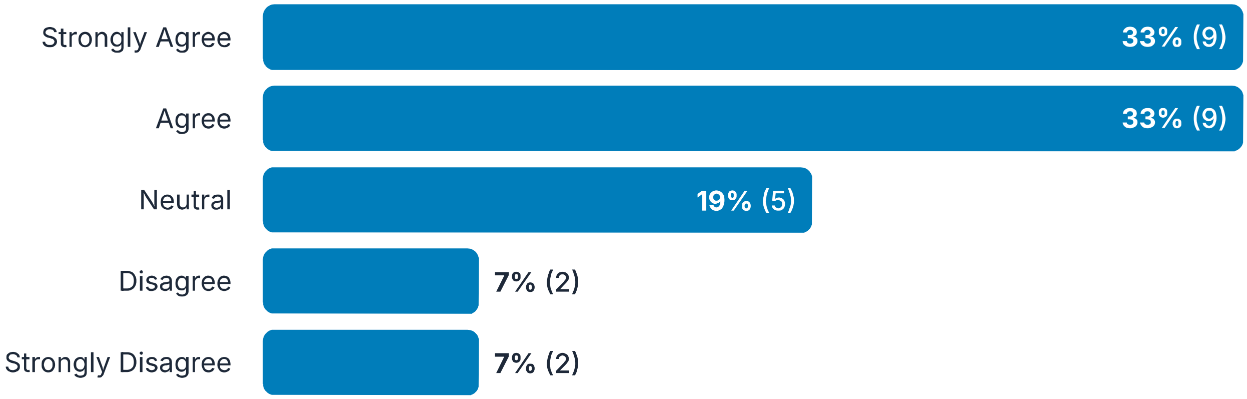

---

#### 32. Comments:

Responses: 14

> Ideally it would, but not every LSR will be done with downstream products in mind.

> I disagree that CPGs and HTA are 'downstream ES' - I would say they are ES products

> I'm not sure the PRISMA-LSR reporting standards mention is relevant here

> Yes, where there is alignment of scope, and likely future alignment of scope..

> Why link specifically to reporting standards in this statement? is another (albeit important) related, but separate issue. Yes, ideally a LSR informs decision-making, otherwise why do it (unless purely intellectual)..

> This is unclear to me. Does it imply that any downstream evidence synthesis (e.g., CPG) needs to have a LSR?

> This is unclear. "A living SR should inform a downstream evidence synthesis"--isn't a systematic review an evidence synthesis? "...such as a clinical practice guideline"--evidence synthesis is not the same thing as a clinical practice guideline

> This sounds good in principle but in practice may not work out - e.g. an evidence synthesis done by a non-member state may well not be used by the WHO for organizational reasons. This also seems like it's about use of LSRs, instead of LSR conduct itself. I would remove.

> This is imperative for impact.

> Just ensuring that authors state that the aim of the living evidence synthesis is planned to inform downstream evidence synthesis. This is vital to understand that living evidence synthesis is not an academic activity and serves a higher purpose

> Again, disagree with broad term of living evidence synthesis as specific challenges and solutions apply to systematic reviews and guidance products, respectively. PRISMA LSR is for systematic reviews and therefore different scope

> All synthesis should be done with downstream use in mind.

> Where possible, in an ideal world, sure!

> Unless I am misunderstanding, I don't think a living evidence synthesis needs to inform another evidence synthesis

---

##### 33. The living systematic review and downstream evidence synthesis should maintain diagnosis and procedure code tags as well as structured content according to Fast Healthcare Interoperability Resources (FHIR) standard (see Digital Guidelines).

Responses: 27

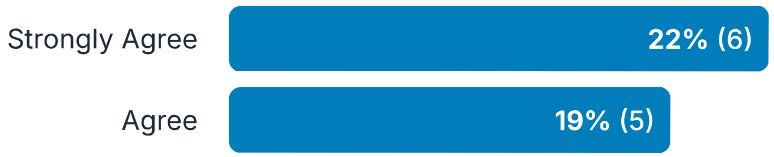

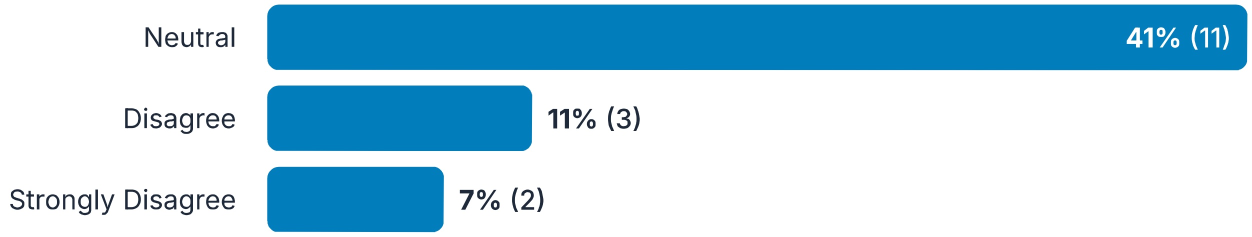

##### 34. Comments:

Responses: 9

> Not all LSRs will use diagnosis and procedure code tags. Also, this line mentioned 'downstream evidence synthesis' which is beyond the remit of this tool. Finally, isn't this relevant for all ES, not just living?

> Should link this to a prior question on informatician.

> There may be contexts where FHIR is less relevant (such as preclinical reviews), but in general, interoperability is valuable in any relevant review.

> If you are referring to ICD10 and CPT codes, I would not make this a "should" statement. This is related to the usefulness but is should not be a standard for a living SR. Also this would be applicable to living reviews on human health, but not applicable to reviews in other areas such as education, social care, or the environment.

> This seems very specific to types of review and topics and could be problematic for other topics

> Again, I do not think solutions are in place here that justifies making this a requirement although I fully endorse the concept to be developed according to FHIR

> It is a good addition, but not a requirement.

> It would be nice if there were generally recognised standards, but I'm not sure this is ready for roll-out yet, and it can be extremely difficult to incorporate

> See above

##### 35. The unit of update should be specific to the living evidence synthesis (more established for living guidelines, see Living Guidelines Framework; less so for living health technology assessments, see Living Health Technology Assessments).Remark: The entire document need not be updated each time. Relevant text sections of a systematic review, a recommendation for a

Responses: 27

guideline, or a coverage decision for a health technology assessment are examples of a unit of update.

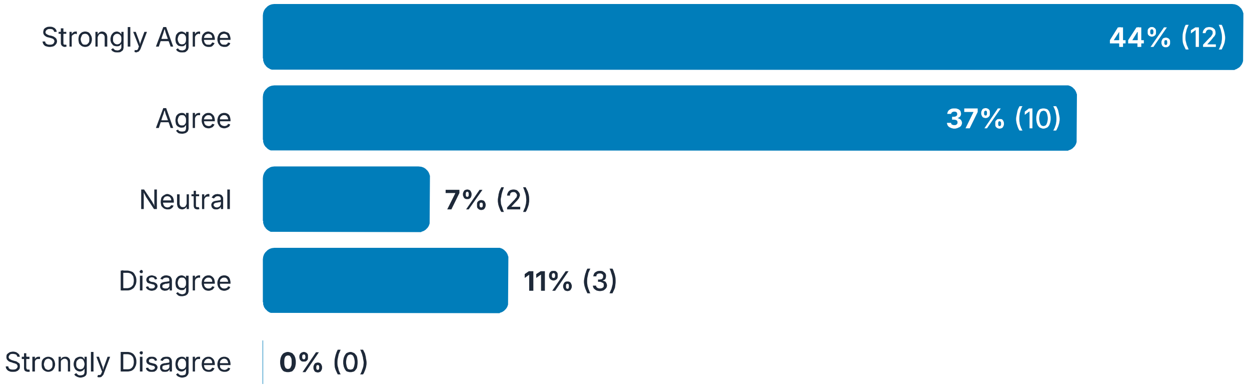

##### 36. Comments:

Responses: 4

> Nice to see this in there

> This is unclear. You may have many evidence syntheses informing a decision/recommendation. of course the "unit of update" is specific to the "living evid syn" as the latter is what you are updating. What do you mean by "document"? the review? guideline? the latter contains many recommendations, each informed by 1+ Living evid syn. Statement needs to be revised.

> Unclear. The unit of update should be less specific for living HTA?

> Unclear to me why guidelines and coverage decisions are being combined with the LES? I agree that the unit of update for LES does not need to be the entire document. But what a CPG developer does with the LES is up to them. It might not be reasonable/realistic to update any unit of the GL based on the updated LES.

##### 37. If the published version of the living evidence synthesis is not the original, a link to the previous version should be prominently displayed along with date published, and total number of versions should be stated.

Responses: 27

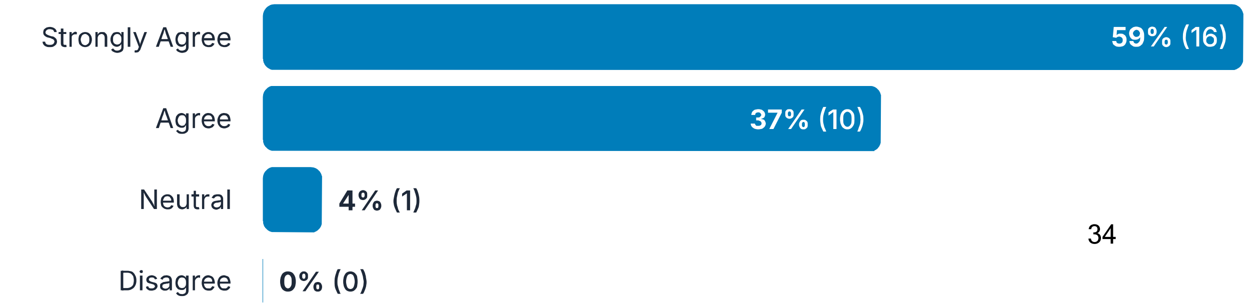

Strongly Disagree | **0%** (0)

38. Comments: Responses: 3

> This should be part of the version history

> Why is number of versions important? what is improtant is the date and changes from prior.

> This is especially important to maintain the authorship and enable citation analysis for academic credit.

#### Draft consensus statements for living evidence syntheses

Implementation/Appraisal Other considerations for ongoing use of the living model.

39. Users of the published living evidence synthesis should be notified as soon as possible of any updates via appropriate communication strategies.Remark: The option should be provided for users to subscribe or be notified of any updates. If an authoring tool is used, this may be accomplished via a push notification. Responses: 27

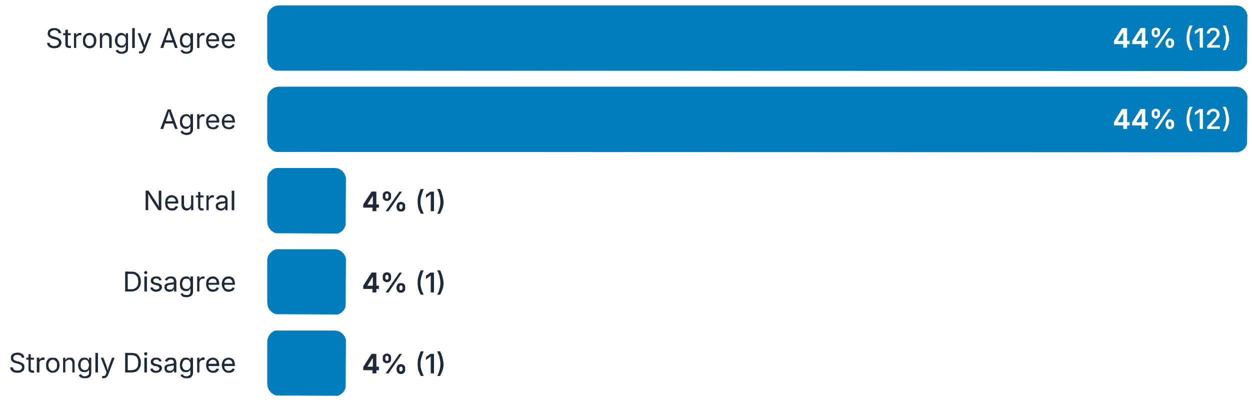

40. Comments: Responses: 5

> This feels beyond the control of the author team. It would be ideal but may not always be possible.

Ideally, users should be able to subscribe to an update level of their choice. For example, guideline developers who are using a living guideline might want to know every time the living guideline has been changed, whereas clinicians or policymakers might only want to know if a recommendation has changed (regardless of the evidence that supports it).

Public health programmes can take 2-3 years to make changes - do you really want to keep sending updates that are minor or simply re-validate existing recs?

I am finding it difficult to understand the need to specify a 'push notification' in this remark. It is meant to provide an example for this item?

I think the key point is that any website or repository that hosts living evidence needs to be customised to end-users/ target audience so allow them to go and find the most recent updates. ALEC COVID-19 website best example so far. Push notifications is easy if you have software for it (E.g. MAGICapp) but warrants people to sign up (which is also easy) but we know few people do. So not sure about the coverage one would reach. That means dissemination strategies need to be carefully developed, where portals/ websites/ platforms are key

#### 41. If incorporating into another evidence product at the point of care (e.g., insurance policy, electronic health record, decision aid, etc.), ensure appropriate translation of the living evidence synthesis in the end product.

Responses: 27

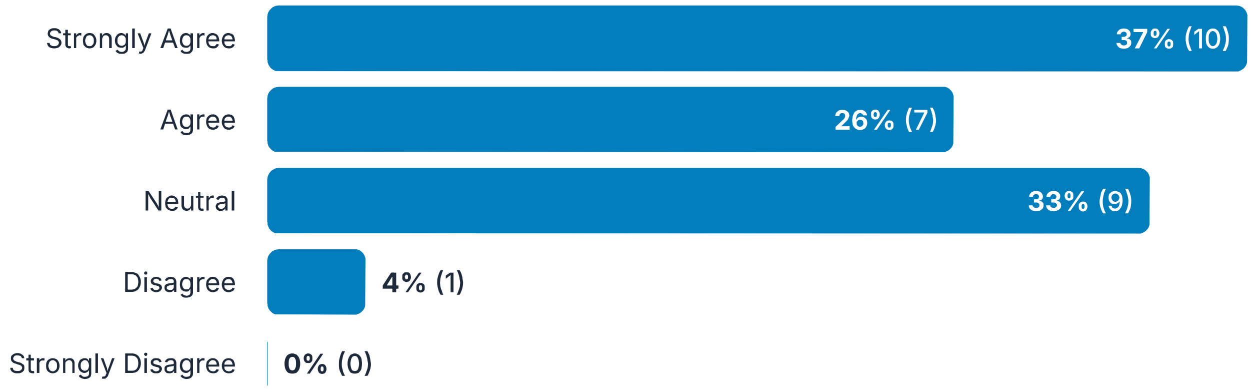

#### 42. Comments:

Responses: 9

Co-developing a standards-based (e.g., FHIR) evidence synthesis with CPGs and the end product rather than having numerous downstream interpretations should help ensure appropriate translation.

That's not in the hands of the LSR producers.

I'm not sure what is meant by 'translation'. Can you re-word this one?

This is partially the responsibility of those producing the LES and those translating it. Those who produce the LES need to ensure that there is sufficient transparency and detail to enable accurate and appropriate translation. Those who translate the evidence syntheses, have the responsibility to ensure that they are interpreting and using the LES accurately.

I don't know what "translation" means.

I don't disagree with this but it feels like it's about another end product as opposed to the LSR conduct, and I think this would be a neater project if it focussed on LSR conduct rather than their downstream use

This may be a heavy lift for the LES team. could fall into the 'nice to have' bucked rather than 'must have'.

I think this is beyond the remit of those doing the evidence synthesis

Will there be additional statements to clarify what "appropriate translation" entails?

---

##### 43. There should be a feedback mechanism put in place for end users (e.g., patients, providers, etc.) to provide input, and it is communicated appropriately to the creators of the evidence synthesis to help inform future updates of the living evidence synthesis.Remark: This may include notifications of evidence that was missed by the author team that may impact findings of the living evidence synthesis.

Responses: 27

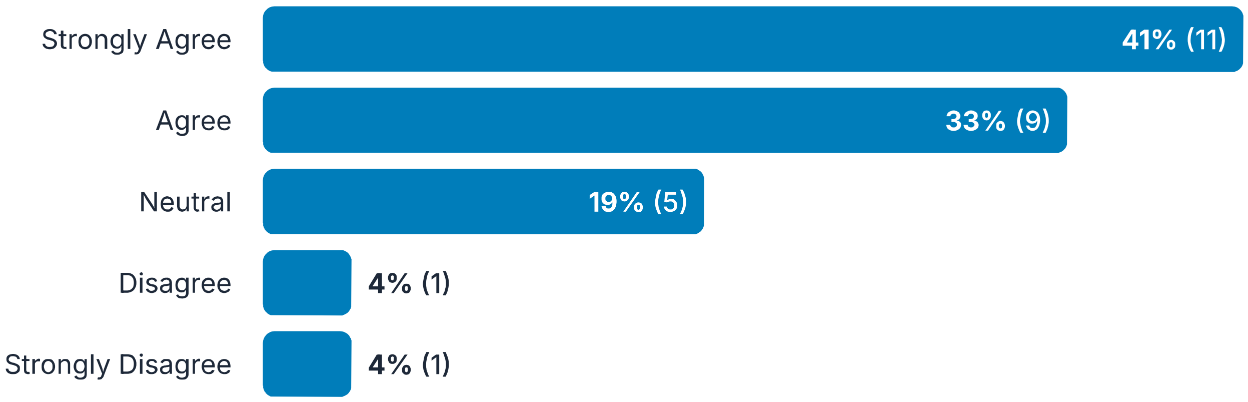

---

##### 44. Comments:

Responses: 3

Again, this seems like a good idea but is beyond agreed standards for evidence synthesis. Is it fair to expect author teams sign up for this when they do an LES?

This would be helpful and applicable to any type of evidence synthesis

Sure - though the evidence synthesis needs to be independent

##### 45. If available, living appraisal tools should be used to assess living evidence syntheses.Remark: When unavailable, appraisal tools for traditional evidence syntheses may be modified.

Responses: 27

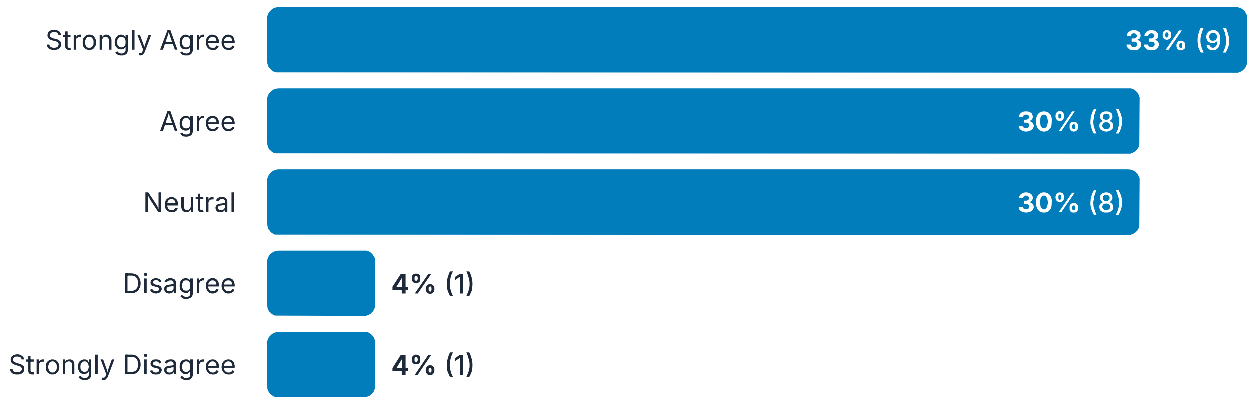

##### 46. Comments:

Responses: 4

Do we have any living appraisal tools? How far away is this?

I have no idea what 'living appraisal tools' mean - does this mean the content of the tool (e.g. signalling questions) change? If so, I would be very averse to this as it might create a lot more work version to version if you had to reassess previous articles? Or does it mean something else?

Not sure what this entails as I have not seen any such living appraisal tools

This is unclear-- who is doing the assessing? The authors of the LES? End-users? For what purpose? Is there a specific tool (or tools) you have in mind?

#### Draft consensus statements for living evidence syntheses

Publishing This section includes considerations for publishing a living document to allow for transparency as the living evidence synthesis evolves.

47. A plan should be developed to mitigate delays in publishing once the pre-defined update threshold is met, given constraints of the specific synthesis process.Remark: For example, guidelines or health technology assessments typically require input of a panel or committee to make decisions, and therefore cannot be immediately updated.Remark: For increased transparency, the evidence synthesis should display a visual threshold for when the synthesis will be updated, as well as an estimated update timeline specific to the synthesis process.

Responses: 27

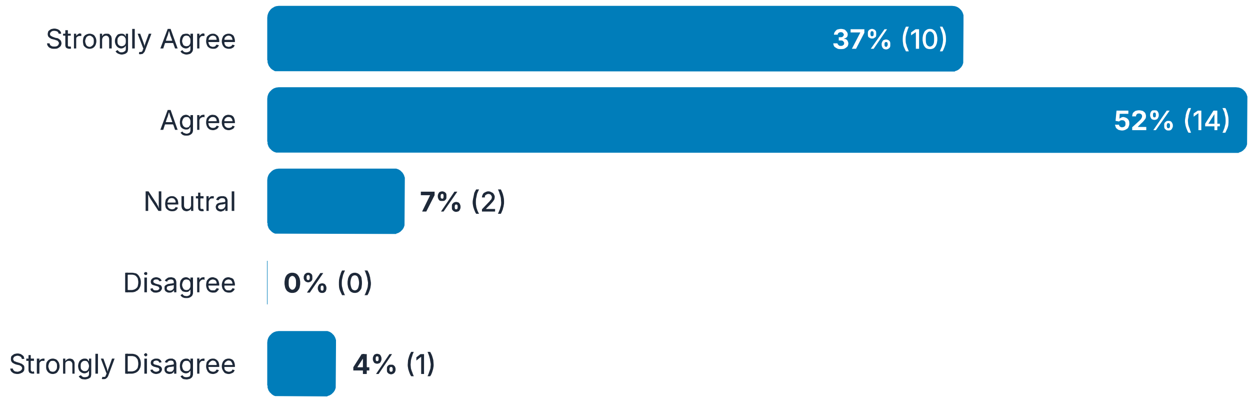

48. Comments:

Responses: 6

> This statement and the remarks could do with some re-wording for clarity.

> I might rather word this in the positive - eg "Plans should be in place to ensure that updates are published as soon as they are available..." or similar. For the first remark, could you not publish the LES when availble, and then the (udpated or revalidated) recommendation once the expert panel meets? Do you have to wait to publish the LES?

> Remark should also include mention of delays related to publication processes (journal peer review)

> This depends on teh topic. A topic like COVID has greater urgency than a newly approved drug to treat type 2 diabetes.

> Agree with the plan but do not understand the remark on visual threshold. For living guidelines we have shown that multiple factors may result in an update. For example, evidence that warrants change in strength and direction of recommendations is an easy thresholds. But lots of other scenarios I could add here

Though this can cause problems in sensitive areas

49. The process for review and appraisal for updating should be transparently posted with the publication of the living evidence synthesis.

Responses: 27

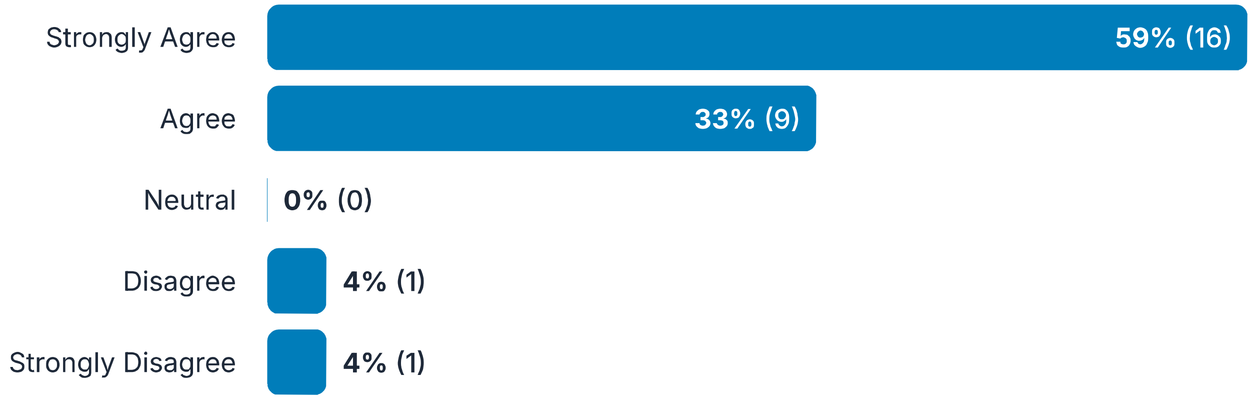

50. Comments:

Responses: 2

Maybe a living protocol could be used to be transparent as to when changes.updates are made. Dated and documented

As above, if a review is updated post facto rather than a priori, this may not be possible.

51. If possible, a publication method should be chosen that is specific to living documents.Remark: For example, BMJ Rapid Recommendations, Cochrane Systematic Reviews, customized website. It should be noted if the content has been or will be peer reviewed, as that can affect the timeliness of the update process.

Responses: 27

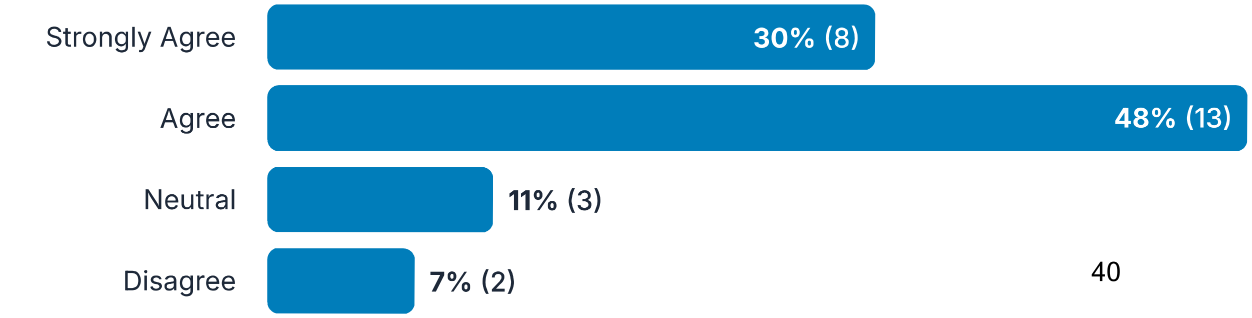

Strongly Disagree 4% (1)

#### 52. Comments:

Responses: 6

> Why does the site have to be specific? all systematic reviews should be updated when indicated - the interval just (appropriately) varies.

> Would reword--instead of "specific to living documents" rather "conducive to living documents" or something like that

> I agree with this in principle but in reality think this is hard and also am not sure that I'd say Cochrane is specific to living documents at this point!!

> it may be easier to choose a customized website over a journal may have other priorities.

> Perhaps it would be important to state when in the living evidence synthesis life cycle, the publication method may be chosen

> I think scientific publications are not at the moment well suited unfortunately. So this means repositories, customized websites are key. Just a thought

### Living Evidence Delphi - Round 2

#### How to use this Delphi

#### Living evidence synthesis Delphi

Definitions for this context

#### Draft consensus statements for living evidence syntheses

Conduct Standards for conducting a living evidence synthesis, including initial considerations for set up, ongoing maintenance, and funding/resources specific to the living mode. As with other types of evidence syntheses, living evidence syntheses should be planned in advance and clearly documented in a published protocol. Any differences from protocol should be transparently reported.

1. A living evidence approach should be considered when i. a clearly defined evidence need exists for decision making (e.g., use in a clinical policy or guideline), ii. there is uncertainty in the evidence base, iii. there is reason to expect that new evidence will be published on the topic, and iv. no other suitable living evidence synthesis is already available.

Responses: 26

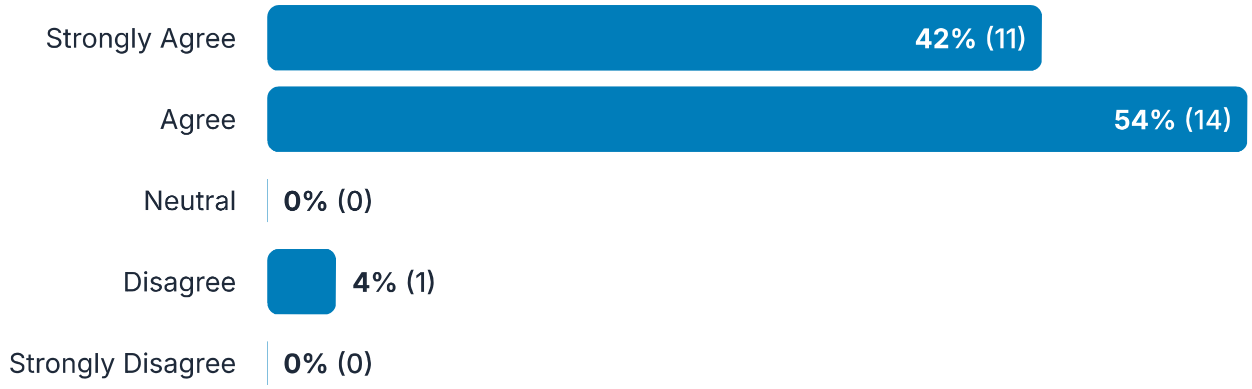

2. Comments:

Responses: 7

> perhaps expand uncertainty in evidence base to include also "rapidly developing"

> I wonder whether the example implies only for clinical, and only for policy/guideline use, whereas the definition should extend more broadly to public health including community-setting interventions and to inform practice (as policies and guidelines do not always exist in non-clinical settings eg. schools)

Need to clarify if all conditions must be met or just one

An additional component is resourcing and commitment to keep LES updated (too many stop after one or two updates). Consider adding "when the producer of the LES is adequately resourced and commits to meeting an agreed schedule/or reviewed schedule of updates.

I have come to conclude that guidance should not be included in the concept of living evidence synthesis. I would like this to be explicitly brought up and dealt with in the consensus process. I can elaborate on why but item above is one example. There are other reasons for moving to living guidelines or HTA, that go beyond the research evidence. For example, change in regulatory approval or patents for drugs and devices. I will keep on commenting from the perspective on separating evidence synthesis and guidance

I. Add (e.g. to identify information research prioritisation to address evidence gaps)

I would like to see more for iii - e.g., that there is reason to expect ongoing or emerging evidence or some indication of time factor. I don't know that a living approach is required if there might be new evidence but it may be sporadic or over many years.

---

##### 3. Thresholds for updating the living evidence synthesis (e.g., elapsed time, evidence threshold, changes in effect estimates, etc.) should be designed to instill confidence in the trustworthiness of the evidence synthesis, and be:· developed a priori,· customized to the topic and anticipated publication of new evidence,· considerate of only publishing version updates when necessary.

Responses: 26

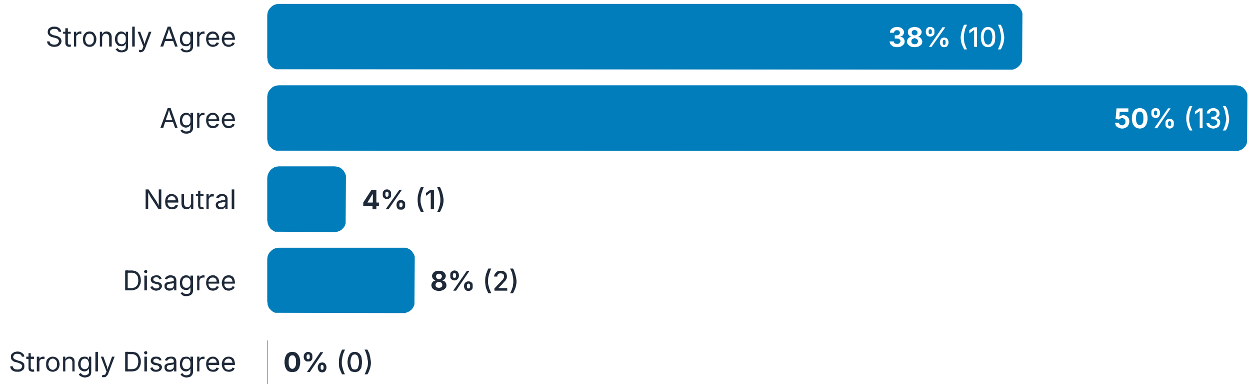

---

##### 4. Comments:

Responses: 7

Agree if, there is clear guidance on how the thresholds for updating the living evidence synthesis (e.g., elapsed time, evidence threshold, changes in effect estimates, etc.) should be customized to the topic and anticipated

> publication of new evidence. In general, in a limited recourse setting, important considerations should be regarding anticipated healthcare impact and increasing trust in the evidence and confidence that it is up-to-date.

> I think the wording for the last bullet point needs changing. I don't think 'considerate' is the right term to use. Are you wanting to say that new versions should only be published when the evidence base has changed in some way?

> Re developed a priori, I agree with this, however think it might also be good to also include mention that it needs to be periodically reviewed...i.e. over time the a priori frequency may not suit the generation of evidence and more or less frequent searches etc may be required. Re considerate of only publishing version updates when necessary: I think 'when necessary' is vague and wonder if this can be more prescriptive...when conclusions change, when new intervention approaches identified?

> Suggest re-wording the last dot point. Do you mean, 'can include publishing version updates only'? Its not clear to me.

> Interesting, I believe the remark from round 1 reflects my overarching comment: other information may prompt the need to update. This holds true for example for HTA and guidelines but maybe less so for systematic reviews?

> Add: public health emergencies, epidemics/pandemics (for communicable diseases this is when new evidence will arise)

> the last bullet point needs clarification - what is meant by "when necessary" - this could lead to selective reporting

---

5. Details of how the evidence synthesis will be maintained as a living document should be transparently documented a priori in the protocol, including:· approaches to decision-making about searches and screening (e.g., sources, frequency, etc.)· thresholds for incorporation/published version updates· use of software or artificial intelligence tools· plans for review of methods· decisions about ceasing updatesRemark: This may include a distinction of major versus minor updates.

Responses: 26

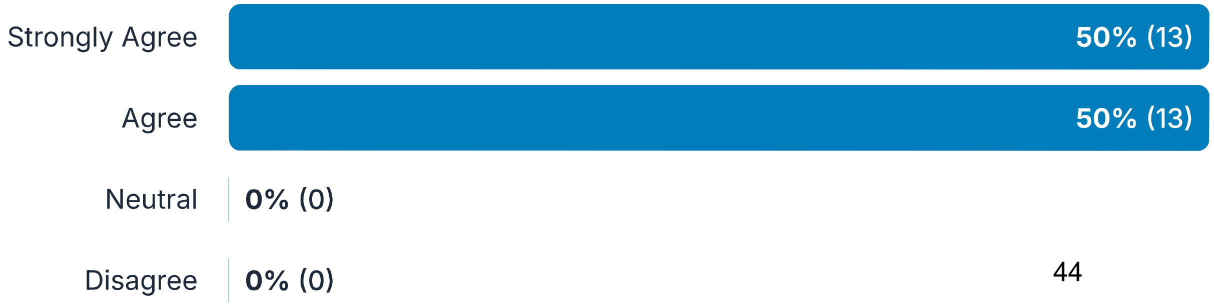

Strongly Disagree 0% (0)

#### 6. Comments:

Responses: 1

> Note that, in the case where a priori planning is not undertaken, it is sub optimal but acceptable to turn a non-living review into a living review by drafting a protocol for such updates.

#### 7. The method(s) for updating should be clearly reported and any potential for error acknowledged (e.g., manual input, specific authoring tool) .Remark: An authoring tool could enable digitally structured and computable data, if resources allow.Remark: For example, if the diagnosis code changes for the relevant patient population, an appropriate databasing tool allows for changes to the code that will update all associations with that code.

Responses: 26

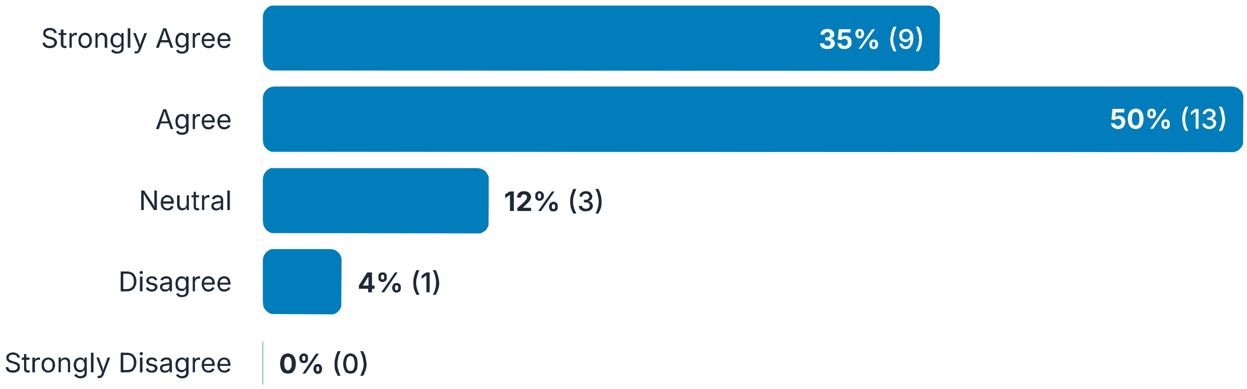

#### 8. Comments:

Responses: 5

> I guess you'd want the potential for error reported, but also the steps the team was taking to minimise the impact of errors?

> Sorry don't quite understand how the 'remark' comes into the definition?

> This is better but the opening lines 'the method(s) for updating' could be made more specific to describe that you are talking about authoring tools.

> I am hesitant when it comes to the second remark as ontologies are inherently complex to make work, as

documented in Cochrane PICO and ontology efforts. I recommend checking with experts who can confirm or refute if changes of codes can easily allow software to update all associations with that code, as stated here.

The tools available are yet limited, but new tools may become available in the future. Best not to limit it to existing tools.

---

9. The evidence need for the living evidence synthesis should be structured and appropriately-framed (e.g., PICO, SPICE, etc.) into a clear research question, transparently documenting which elements may change as it evolves over time; significant shifts may require a separate synthesis.Remark: Identify and tag relevant diagnosis and procedural codes as appropriate in the structured research question; software may be able to assist with this.

Responses: 26

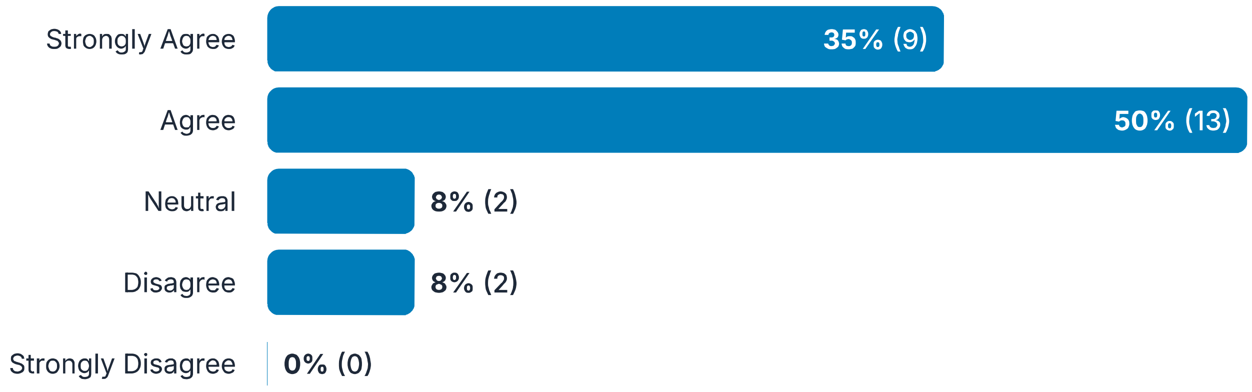

---

10. Comments:

Responses: 6

This bit in brackets - (e.g., PICO, SPICE, etc.) - should come after 'research question'

'significant shifts...' - think this might need to expand on this and have as a separate sentence.

Suggest rewording the opening lines 'the evidence need' to 'the research question'. I am also not sure about the remark. Is this specific to LSRs?

Again, this should work for systematic reviews I guess but not so well for guidelines, as basically all EtD factors can come into play. Same concern here about tagging, the use case for this remains uncertain (even though MAGIC has been allowing this for 12 years, almost no one uses it). So that remark warrants further thinking in my view

These statements seems too focused on hospital care data. To make them less restrictive and the method

> applicable to other the examples needs to be broadened.

> There are cases/topics on which a focused PICO question is not the best approach for living evidence synthesis. One example of this was the resrnt COVID 19 pandemic.

---

11. Plans should be in place to maintain engagement with relevant interest-holders (e.g., patient representatives, members of the public, caregivers, clinicians, providers) to maintain consistency in informing relevant aims, objectives, and methodologies for the duration of the living evidence synthesis (e.g., via public comment, panel involvement, structured feedback, or other appropriate representation).Remark: All interest-holder perspectives should be represented when feasible, including diverse viewpoints on the topic that may be relevant to a living evidence synthesis (such as for ethical or legal considerations).Remark: Consideration should also be given to how often to convene interest-holders, who to involve if not all representatives will be included each time, and when an update is needed if there are different thresholds for different update types.

Responses: 26

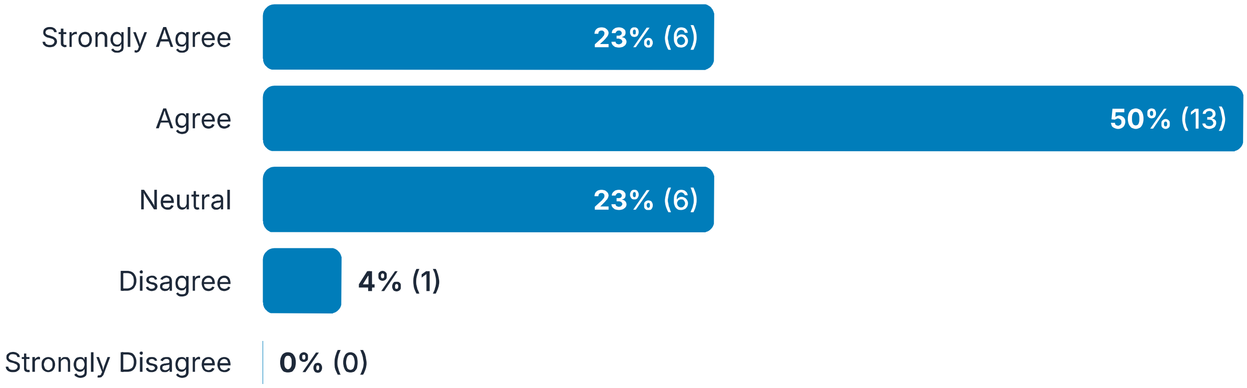

---

12. Comments:

Responses: 6

> Relevant stakeholders should also include informaticists

> Note that reviews may be undertaken by expert researchers without extensive pre-engagement of stakeholders if needed, but the plans outlined here represent optimal practice.

> I strongly support coproduction but I feel it is over-reach to put engagement as mandatory for LSRs when it is not mandatory in other evidence syntheses.

Resource constraints (especially time) may limit the input that advisory groups can have.

Agree but it is not always feasible to achieve from our experience, eg if the topic of the review is a disease eg Ebola affecting people in resource-deprived settings and during an outbreak. The need for data and the resource constraints plus need for protection during epidemics may prevent this at a time when data is urgently needed . Please add eg: whenever possible, but should not prevent a review if not feasible or appropriate.

any consideration around compensation for patient representatives, members of the public?

---

##### 13. If possible, validated and justified software and/or automation tools should be used to assist in various phases of a living evidence synthesis (e.g., update searches and screening, awareness for when updates are warranted).

Responses: 26

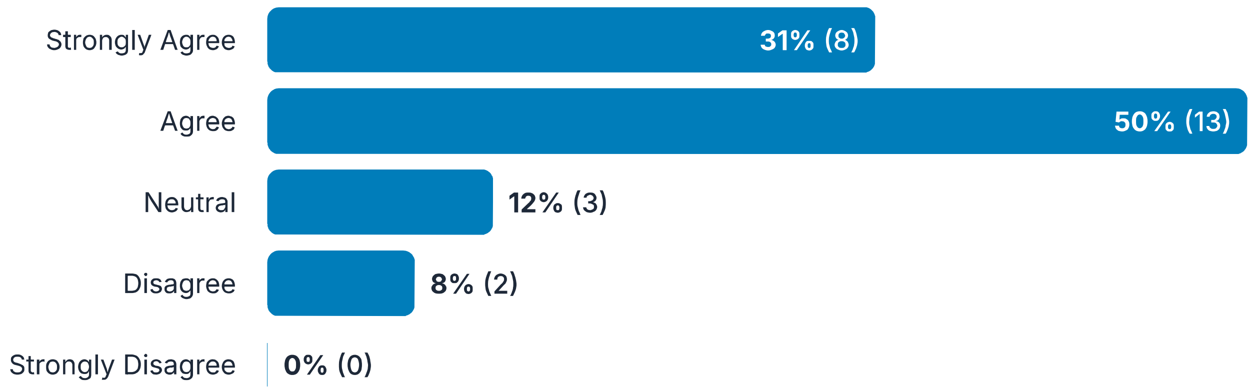

---

##### 14. Comments:

Responses: 6

Or 'Validated and justified software and/or automation tools can be used to assist in various phases of a living evidence synthesis'. I think I'm reacting to the 'if possible' and 'should', obviously we would recommend use of any validated tools for any evidence synthesis to improve efficiency and accuracy, however think for living evidence it is particularly useful

If one is going to sue such tools, they need to be validated and justified, but I don't think the use of such tools should be recommended. This question is a little unclear still. Do you mean rather, "If X,Y,Z tools are used, then they should be validated...". I would strongly agree with tat statement.

I disagree with recommending that tools should be used. I would prefer this is framed as if used, tools should be validated and justified

I believe versioning is a critical feature and rationale for software tools and should be included in the remark

> Strongly agree with all except the remark. From our experience the automation works best for programming the search, data analysis and data dissemination dashboards. Please add. (Automated screening is not robust enough fir topics that includes non-RCT study designs).

> "properly vetted" needs more definition

#### Draft consensus statements for living evidence syntheses - Duplicate

Conduct Standards for conducting a living evidence synthesis, including initial considerations for set up, ongoing maintenance, and funding/resources specific to the living mode. As with other types of evidence syntheses, living evidence syntheses should be planned in advance and clearly documented in a published protocol. Any differences from protocol should be transparently reported.

##### 15. Living evidence synthesis methods should anticipate and adapt to changing needs of the topic, the evolving evidence landscape, and other contextual changes to the extent possible.Remark: Authors should transparently communicate any known or anticipated challenges in advance in the protocol and methods when possible. For example, an anticipated challenge during the Covid pandemic was the change in variants.

Responses: 26

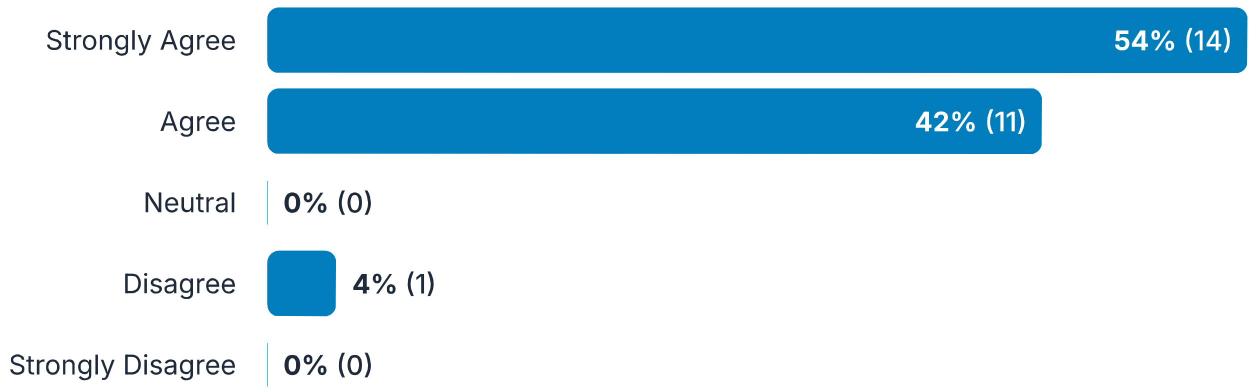

##### 16. Comments:

Responses: 5

> I guess this is OK. Its hard to anticipate some of these changes, so I would prefer the wording was altered was changed to 'expect and adapt to' but I can live with it.

> I am not sure this is a good example. At the very beginning of the COVID-19 pandemic, the emergence of different variants was not fully anticipated. While scientists knew that RNA viruses like SARS-CoV-2 could mutate, the extent and impact of these mutations—such as increased transmissibility or immune escape—became clearer only as the pandemic progressed. The need for continuous genomic surveillance and adaptation

> of public health measures became evident over time.

> It is not possible to anticipate all changing needs apriori. As such the language should be flexible to allow new consideration of elements not considered previously.

> Strongly agree, except to the note. We did not anticipate a variants in Jan. 2020 when starting our Covid response & the disease is called Covid-19. I suggest to make it disease agnostic and state: ..such as new disease strains.

> how to handle unanticipated changes?

---

#### 17. Consistency should be ensured throughout the living evidence synthesis, including proper tagging and coding of relevant information as appropriate. Depending on expertise of the team as well as functionality of available software and tools, this may be achieved by involving a developer or informatics specialist in the process.

Responses: 26

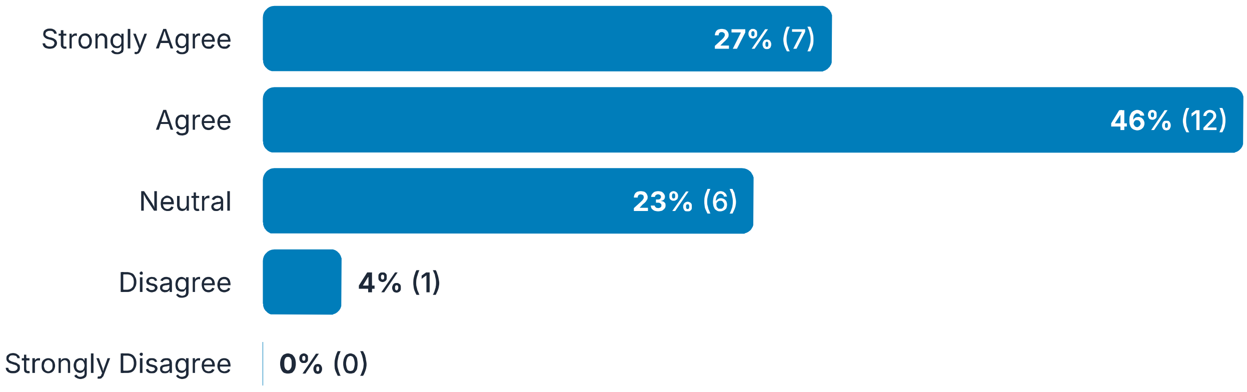

---

#### 18. Comments:

Responses: 7

> Depending on resource constraints, use of available tools may be a replacement for having an informatics specialist on the project.

> I am not certain if a web developer is required in an LSR. It is not clear to me what are the necessary tasks to be conducted/ completed in an LSR by a developer.

> Its unusual to see a focus on tagging and coding of information. Is this different to what you would recommend in a non-living review? Why is it particularly important in an LSR?

> Again, I am hesitant to put requirements to tagging and coding of reasons outlined above. What is the rationale and use-case for this?

> Suggest to edit to: .. involving a data, AI or and machine learning specialist,

> The statement is unclear. What is "relevant information" in this regard? I suggest adding an example or defining the relevance of the information for clarity.

> I'm not familiar with involving a developer or code informatician in review work - this may need more details to understand the role, training, etc.

#### Draft consensus statements for living evidence syntheses

Conduct Standards for conducting a living evidence synthesis, including initial considerations for set up, ongoing maintenance, and funding/resources specific to the living mode. As with other types of evidence syntheses, living evidence syntheses should be planned in advance and clearly documented in a published protocol. Any differences from protocol should be transparently reported.

**19. When possible, a structured, reproducible database management approach should be used to maintain data in the living evidence synthesis, ensuring transparency, organization, and ease of updating.Remark: This allows for easier incorporation of real-world data or unpublished literature (if desired) to supplement available data until peer-reviewed evidence is published, potentially representing a wider range of patient populations where peer-reviewed evidence is lacking. Data should be tagged as unpublished and not peer reviewed.**

Responses: 26

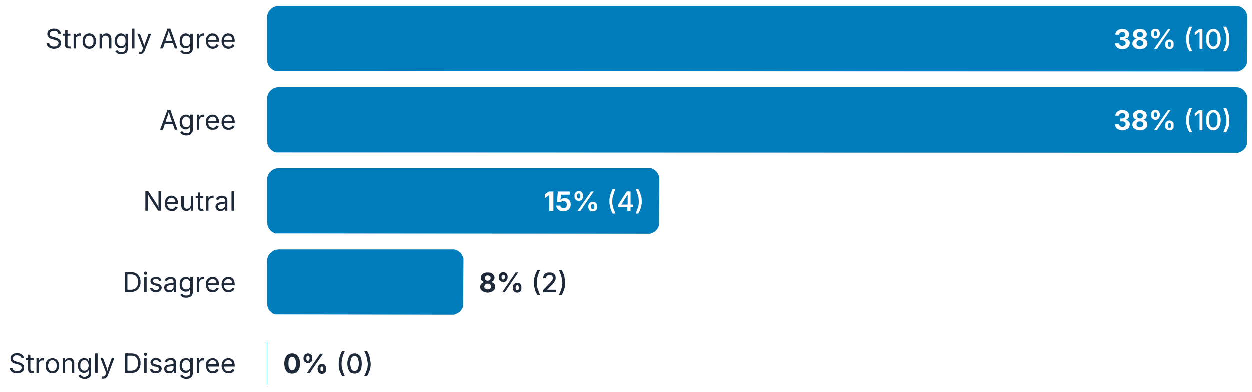

---

**20. Comments:**

Responses: 5

> I think this muddles two issues: the incorporation of real-world data, and the utility of using a good database system. Integrating real-world data is a whole programme of work and methods beyond just infrastructure to maintain good record-keeping

> Again, this is a recommendation on "how to" do a living review, which I guess fits into this exercise.

> Its not clear to me what a structured, reproducible database management approach is.

> I do not understand the emphasis on unpublished literature in the remark, in terms of how that will be applied in the living evidence synthesis. I can see other more important reasons for having a database

> I think the inclusion of unpublished or not peer reviewed data would need to be consistently applied - e.g., older unpublished studies vs. new, emerging real world evidence

---

##### 21. As living evidence syntheses are time-sensitive and necessitate ongoing investment of resources, efforts should be made to develop joint living evidence syntheses with relevant organizations to allow for pooling funding and resources.Remark: A carefully crafted memorandum of understanding between groups should be mutually agreed upon and followed.Remark: An authoring tool that allows for language translation could facilitate collaboration and harmonization.

Responses: 26

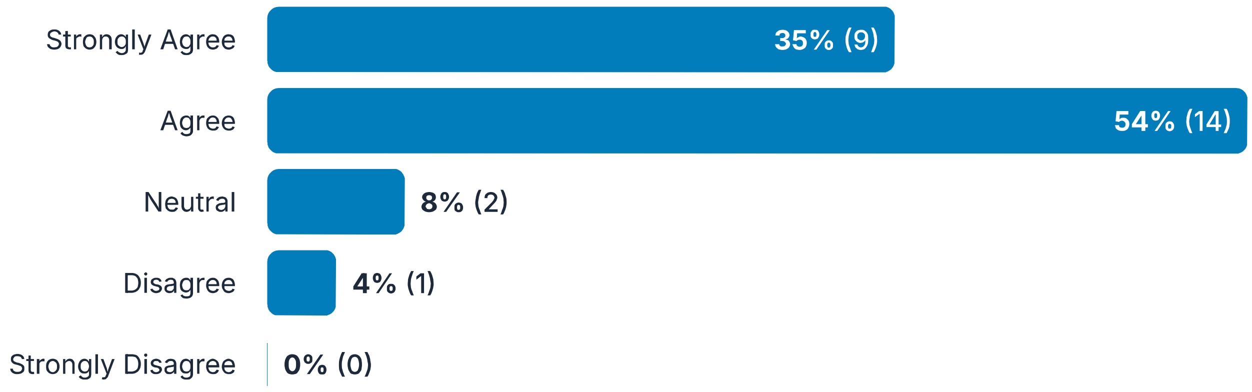

---

##### 22. Comments:

Responses: 6

> I agree if that "efforts should be made". But a living evidence project should not be dependent on the availability of funding.

> Though the 'language translation' bit seems off topic for this one. Also see ALIVE of course.

> Not sure of language 'should'...maybe it can be time and resource efficient to?

> Or the team might have the resources/expertise in house. This feels like a 'could do' not that it is something we should recommend.

> Remark: add data sharing agreement aa well as mou;

> I think the second remark about language translation should be separate from this item about investment of resources; I'm not sure what language translation applies to - inclusion of non-english studies or production/"publication" of the LES in different languages...?

#### Draft consensus statements for living evidence syntheses

Reporting The reporting of systematic reviews that adopt a living methodology has been addressed by the PRISMA-LSR group; however, some elements relevant to reporting of broader evidence synthesis are included in this Delphi. As with other types of evidence syntheses, absence of or deviation from a priori thresholds should be justified transparently.

##### 23. A quality living evidence synthesis should serve as the foundation for any downstream evidence products, such as clinical practice guidelines or health technology assessments.Remark: Some evidence products integrate the evidence synthesis into the same publication, while others publish separately.

Responses: 26

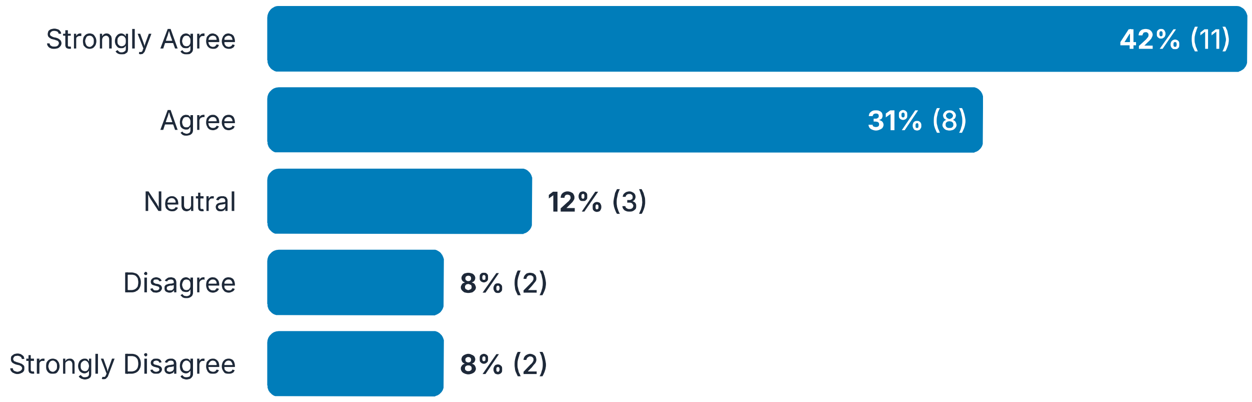

##### 24. Comments:

Responses: 8

> Yes a "foundation". But not all living evidence projects reports on findings that can directly influence clinical practice.

> Assuming that the research topic meets the 4 criteria to be considered? Or are we saying that at least for clinical practice guidelines and HTAs that any evidence foundation 'has' to have a living evidence synthesis?

> A living SR should NOT inform all CPGs or HTAs - sometimes a living approach is not needed for a CPG or HTA. And sometimes SRs are performing an an end-product in themselves.

> Where available and relevant, and where there processes are in place to update the HTA or guideline..

> I don't see an HTA as a downstream evidence product, but rather an evidence synthesis itself. HTAs are very similar to SRs, just usually with a modeling component (and also the intervention is, I believe, always a technology). HTAs can inform CPGs (as well as coverage decisions, pricing, etc) which may be US-centric (so take or leave this comment) :)

> So this one I actually agree on but chose to state disagree to reiterate the point that here you make the right distinction between living evidence synthesis and downstream guidelines or HTA. So confirms the need clarify the terminology throughout and in my view make the distinction as you have it here

> Not always feasible to set up and sustain a living evidence review. As funding is generally short term. Plus for some diseases or need not needed as new evidence may be rare.

> I think a LES "could" serve as a foundation for downstream products but might not? A given LES might not meet the needs of a given guideline panel. What is meant by "some evidence products" - provide examples?

---

25. As applicable, the living evidence synthesis and downstream evidence products should maintain diagnosis and procedure code tags as well as structured content according to Fast Healthcare Interoperability Resources (FHIR) standard.

Responses: 26

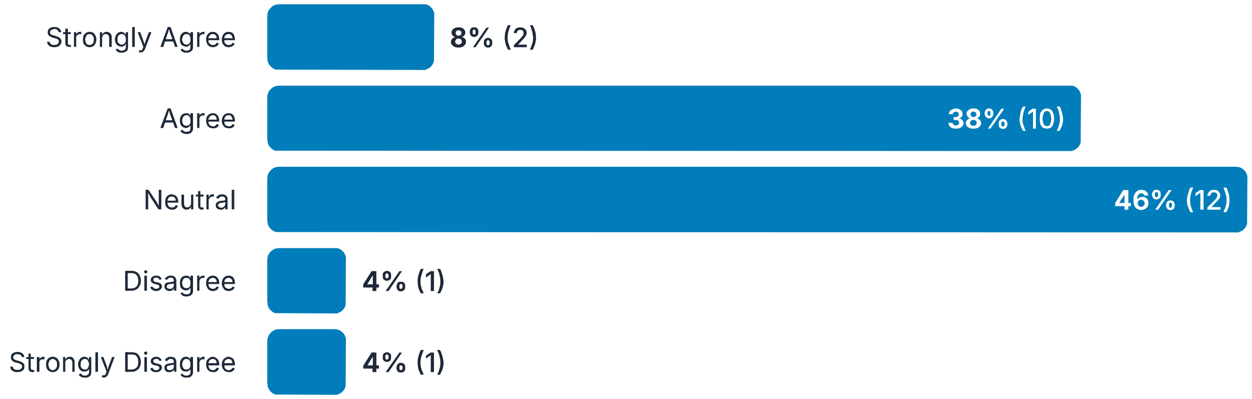

26. Comments:

Responses: 9

> This may not be relevant to all reviews, but as applicable, FHIR standards and maintenance of code tags should be maintained.

> I think this is 'nice to have', but 'should' is a bit strong. You can achieve all the objectives of a good LSR without this

> This seems very clinically focused and wonder if the language could be broadened to encompass other public health interventions? and maybe just give the diagnosis etc as an example instead? Alternatively, might need to specify that this statement only applies to clinical guidelines?

> I don't know enough about FHIR to comment on this one.

> On one hand, coding helps maintain consistency across the living synthesis lifetime (which is already mentioned in another point). At the same time, coding the results can be a separate process, potentially done by another group. Perhaps it would be helpful to elaborate on the "as applicable" part. For instance, this could depend on the downstream use or availability of health informaticians

> I'm agnostic to the FHIR standard

> As above, I am a big advocate of structured and tagged evidence and recommendations and believe FHIR is a great step forward but remain concerned about the use case here

> Change to may not always feasible or needed.

> is this assuming that the same team/group is doing the LES and the downstream evidence products? If not, how does data sharing happen?

---

#### 27. The unit of update should be specific to the living evidence synthesis, as it will be different for each type of evidence synthesis (e.g., a living guideline as opposed to a living health technology assessment); the entire document need not be updated each time.

Responses: 26

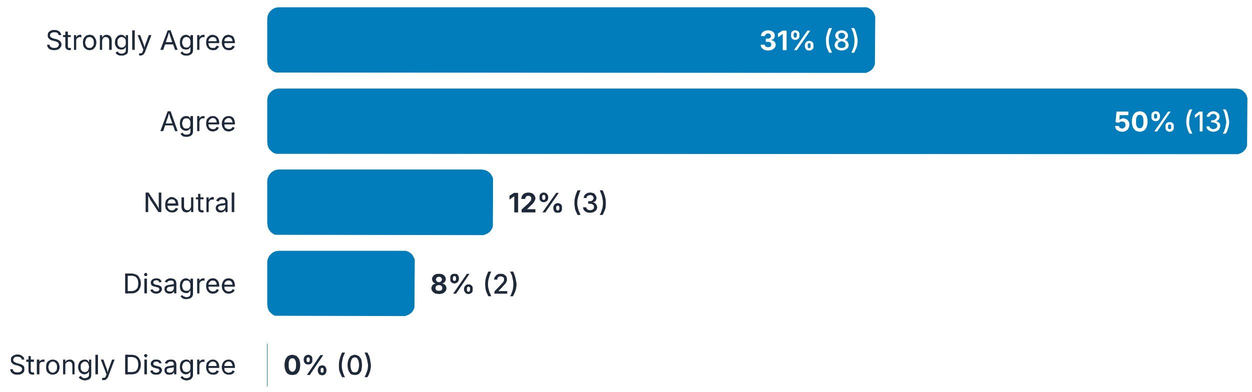

---

#### 28. Comments:

Responses: 5

> Find this one hard to read. Also wonder if at least the examples could be broadened to encompass living systematic reviews as well...i.e. update may be necessary for only one of a LSR objectives, only 1 intervention approach, or outcome

> A living guideline is not the same thing as a living evidence synthesis.

> As commented previously, this does not makes sense to me as the distinction of evidence synthesis and guidance is critical to provide better standards

> I think it will be important to provide/retain examples of units of updates

> the first part of this is confusing for me - is a living guideline and a living HTA an evidence synthesis - or is a guideline different from an evidence synthesis?

#### Draft consensus statements for living evidence syntheses

Publishing This section includes considerations for publishing a living document to allow for transparency as the living evidence synthesis evolves.

29. Plans should be in place to ensure that updates are published as soon as the pre-defined update threshold is met, given constraints of the synthesis and publication process.Remark: For example, guidelines or health technology assessments typically require input of a panel or committee to make decisions, and therefore cannot be immediately updated, but readers could be made aware of any pending updates.Remark: For increased transparency, the evidence synthesis should display a threshold (visual or otherwise) for when the synthesis will be updated, as well as an estimated update timeline specific to the synthesis process.

Responses: 26

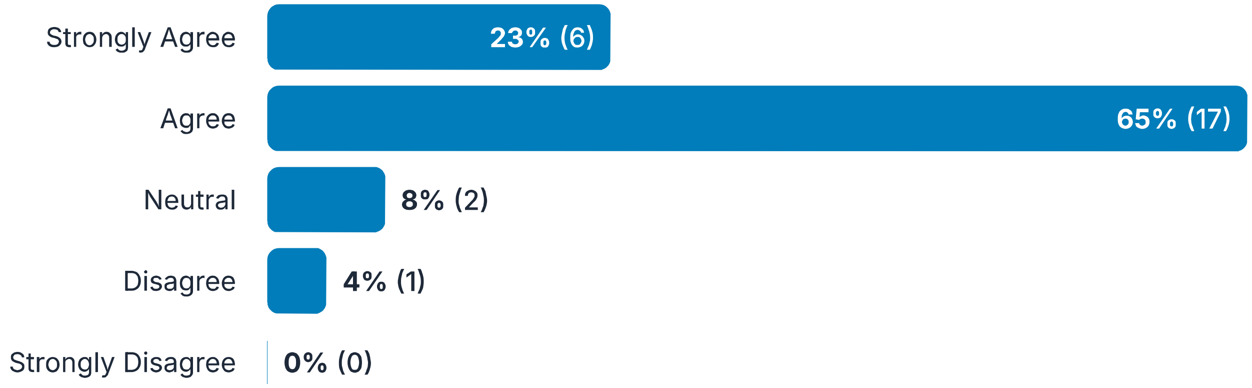

30. Comments:

Responses: 8

> Do we want to comment here on a priori agreement from journals to publish LSRs?

> What about qualitative evidence syntheses - they have no numeric threshold.

> In my experience this is very hard to do in practice and is out of authors control to some extent

> And where this is practicable.

> Same comment as before (that you can disregard)-- HTAs are evidence synthesis products, not downstream products. My understanding is they inform panel deliberations, and that panel deliberations are not part of the HTA itself. Similar to CPGs that are downstream from/underpinned by a SR.

> Same problem as above for blurring evidence synthesis and guidance. I would remove the guidance and therefore also remark 1

> Is it possible to use an alternate word for 'visual threshold'? While it is clear to me, visual thresholds have different meanings. Perhaps 'visual timeline' to be clear.

> What is meant by publishing (maybe that is defined earlier in the survey) - the reviewers themselves aren't necessarily in control of that if it needs to be peer-reviewed or published by a journal - or does this mean simply made publicly available? The first remark conflates the products - is the item about a LES? The first remark is about guidelines which may incorporate one or more LES.

---

##### 31. If possible, a publication method should be chosen when in living mode that is conducive to living documents.Remark: For example, BMJ Rapid Recommendations, Cochrane Database of Systematic Reviews, customized website. It should be noted whether the content has been or will be peer reviewed, as that can affect the timeliness of the update process.

Responses: 26

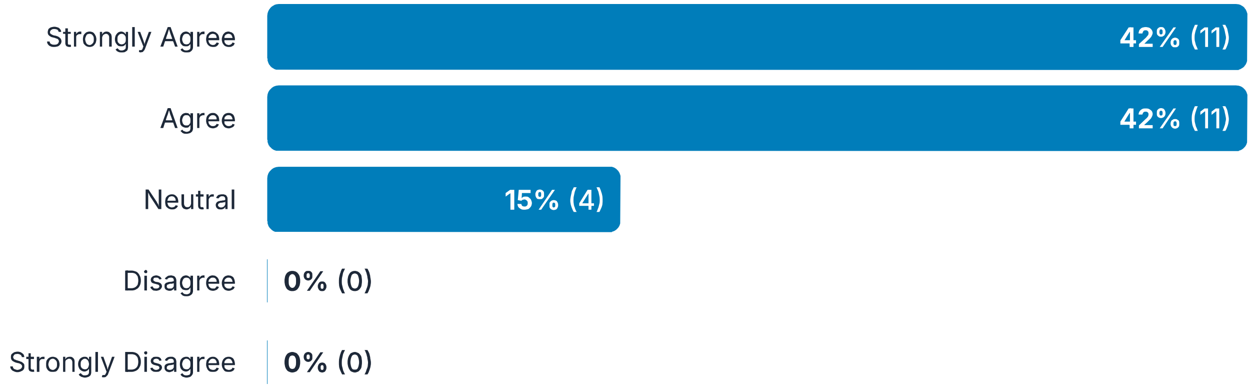

---

##### 32. Comments:

Responses: 4

> Great that you use BMJ RR as example and could have been a way to communicate living evidence but is not. So, I would rather point to ALEC COVID-19 portal, probably the best example out there of a good dissemination mechanism for living evidence. This brings me to another point: publication method is critical (knowing that nearly no journals have found a good solution) but should be part of a wider dissemination strategy that would include more aspects than publication methods. The standards should ideally cover the broader dissemination strategy in my view. Happy to elaborate

> Perhaps also consider adding-if authors choose to deposit it as a preprint, this should be decided apriori

> Not enough journals that publishes living review nor knows how to deal with updates/replace or link updates. Further the review process is slow. Disseminating data via dashboards and websites will make the data available in a more timely and equitable manner. (Journals are expensive, may prevent some LSRs to ever be done & published eg from researchers in LMICs). We need new, better & more equitable ways of disseminating data, and

peer review.

I think alternatives to BMJ and Cochrane should be considered and listed. My experience is that working through BMJ and Cochrane can be time-consuming and costly.

#### Draft consensus statements for living evidence syntheses

Implementation/Appraisal Other considerations for ongoing use of the living model.

33. Users of the published living evidence synthesis should be notified as soon as possible of any updates via appropriate communication strategies.Remark: The option should be provided for users to subscribe or be notified of any updates, allowing for various interest levels in major versus minor updates.

Responses: 26

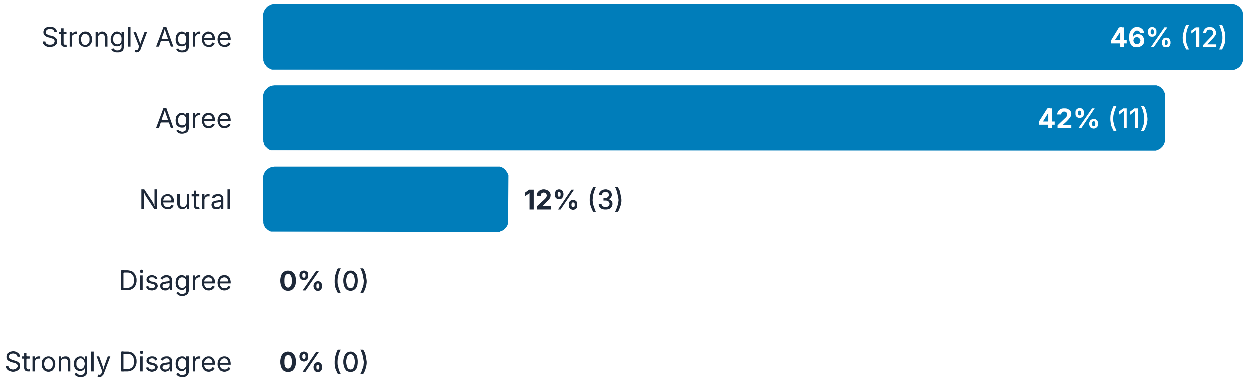

34. Comments:

Responses: 2

Rephrase to When possible,

Whose role would this be? Seems to be outside of the scope of typical reviewers. But ok if it can be automated somehow - but then need more details here I think.

35. Development of a standards-based living evidence synthesis should ensure consistency for any evidence products used at the point of care (e.g., insurance policy, electronic health record, decision aid, etc.).

Responses: 26

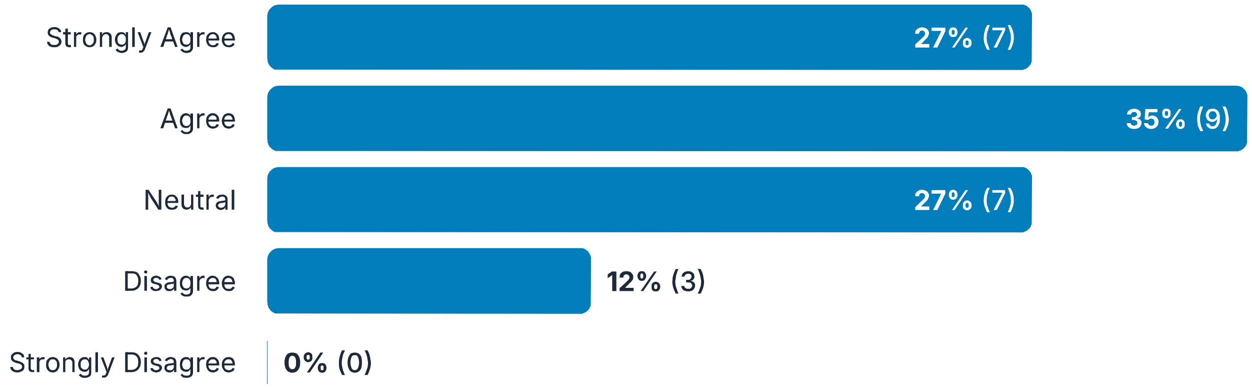

##### 36. Comments:

Responses: 8

> 'Ensure consistency' might be a bit strong. Using standards might 'facilitate' downstream use?

> Could we include a public health related example here too. eg. for community-based intvs this may be in practice guidelines or policies, or even implementation plans

> This statement is unclear to me. It may be beyond the evidence synthesis to ensure the quality of the work of those developing a derivative product (guideline, policy, etc)

> I don't know what is meant by standards-based evidence synthesis

> Can we elaborate on consistency here? Does it mean that we want to prevent situation in which different evidence products are using different versions/updates of the same synthesis?

> what is meant by 'standards-based'? What standards?

> A bit unclear, rephrase to clarify & aid interpretation.

> This is challenging and would involve much more work that reviewers typically do when producing an evidence synthesis. I think this goes beyond the LES piece which I think should be the focus of this effort.

##### 37. A feedback mechanism should be put in place for end users (e.g., patients, providers, etc.) to provide input, and that feedback should be communicated appropriately to the creators of the evidence synthesis to help inform future updates of the living evidence synthesis.Remark: This may include notifications of evidence that was missed by the author team that may impact

Responses: 26

findings of the living evidence synthesis.

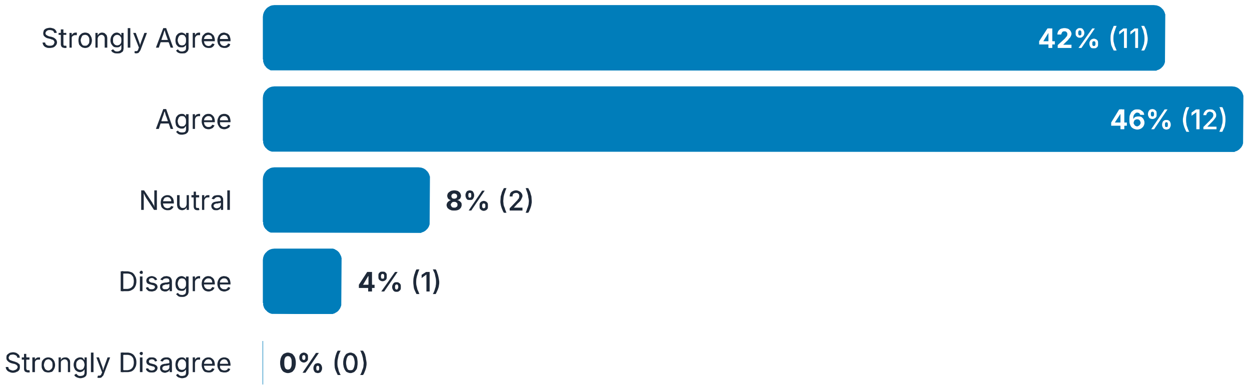

38. Comments:

Responses: 3

> Ideally but do we have the systems and processes in place to actually do this?

> as long as any evidence supplied by end-users is properly vetted by the review team for inclusion

> This seems to be applicable to NICE lsrs. Rephrase to make it applicable to all lsr's. Eg when possible..

39. Where available, evidence synthesis appraisal tools specific to the living mode should be used to assess the trustworthiness of living evidence syntheses.Remark: Where unavailable, appraisal tools for traditional evidence syntheses may be modified as appropriate and applied to living evidence syntheses.

Responses: 26

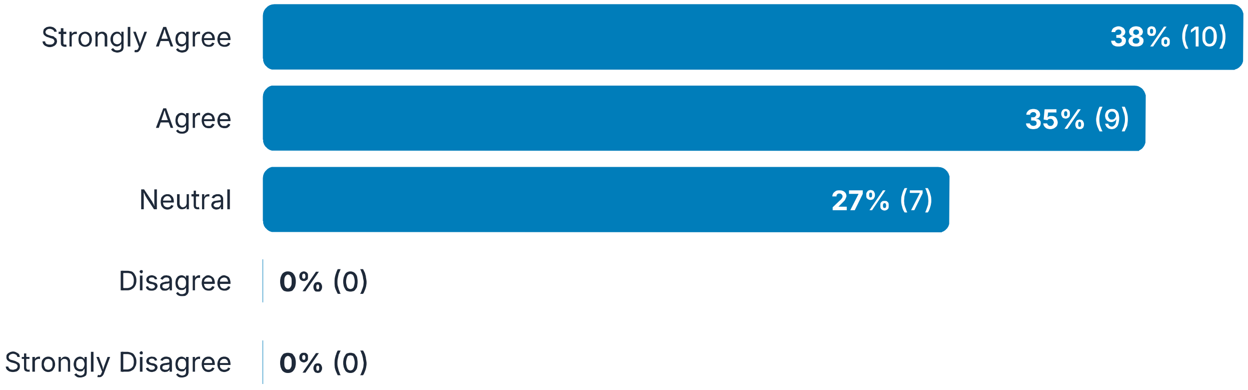

#### 40. Comments:

Responses: 4

> Would this be done by the evidence synthesis team or someone else?

> Since LES are resource-intensive but have the potential for many downstream uses, they must be usable. Therefore LES must be held to higher standards than SRs or other reviews.

> I tend to not endorse any one tool, and am not sure one exists for LES? A quality, valid tool in the wrong hands is useless.

> are there appraisal tools specific to the living mode - if so I think these should be specified otherwise this item will not be tenable; also what is meant by "trustworthiness" - current appraisal tools often focus on methodological quality or reporting quality - trustworthiness is emerging as another concept in the field of evidence synthesis but is different - I think this needs more clarity or it may lead to confusion

### Living Evidence Delphi - Round 3

#### How to use this Delphi

#### Living evidence synthesis Delphi

Definitions for this context

#### Draft consensus statements for living evidence syntheses

Conduct Standards for conducting a living evidence synthesis, including initial considerations for set up, ongoing maintenance, and funding/resources specific to the living mode. As with other types of evidence syntheses, living evidence syntheses should be planned in advance and clearly documented in a published protocol. Any differences from protocol should be transparently reported.

1. Updated statement:Thresholds for updating the living evidence synthesis (e.g., elapsed time, evidence threshold, changes in effect estimates) should be designed to instill confidence in the trustworthiness of the evidence synthesis, and be:· developed a priori,· customized to the topic and anticipated publication of new evidence,· devised to avoid publishing version updates unless findings have materially changed.Previous statement:Thresholds for updating the living evidence synthesis (e.g., elapsed time, evidence threshold, changes in effect estimates, etc.) should be designed to instill confidence in the trustworthiness of the evidence synthesis, and be:· developed a priori,· customized to the topic and anticipated publication of new evidence,· considerate of only publishing version updates when necessary.

Responses: 27

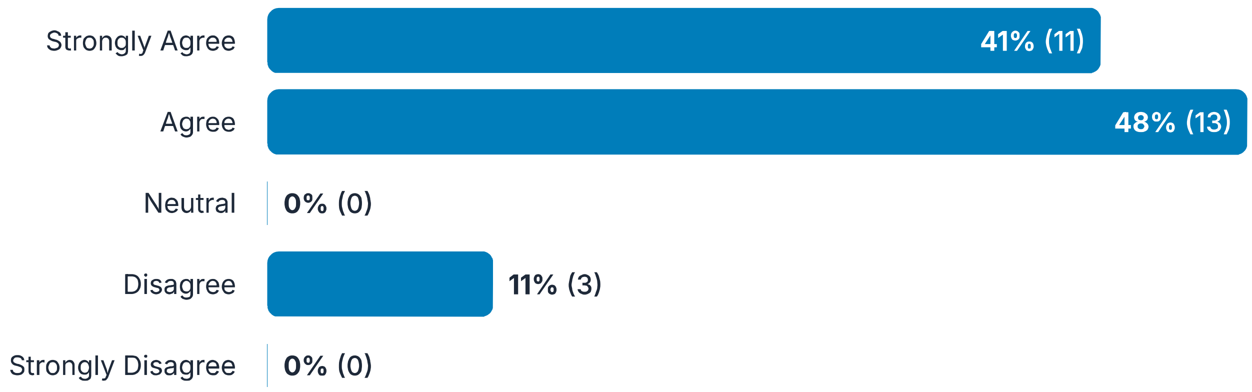

2. Comments:

Responses: 7

> As before, if not a priori, following all other guidelines is acceptable for turning a previously-non-living review into a LES review.

> Regarding the 3rd bullet point, I wonder if we would like to mention that if an updated review is conducted and results have not materially changed, then it should be mentioned in the living mode of the review (in the relevant journal or repository) where it is published, that results are up to date with updated review on the relevant date.

> I think "devised to avoid publishing version updates unless findings have materially changed" needs re-phrasing to ensure that the currency of the evidence synthesis is clear to the reader. i.e. while an entire version update might not be warranted, the reader needs to know that, e.g., the searches were updated a month ago etc.

> like this part unless findings have materially changed

> I do not agree with the third bullet point. The maintenance of a living review as current as possible is desirable, specially in web format. According to the definitions provided at the beggining, publishing is not limited to journal publication. So, I do not agree with this statement if it applies to living web-based versions, which should be updated as frequently as possible.

> Saying that somethign should be designed to instill confidence is totally vague and unhelpful. The rest is almost as non-specific.

> While I agree with the third point, I still think that there should be a note that findings have not materially changed in the evet that a threshold (e.g., elapsed time) has been met and no new (or influential) evidence was found

---

3. Updated statement:Methods for updating should be clearly reported and include any potential for error (e.g., manual input, specific authoring tool), along with strategies for mitigation.Previous statement:The method(s) for updating should be clearly reported and any potential for error acknowledged (e.g., manual input, specific authoring tool) .Remark: An authoring tool could enable digitally structured and computable data, if resources allow.Remark: For example, if the diagnosis code changes for the relevant patient population, an appropriate databasing tool allows for changes to the code that will update all associations with that code.

Responses: 27

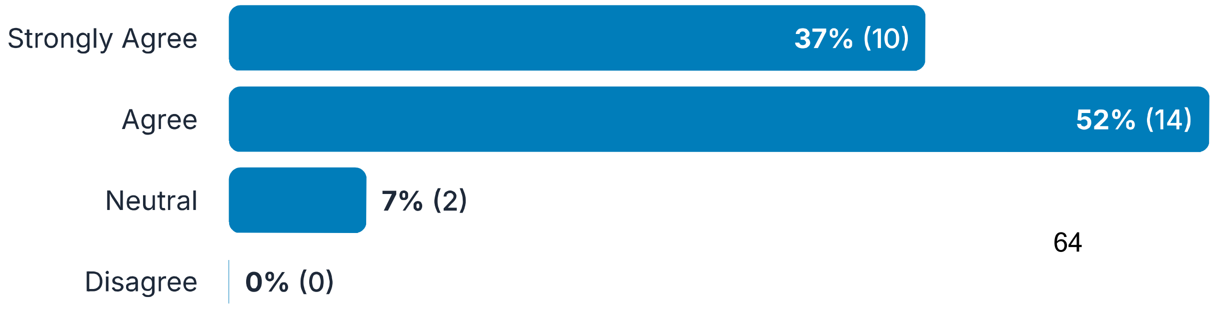

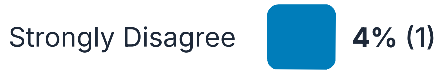

4. Comments:

Responses: 3

> I don't know what the purpose of this one is - it seems to confuse two concepts. #1 you should state your updating methods clearly. Great - we can agree with this one. #2 address possible potentials for error. Surely this is a different point? There are very many possible potential sources of error...

> Perhaps consider including example strategies for mitigation.

> Such a concern exists for all systematic reviews.

5. Updated statement:Plans should be in place to maintain engagement with relevant interest-holders to ensure consistency in informing relevant aims, objectives, and methodologies for the duration of the living evidence synthesis (e.g., via public comment, panel involvement, structured feedback, or other appropriate representation). This may not be possible with every update given time and resource constraints of the topic, so consider:· how often to convene interest-holders,· who to involve if not all representatives will be included each time, and· different thresholds for different update types.Remark: All interest-holder perspectives should be represented when feasible (e.g., patient representatives, members of the public, caregivers, clinicians, providers), including diverse viewpoints on the topic that may be relevant to a living evidence synthesis (such as for ethical or legal considerations).

Responses: 27

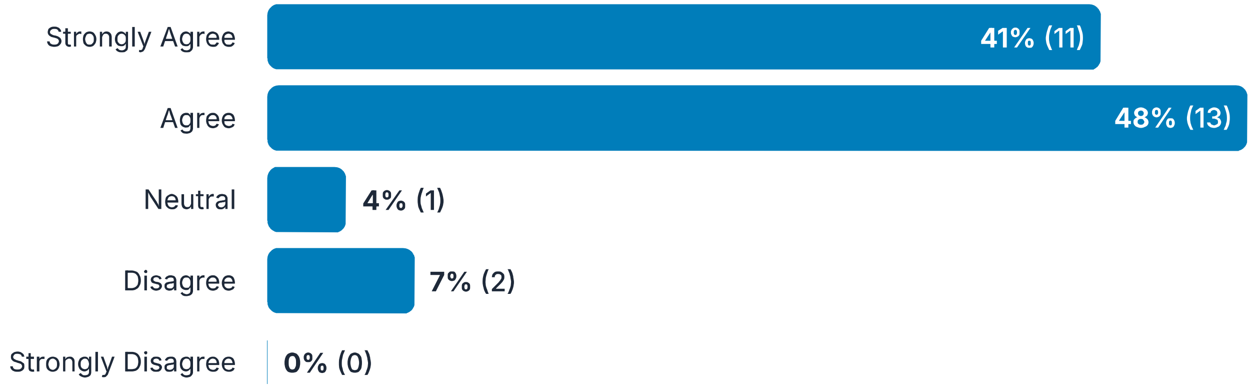

#### 6. Comments:

Responses: 6

> Note that, if this is not feasible for updates to the LES project, it's most important for this to be included as early as possible in LES planning.

> Why "interest-holders" and not "stake-holders"? And how is this unique to living reviews/recommendations? Or is that not the intention?

> I am skeptical on putting too much burden on evidence synthesizers so how about softening the remark a bit, by adding relevant: Remark: All relevant interest-holder perspectives should be represented when feasible (e.g., patient representatives, members of the public, caregivers, clinicians, providers), including diverse viewpoints on the topic that may be relevant to a living evidence synthesis (such as for ethical or legal considerations)

> Interest holders may (and ideally should) be vast. It might be helpful to add a note about considering all potential interest holders. These would include those who are the immediate intended recipients of the LES as well as those who are not the immediate intended recipients but who may use the LES, such as decision-makers from other regions/countries including LMICs..

> Panel members should be clearly distinguished from other interest-holders

> should clearly indicate if interest-holders were not involved (or less involved) in a given update and reasons why (e.g., time or resource constraints)

7. Updated statement:If possible, validated and justified software and/or automation tools should be considered to assist with various phases of a living evidence synthesis (e.g., update searches and screening, awareness for when updates are warranted).Previous statement:If possible, validated and justified software and/or automation tools should be used to assist in various phases of a living evidence synthesis (e.g., update searches and screening, awareness for when updates are warranted).

Responses: 27

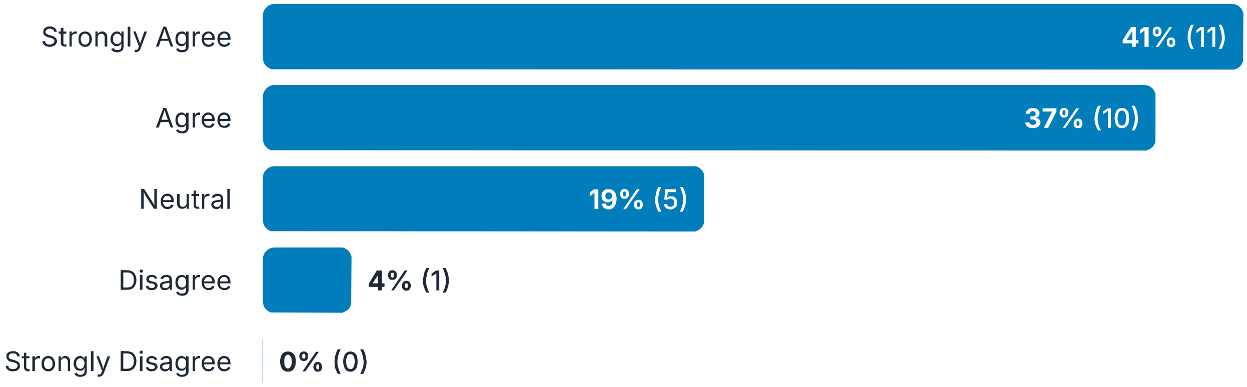

---

8. Comments:

Responses: 4

> What does "justified" mean here? And isn't reliability also important?

> Could you define "justified"?

> "Should be considered" too vague. What do you mean "if possible"? What do you mean "should be considered"d implemented if judged suitable"

> may need to define "validated" and "justified"

#### Draft consensus statements for living evidence syntheses

Conduct Standards for conducting a living evidence synthesis, including initial considerations for set up, ongoing maintenance, and funding/resources specific to the living mode. As with other types of evidence syntheses, living evidence syntheses should be planned in advance and clearly documented in a published protocol. Any differences from protocol should be transparently reported.

9. Updated statement:The living evidence synthesis and downstream living evidence products should be structured in such a way as to maximize

Responses: 27

interoperability.Remark: This may include adherence to Fast Healthcare Interoperability Resources (FHIR) standard, proper tagging and coding of relevant information, or other considerations as applicable. Depending on expertise of the team as well as functionality of available software and tools, this may be achieved by involving a developer or informatics specialist in the process.Previous statement:Consistency should be ensured throughout the living evidence synthesis, including proper tagging and coding of relevant information as appropriate. Depending on expertise of the team as well as functionality of available software and tools, this may be achieved by involving a developer or informatics specialist in the process.

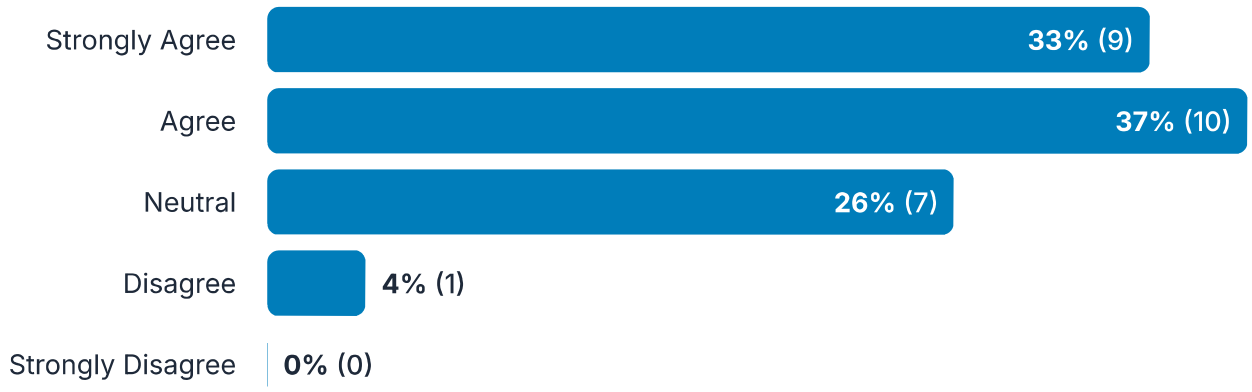

---

#### 10. Comments:

Responses: 5

> A few examples in the remark would be helpful - e.g., in the statement "proper tagging and coding of relevant information" a couple of examples would make the remark more informative

> The statement is fine, but you don't mention *open* data? Interoperability doesn't mean much if people can't access the data

> I don't understand interoperability enough to comment usefully.

> I'm not sure what is meant by interoperability.

> I find the statement and the remark to be a little disconnected. The statement appears to be about clarify of message, grouping and structure of the way results are reported. The first example in the remark is very specific, and I wonder if it should go later in the list of examples and go with the simpler examples first. Also a little unclear how the software/tools and information specialists connects to the statement? If this about managing searches so that like studies are tagged? I think this is not clear

#### Draft consensus statements for living evidence syntheses

Conduct Standards for conducting a living evidence synthesis, including initial considerations for set up, ongoing maintenance, and funding/resources specific to the living mode. As with other types of evidence syntheses, living evidence syntheses should be planned in advance and clearly documented in a published protocol. Any differences from protocol should be transparently reported.

11. Updated statement:When possible, a database management approach should be used to maintain structured, reproducible data in the living evidence synthesis, ensuring transparency, organization, and ease of updating.Previous statement:When possible, a structured, reproducible database management approach should be used to maintain data in the living evidence synthesis, ensuring transparency, organization, and ease of updating.Remark: This allows for easier incorporation of real-world data or unpublished literature (if desired) to supplement available data until peer-reviewed evidence is published, potentially representing a wider range of patient populations where peer-reviewed evidence is lacking. Data should be tagged as unpublished and not peer reviewed.

Responses: 27

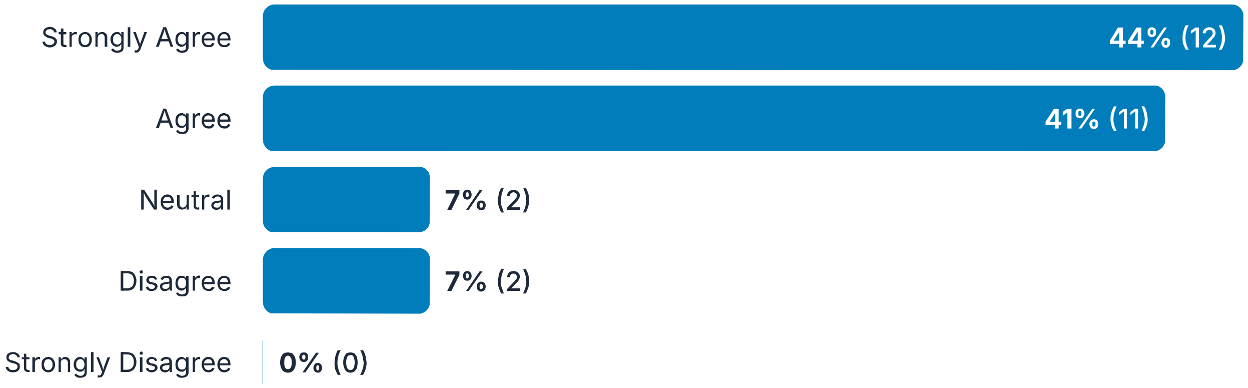

12. Comments:

Responses: 4

> This may be readily obtained from the statement, but could also add consistency in data across updates of the review is required. Also, sometimes, we may consider that we would like to consider additional potential effect modifiers in an update of the review - in such a case, we need to go back to the previous studies and abstract this information for consistency across all reviews and usability of all information in the analysis. Also, another example would be that we used a different tool for assessing quality in the included studies in the first few updates, but then because methodology evolves in meta-analysis quickly, we may need to use a different more advanced approach in the assessment of quality of studies (e.g. risk of bias v1 vs. risk of bias v2 Cochrane tool)

> Using real-world data and tagging the source/type of data is an important component of this element and should not be removed completely.

> What is a "database management approach"? Use of a reliable and efficient relational database?

> I don't think this is useful without identifying how folks should identify an approach that would achieve these goals.

---

##### 13. Updated statement:As living evidence syntheses are time-sensitive and necessitate ongoing investment of resources, efforts should be made to develop joint living evidence syntheses with relevant organizations to allow for pooling funding and resources.Remark: A carefully crafted memorandum of understanding between groups should be mutually agreed upon and followed.Previous statement:As living evidence syntheses are time-sensitive and necessitate ongoing investment of resources, efforts should be made to develop joint living evidence syntheses with relevant organizations to allow for pooling funding and resources.Remark: A carefully crafted memorandum of understanding between groups should be mutually agreed upon and followed.Remark: An authoring tool that allows for language translation could facilitate collaboration and harmonization.

Responses: 27

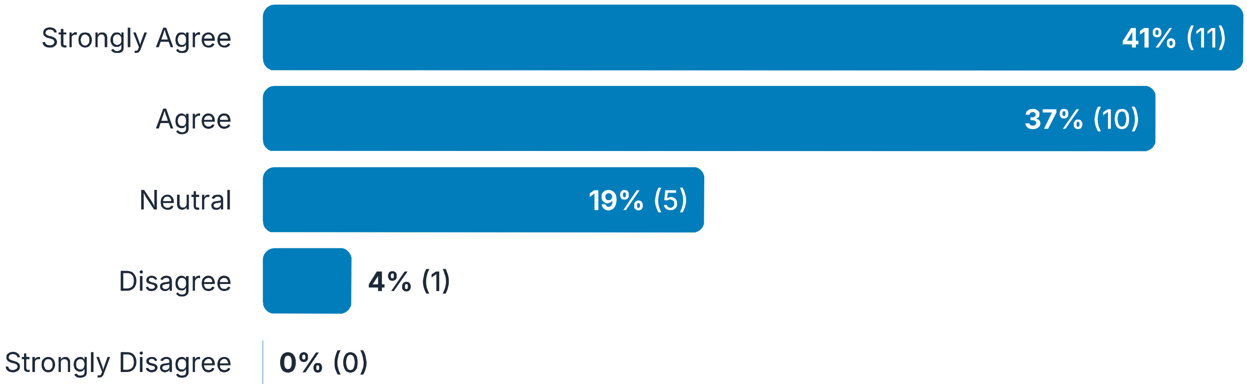

---

##### 14. Comments:

Responses: 2

> "between" refers to 2 groups only. ""among" is >2.

> Is this within scope of what you are trying to do?

#### Draft consensus statements for living evidence syntheses

Reporting The reporting of systematic reviews that adopt a living methodology has been addressed by the PRISMA-LSR group; however, some elements relevant to reporting of broader evidence synthesis are included in this Delphi. As with other types of evidence syntheses, absence of or deviation from a priori thresholds should be justified transparently.

15. Updated statement:A quality living evidence synthesis should serve as the foundation for any downstream living evidence products.Remark: Some evidence products may integrate the evidence synthesis into the same publication, while others publish separately (e.g., clinical practice guidelines).Previous statement:A quality living evidence synthesis should serve as the foundation for any downstream evidence products, such as clinical practice guidelines or health technology assessments.Remark: Some evidence products integrate the evidence synthesis into the same publication, while others publish separately.

Responses: 27

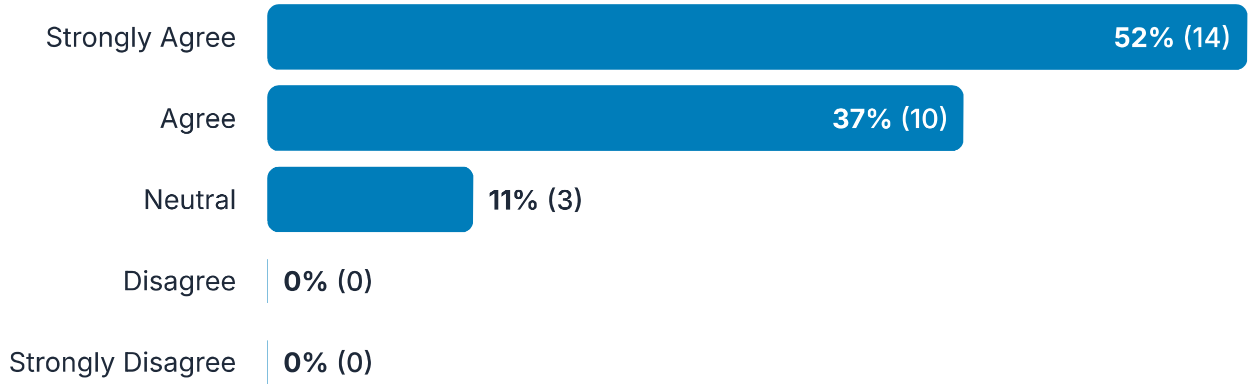

#### 16. Comments:

Responses: 5

> "Downstream" is jargon-ish. Why the remark? and why are CPGs used as an example of separate publishing? That seems to me to be independent of the topic area.

> I feel like this should go much earlier.

> I don't understand "evidence products" well enough to comment.

> I would add "Some evidence based products....."

> Is there a better term to use rather than "quality" - e.g., methodologically sound, or rigorous, or well-conducted, or well-reported...?

---

#### 17. Updated statement:The unit of update should be customized to the living evidence synthesis or product, as it may vary; the entire published version need not be updated each time.Remark: For example, a single recommendation within a living guideline, a conclusion for one condition within a living health technology assessment, or an individual section within a living systematic review.Previous statement:The unit of update should be specific to the living evidence synthesis, as it will be different for each type of evidence synthesis (e.g., a living guideline as opposed to a living health technology assessment); the entire document need not be updated each time.

Responses: 27

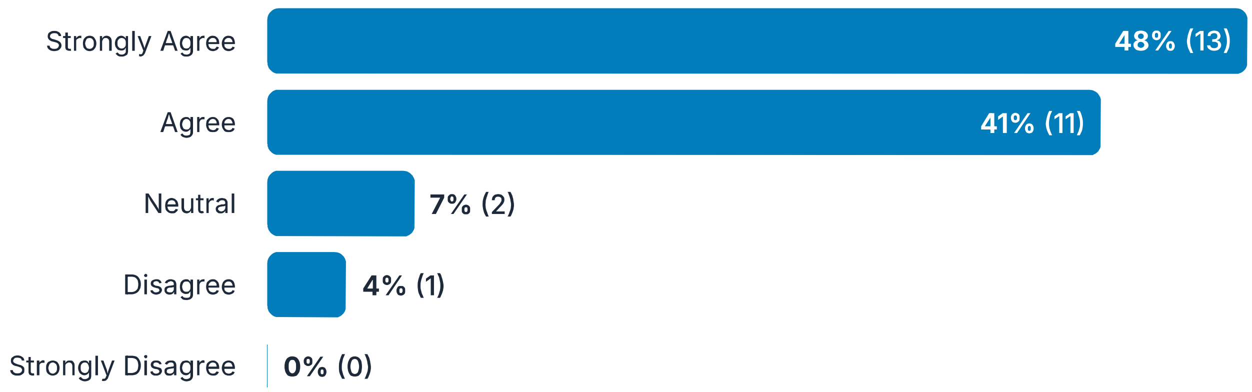

---

#### 18. Comments:

Responses: 5

Sometimes the wording isn't clear as to whether it's talking about the living evidence synthesis, or a downstream product. Does this guidance also cover 3rd party products?

Perhaps it would be better wording to say that the unit of update is the question (not the recommendation in the associated CPG). A recommendation within a CPG incorporates more than just the evidence synthesis; there are other criteria/judgments that go into formulating a recommendation (e.g., EtD criteria)

I don't understand "unit of update" well enough to comment.

or specific outcomes within a living systematic review?

Suggested edit to improve readability: The unit of update for each living evidence synthesis or product should be customized as it can vary; the entire published version need not be updated each time.

#### Draft consensus statements for living evidence syntheses

Publishing This section includes considerations for publishing a living document to allow for transparency as the living evidence synthesis evolves.

19. **Updated statement:Plans should be in place to ensure that updates are published as soon as the pre-defined update threshold is met, given constraints of the synthesis and publication process.Remark: For increased transparency, the evidence synthesis should display a threshold (visual or otherwise) for when the synthesis will be updated, as well as an estimated update timeline specific to the synthesis process.Previous statement:Plans should be in place...(statement remains the same).Remark: For example, guidelines or health technology assessments typically require input of a panel or committee to make decisions, and therefore cannot be immediately updated, but readers could be made aware of any pending updates.Remark: For increased transparency, the evidence synthesis should display a threshold (visual or otherwise) for when the synthesis will be updated, as well as an estimated update timeline specific to the synthesis process.**

Responses: 27

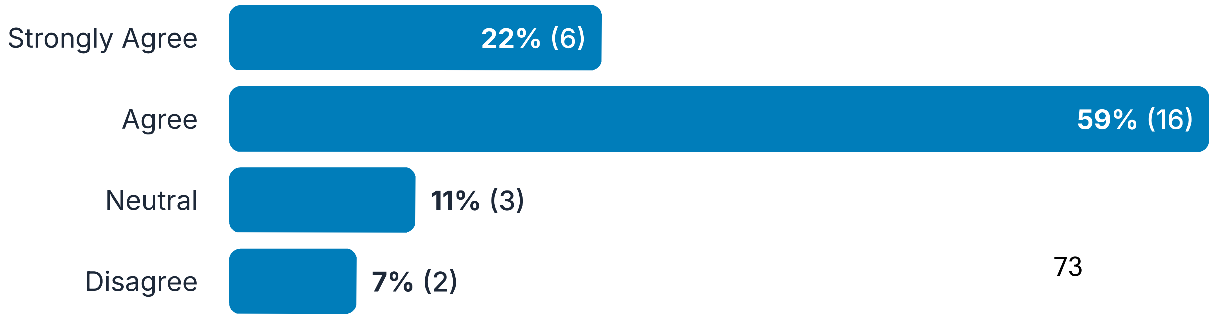

Strongly Disagree | **0% (0)**

#### 20. Comments:

Responses: 8

> Sometimes it can be helpful to partner with a journal that agrees to publish updates.

> "threshold" implies a quantitative synthesis. What if review results are in narrative form?

> I strongly agree with the main statement, but I don't think the remark fits in here - it seems more related to the transparency about the threshold at which a review update is triggered?

> Thresholds are very hard to set, rather transparent criteria for updating, so I think the remark is problematic. For living guidelines one threshold could relate to direction and strength of recommendations and that is easy. But it does not cover all reasons for updating a living guideline (e.g. new recommendation, changed baseline risk estimates or risk stratification of patients etc etc. We have real life examples to confirm these

> Threshold sounds like a date. If that's what you mean don't think it's a great idea.

> I believe we should encourage publication when the incorporation of new available evidence changes the conclusions of the synthesis. This should not be only based on a predefined update threshold.

> Does this need to align better with the earlier item about not publishing unless there have been substantial changes? (e.g., you may meet the threshold for updating, such as time elapsed, but not have sufficient change to warrant publication)

> While a good aim, I'm not sure how feasible it is to estimate this "as well as an estimated update timeline specific to the synthesis process"

#### Draft consensus statements for living evidence syntheses

Implementation/Appraisal Other considerations for ongoing use of the living model.

#### 21. Updated statement:Sufficient detail and transparency should be provided in the living evidence synthesis to ensure accurate and appropriate evidence translation into evidence products used at the point of care (e.g., insurance policy, electronic health record, decision aid).Remark: Though this is important in traditional evidence synthesis, it is of particular importance for

Responses: 27

the living process due to potential evolution of the evidence synthesis.Previous statement:Development of a standards-based living evidence synthesis should ensure consistency for any evidence products used at the point of care (e.g., insurance policy, electronic health record, decision aid, etc.).

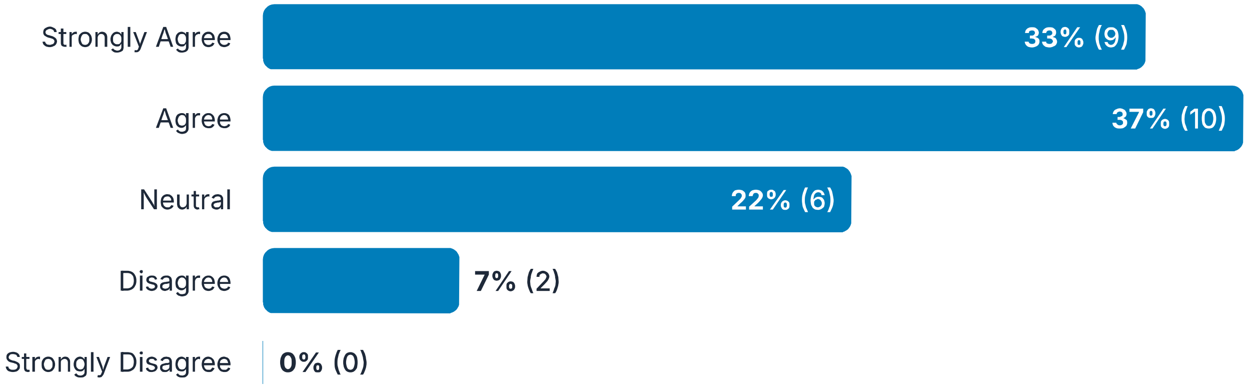

#### 22. Comments:

Responses: 8

> Not all living evidence syntheses are used "point of care"

> I don't fully understand this statement. Do you mean sufficient detail/transparency in the methods? How much detail is reported in the results?

> Note that at the start of a living review/guideline, one always has a "traditional" review/GL. Why is this more important if you plan frequent updates/ie living?.

> I find the remark a bit unclear; potential evolution in what way? What is important is to clarify for improvers/implementert to know what is new and what the implications are for implementation. But describing implementability issues in a guideline adds a lot of work. So I think the statement needs to reflect this; ....in the living evidence synthesis concerning what has changed in an update, to ensure accurate...

> It is not possible for the synthesis to ensure products produced by others are accurate and appropriate.

> I believe that insurance policies, electronic health records, and decision aids are not appropriate examples of "evidence-based products" used at the point of care. I suggest mentioning others, such as summaries for institutional policies, clinical pathways, or decision aids (ej. option grids) as more suitable examples.

> I see this as somewhat problematic as translation into evidence products could be highly variable. How will the authors of the LES know what detail is needed?

> It would be good to have a non-clinical public health example here, or maybe amendment to the language as

> 'point of care' does not relate to broader population health interventions (e.g. settings like schools, childcare, community)

---

23. Updated statement:Where available, evidence synthesis appraisal tools specific to the living mode should be used to assess the quality of living evidence syntheses.Remark: Where unavailable, appraisal tools for traditional evidence syntheses may be modified as appropriate and applied to living evidence syntheses (e.g., adapting AMSTAR 2 to assess methodological quality or AGREE-II for reporting quality).Previous statement:Where available, evidence synthesis appraisal tools specific to the living mode should be used to assess the trustworthiness of living evidence syntheses.Remark: Where unavailable, appraisal tools for traditional evidence syntheses may be modified as appropriate and applied to living evidence syntheses.

Responses: 27

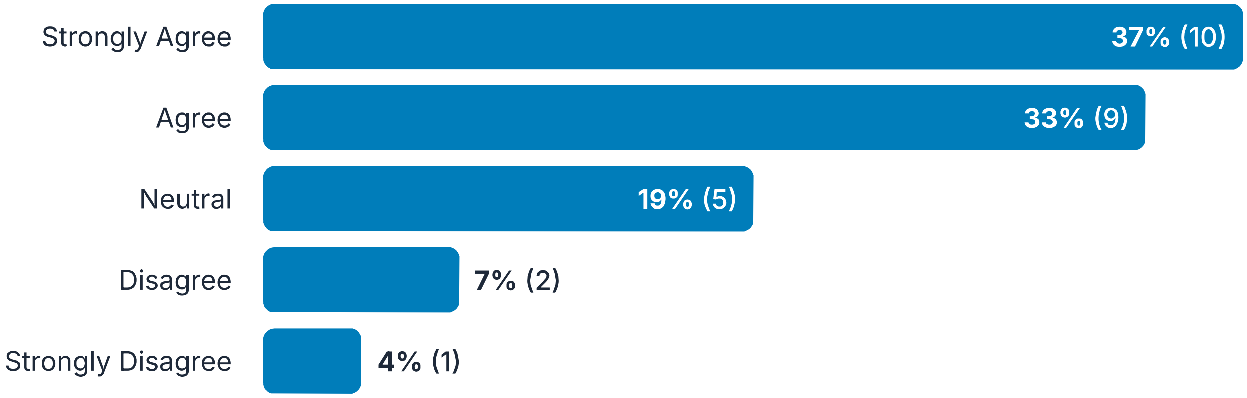

---

24. Comments:

Responses: 7

> Another example to assess methodological quality of network meta-analysis can be the RoB-NMA: https://www.bmj.com/content/388/bmj-2024-079839

> I don't think AGREE II is relevant since it is a tool to assess guideline quality (not SR quality like AMSTAR 2). Also including language about which domains of AMSTAR 2 would be important to modify for the living mode would be helpful.

> AMSTAR is quality of a review; AGREE-II is for guidelines and is both methods quality AND reporting quality. Suggest you mention tools specific for living reviews/GLs

> And these assessments published alongside the LES?

> Use of such tools should be discretionary.

> I don't think it's appropriate to make this recommendation, as there is no AGREE tool for living guidelines or AMSTAR for living systematic reviews. Perhaps we should limit the recommendation to the PRISMA extension for living systematic reviews (LSRs).

> Who is responsible for assessing quality of the LES? Is it the LES authors or those using the LES? Or does this item imply that LES authors should follow methods that align with appraisal tools?

### Appendix 7. Status update.

#### Version history

| Published | Title | Stage | Authors | Version |
|---|---|---|---|---|
| **2024 Jan 08** **Hide revisions** | Electronic cigarettes for smoking cessation | Review | Nicola Lindson, Ailsa R Butler, Hayden McRobbie, Chris Bullen, Peter Hajek, Rachna Begh, Annika Theodoulou, Caitlin Notley, Nancy A Rigotti, Tari Turner, Jonathan Livingstone-Banks, Tom Morris, Jamie Hartmann-Boyce | **https://doi.org/10.1002/14651858.CD010216.pub8** |

| Revision date | Event | Description |
|---|---|---|
| **2024 Jan 08** | New search has been performed | This is a living systematic review. In this update, we incorporate data to 1st July 2023. |
| **2024 Jan 08** | New citation required and conclusions have changed | Certainty of evidence for cessation outcome for comparison with behavioural support/no support upgraded from very low to low |

| Published | Title | Stage | Authors | Version |
|---|---|---|---|---|
| **2022 Nov 17** **Show revisions** | Electronic cigarettes for smoking cessation | Review | Jamie Hartmann-Boyce, Nicola Lindson, Ailsa R Butler, Hayden McRobbie, Chris Bullen, Rachna Begh, Annika Theodoulou, Caitlin Notley, Nancy A Rigotti, Tari Turner, Thomas R Fanshawe, Peter Hajek | **https://doi.org/10.1002/14651858.CD010216.pub7** |
| **2021 Sep 14** **Show revisions** | Electronic cigarettes for smoking cessation | Review | Jamie Hartmann-Boyce, Hayden McRobbie, Ailsa R Butler, Nicola Lindson, Chris Bullen, Rachna Begh, Annika Theodoulou, Caitlin Notley, Nancy A Rigotti, Tari Turner, Thomas R Fanshawe, Peter Hajek | **https://doi.org/10.1002/14651858.CD010216.pub6** |
| **2021 Apr 29** **Show revisions** | Electronic cigarettes for smoking cessation | Review | Jamie Hartmann-Boyce, Hayden McRobbie, Ailsa R Butler, Nicola Lindson, Chris Bullen, Rachna Begh, Annika Theodoulou, Caitlin Notley, Nancy A Rigotti, Tari Turner, Thomas R Fanshawe, Peter Hajek | **https://doi.org/10.1002/14651858.CD010216.pub5** |
| **2020 Oct 14** **Show revisions** | Electronic cigarettes for smoking cessation | Review | Jamie Hartmann-Boyce, Hayden McRobbie, Nicola Lindson, Chris Bullen, Rachna Begh, Annika Theodoulou, Caitlin Notley, Nancy A Rigotti, Tari Turner, Ailsa R Butler, Thomas R Fanshawe, Peter Hajek | **https://doi.org/10.1002/14651858.CD010216.pub4** |
| **2016 Sep 13** **Show revisions** | Electronic cigarettes for smoking cessation | Review | Jamie Hartmann-Boyce, Hayden McRobbie, Chris Bullen, Rachna Begh, Lindsay F Stead, Peter Hajek | **https://doi.org/10.1002/14651858.CD010216.pub3** |
| **2014 Dec 17** **Show revisions** | Electronic cigarettes for smoking cessation and reduction | Review | Hayden McRobbie, Chris Bullen, Jamie Hartmann-Boyce, Peter Hajek | **https://doi.org/10.1002/14651858.CD010216.pub2** |
| **2012 Nov 14** **Show revisions** | Electronic cigarettes for smoking cessation and reduction | Protocol | Hayden McRobbie, Chris Bullen, Peter Hajek | **https://doi.org/10.1002/14651858.CD010216** |

*Status update example.(1)*

### Appendix 8. Published version update.

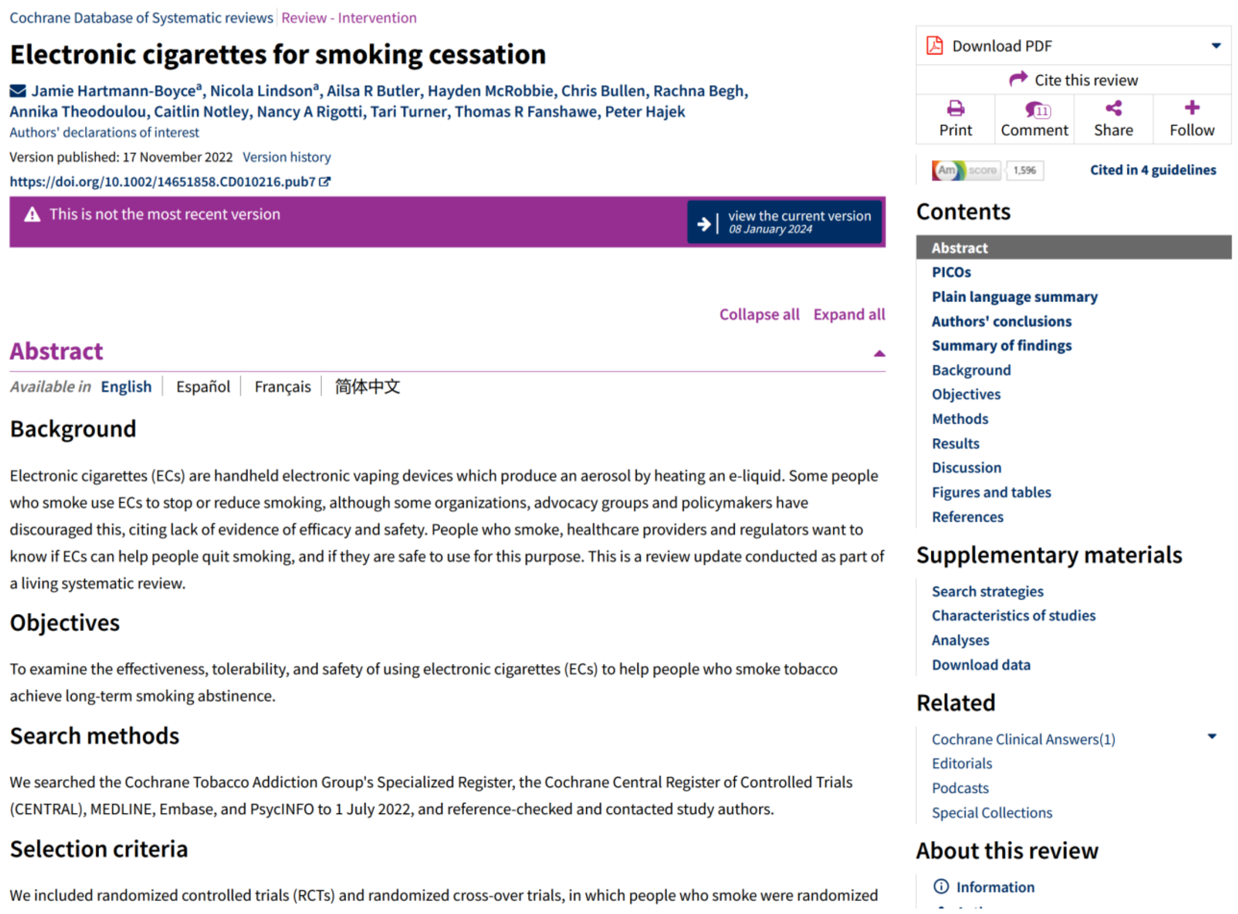

Cochrane Database of Systematic reviews | Review - Intervention

#### Electronic cigarettes for smoking cessation

Jamie Hartmann-Boyce[a], Nicola Lindson[a], Ailsa R Butler, Hayden McRobbie, Chris Bullen, Rachna Begh, Annika Theodoulou, Caitlin Notley, Nancy A Rigotti, Tari Turner, Thomas R Fanshawe, Peter Hajek

Authors' declarations of interest

Version published: 17 November 2022 Version history

https://doi.org/10.1002/14651858.CD010216.pub7

This is not the most recent version

view the current version 08 January 2024

Collapse all Expand all

##### Abstract

Available in English | Español | Français | 简体中文

###### Background

Electronic cigarettes (ECs) are handheld electronic vaping devices which produce an aerosol by heating an e-liquid. Some people who smoke use ECs to stop or reduce smoking, although some organizations, advocacy groups and policymakers have discouraged this, citing lack of evidence of efficacy and safety. People who smoke, healthcare providers and regulators want to know if ECs can help people quit smoking, and if they are safe to use for this purpose. This is a review update conducted as part of a living systematic review.

###### Objectives

To examine the effectiveness, tolerability, and safety of using electronic cigarettes (ECs) to help people who smoke tobacco achieve long-term smoking abstinence.

###### Search methods

We searched the Cochrane Tobacco Addiction Group's Specialized Register, the Cochrane Central Register of Controlled Trials (CENTRAL), MEDLINE, Embase, and PsycINFO to 1 July 2022, and reference-checked and contacted study authors.

###### Selection criteria

We included randomized controlled trials (RCTs) and randomized cross-over trials, in which people who smoke were randomized

Download PDF

Cite this review

Print | Comment | Share | Follow

1,596 | Cited in 4 guidelines

###### Contents

Abstract
PICOs
Plain language summary
Authors' conclusions
Summary of findings
Background
Objectives
Methods
Results
Discussion
Figures and tables
References

###### Supplementary materials

Search strategies
Characteristics of studies
Analyses
Download data

###### Related

Cochrane Clinical Answers(1)
Editorials
Podcasts
Special Collections

###### About this review

Information

*Published version update example.(2)*
